## Supplementary Figures and Tables for "Hierarchical machine learning predicts geographical origin of *Salmonella* within four minutes of sequencing"

### Supplementary Figures + Tables

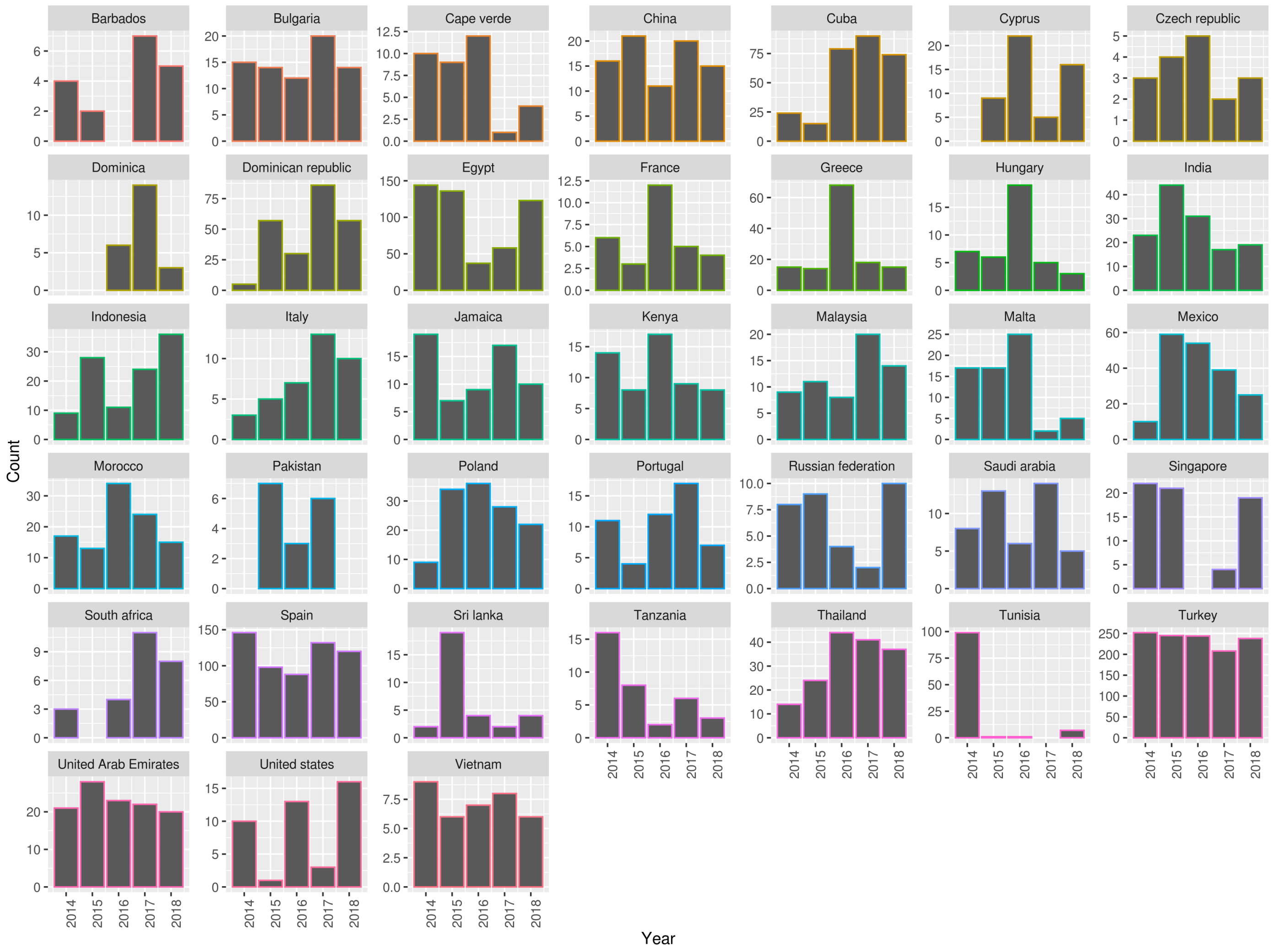


**Figure S1.** Summary of *S*. Enteritidis isolates collected by the UKHSA from UK clinical patients who recently reported foreign travel to 38 individual country classes. Each panel contains a bar chart of isolate counts per year per country class.

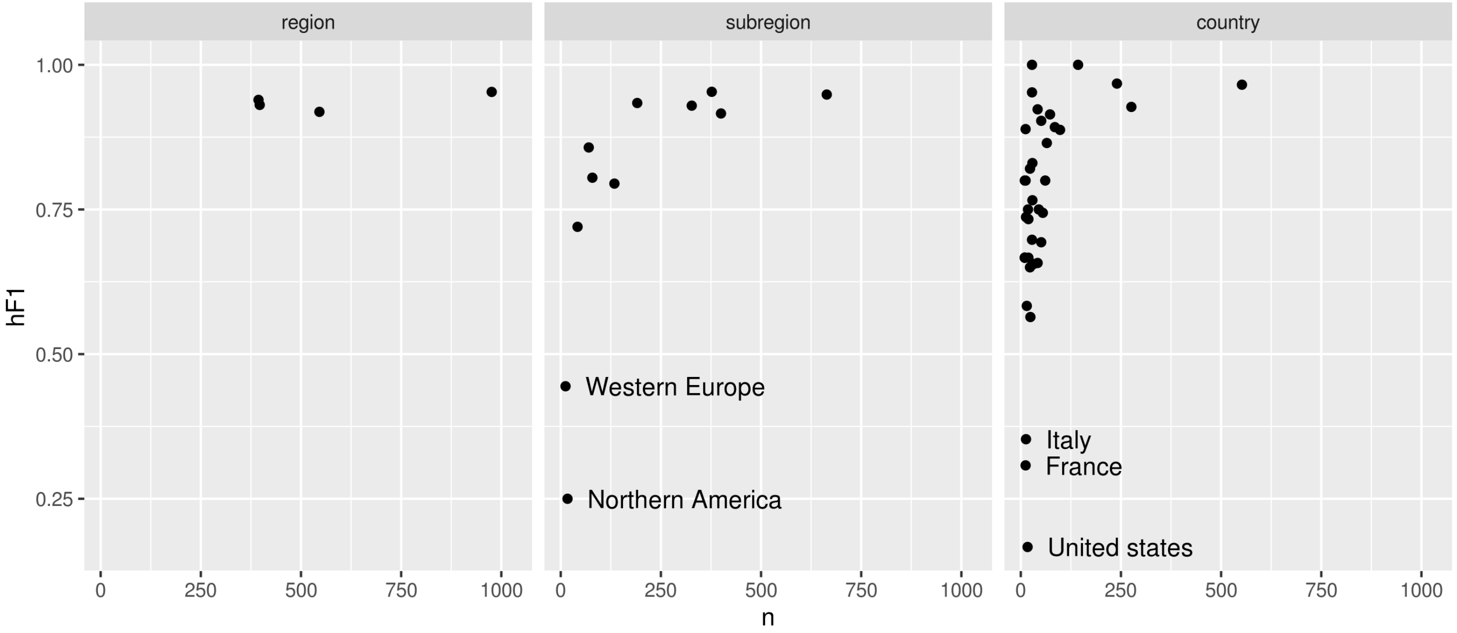


**Figure S2.** Scatter plots summarising the number of samples (x axis) vs hF1 score (y axis) generated by the optimised hML model. The three hierarchical levels (country/subregion/region) are represented by a separate plot panels. Each point is the hF1 score per class at that hierarchical level.

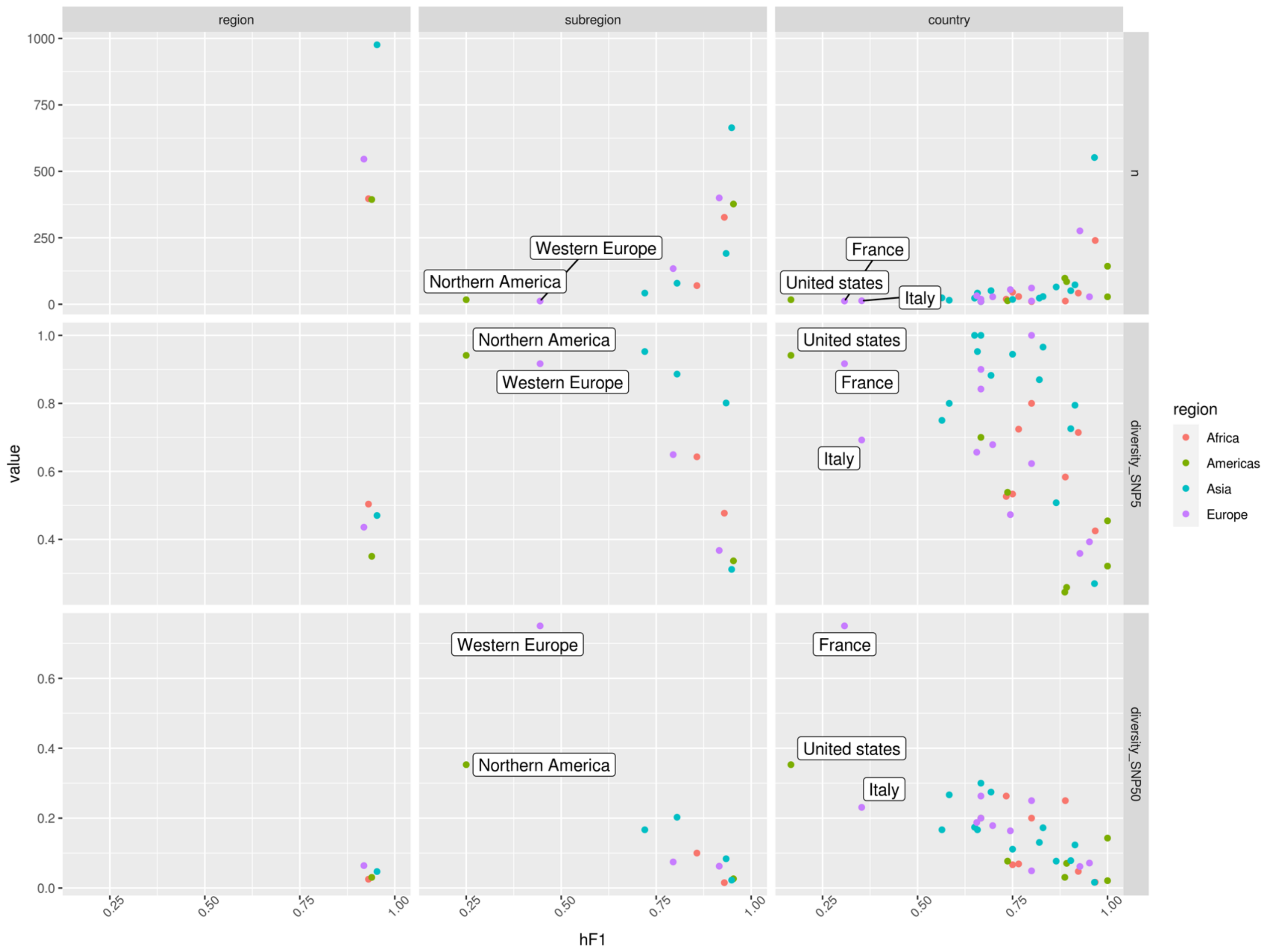


**Figure S3.** Scatter plots summarising hF1 score (x axis) vs. number of samples/genomic diversity at SNP5/genomic diversity at SNP50 (y-axis – top/middle/bottom). Genomic diversity was calculated by dividing the number of single linkage clusters at SNP5/SNP50 (SNP Address) by the total numbers of samples per class. Points were coloured according to region (Africa: red, Americas: green, Asia: blue, Europe: purple).

**Table S1.** Summary statistics for a 'flat' Random Forest multi-class classifier. The classier was trained and tested using a 75%-25% split using the same random seed as was used for the hML classifier. The dataset was randomly oversampled to address class imbalance. Country classes from the hML model were used as labels (i.e region/subregion were excluded).

| | precision | recall | f1-score | support |
|---|---|---|---|---|
| **Barbados** | 1 | 0.67 | 0.8 | 3 |
| **Bulgaria** | 0.22 | 0.25 | 0.24 | 8 |
| **Cape Verde** | 1 | 0.6 | 0.75 | 5 |
| **China** | 0.7 | 0.64 | 0.67 | 11 |
| **Cuba** | 1 | 1 | 1 | 36 |
| **Cyprus** | 0.5 | 0.5 | 0.5 | 6 |
| **Czech republic** | 0 | 0 | 0 | 2 |
| **Dominica** | 0 | 0 | 0 | 3 |
| **Dominican republic** | 0.86 | 0.96 | 0.91 | 25 |
| **Egypt** | 0.89 | 0.98 | 0.94 | 60 |
| **France** | 0 | 0 | 0 | 3 |
| **Greece** | 0.88 | 0.5 | 0.64 | 14 |
| **Hungary** | 0 | 0 | 0 | 5 |
| **India** | 0.7 | 0.54 | 0.61 | 13 |
| **Indonesia** | 0.71 | 0.92 | 0.8 | 13 |
| **Italy** | 0 | 0 | 0 | 3 |
| **Jamaica** | 1 | 1 | 1 | 7 |
| **Kenya** | 0.5 | 0.71 | 0.59 | 7 |
| **Malaysia** | 0.44 | 0.57 | 0.5 | 7 |
| **Malta** | 1 | 0.86 | 0.92 | 7 |
| **Mexico** | 0.95 | 0.9 | 0.93 | 21 |
| **Morocco** | 0.82 | 0.82 | 0.82 | 11 |
| **Pakistan** | 1 | 0.5 | 0.67 | 2 |
| **Poland** | 0.41 | 0.47 | 0.44 | 15 |
| **Portugal** | 0.67 | 0.29 | 0.4 | 7 |
| **Russian federation** | 1 | 0.67 | 0.8 | 3 |
| **Saudi arabia** | 0.5 | 0.67 | 0.57 | 6 |
| **Singapore** | 0.5 | 0.17 | 0.25 | 6 |
| **South africa** | 1 | 0.5 | 0.67 | 2 |
| **Spain** | 0.69 | 0.99 | 0.81 | 69 |
| **Sri Lanka** | 0.8 | 0.8 | 0.8 | 5 |
| **Tanzania** | 1 | 0.67 | 0.8 | 3 |
| **Thailand** | 0.93 | 0.78 | 0.85 | 18 |
| **Tunisia** | 1 | 0.91 | 0.95 | 11 |
| **Turkey** | 0.96 | 0.93 | 0.95 | 138 |
| **Arab Emirates** | 0.87 | 0.81 | 0.84 | 16 |
| **United States** | 0 | 0 | 0 | 4 |
| **Vietnam** | 1 | 0.5 | 0.67 | 4 |
| **accuracy** | 0.82 | | | 579 |
| **macro avg** | 0.67 | 0.58 | 0.61 | 579 |
| **weighted avg** | 0.81 | 0.82 | 0.8 | 579 |

**Table S2.** Summary of the all-vs-all comparison of classifier vs resampler models used to identify the most suitable combinations for feature selection and additional optimisation. The implemented resampling schemes included downsampling (smallest class), upsampling (largest class), resampling to the mean count of all classes and hierarchically aware implementation for the previously described samplers. Hierarchically aware resampling was developed using in-house scripts to iteratively apply a resampler to each of the lowest levels of the hierarchy (country) before passing the resampled data to higher levels in the hierarchy for further resampling. Abbreviations; RF: Random Forest, KNN: K Nearest Neighbour, SVM: Support Vector Machine, GNB: Gaussian Naïve Bayes and XGB: Xtreme Gradient Boosting.

| classifier | resampler | macroR | macroP | macroF1 | microR | microP | microF1 | hR | hP | hF1 | hAcc | time |
|---|---|---|---|---|---|---|---|---|---|---|---|---|
| **RF** | **NoResampling** | 0.717 | 0.709 | 0.713 | 0.857 | 0.821 | 0.838 | 0.849 | 0.892 | 0.870 | 0.870 | 110.872 |
| **RF** | **RandomUnderSampler** | 0.570 | 0.662 | 0.612 | 0.511 | 0.699 | 0.590 | 0.487 | 0.775 | 0.598 | 0.582 | 440.603 |
| **RF** | **RandomOverSampler** | 0.740 | 0.744 | 0.742 | 0.857 | 0.841 | 0.849 | 0.850 | 0.903 | 0.876 | 0.877 | 801.726 |
| **RF** | **Balancing_mean** | 0.708 | 0.703 | 0.706 | 0.756 | 0.794 | 0.774 | 0.745 | 0.870 | 0.803 | 0.794 | 840.412 |
| **RF** | **HierMean** | 0.690 | 0.649 | 0.669 | 0.683 | 0.690 | 0.686 | 0.666 | 0.788 | 0.722 | 0.712 | 2490.542 |
| **ExtraTrees** | **NoResampling** | 0.738 | 0.727 | 0.733 | 0.868 | 0.826 | 0.846 | 0.861 | 0.897 | 0.878 | 0.881 | 94.545 |
| **ExtraTrees** | **RandomUnderSampler** | 0.650 | 0.568 | 0.606 | 0.606 | 0.605 | 0.605 | 0.572 | 0.707 | 0.633 | 0.633 | 382.681 |
| **ExtraTrees** | **RandomOverSampler** | 0.732 | 0.733 | 0.733 | 0.862 | 0.828 | 0.845 | 0.855 | 0.897 | 0.876 | 0.880 | 808.948 |
| **ExtraTrees** | **Balancing_mean** | 0.729 | 0.697 | 0.713 | 0.785 | 0.796 | 0.790 | 0.774 | 0.872 | 0.820 | 0.807 | 911.307 |
| **ExtraTrees** | **HierMean** | 0.698 | 0.672 | 0.685 | 0.732 | 0.739 | 0.735 | 0.718 | 0.827 | 0.769 | 0.746 | 2463.553 |
| **KNN** | **NoResampling** | 0.589 | 0.582 | 0.585 | 0.753 | 0.701 | 0.726 | 0.737 | 0.805 | 0.770 | 0.755 | 31.787 |
| **KNN** | **RandomUnderSampler** | 0.446 | 0.476 | 0.461 | 0.373 | 0.531 | 0.438 | 0.332 | 0.603 | 0.428 | 0.385 | 360.199 |
| **KNN** | **RandomOverSampler** | 0.702 | 0.541 | 0.611 | 0.744 | 0.605 | 0.667 | 0.710 | 0.717 | 0.714 | 0.712 | 664.673 |
| **KNN** | **Balancing_mean** | 0.646 | 0.474 | 0.547 | 0.586 | 0.477 | 0.526 | 0.522 | 0.572 | 0.546 | 0.538 | 761.527 |
| **KNN** | **HierMean** | 0.618 | 0.447 | 0.519 | 0.548 | 0.432 | 0.483 | 0.500 | 0.543 | 0.521 | 0.520 | 2160.467 |
| **SVM** | **NoResampling** | 0.474 | 0.509 | 0.491 | 0.674 | 0.669 | 0.671 | 0.655 | 0.779 | 0.711 | 0.684 | 1667.747 |
| **SVM** | **RandomUnderSampler** | 0.174 | 0.586 | 0.268 | 0.137 | 0.700 | 0.229 | 0.136 | 0.703 | 0.229 | 0.300 | 377.683 |
| **SVM** | **RandomOverSampler** | 0.661 | 0.643 | 0.652 | 0.682 | 0.691 | 0.686 | 0.664 | 0.779 | 0.717 | 0.732 | 56366.899 |
| **SVM** | **Balancing_mean** | 0.468 | 0.559 | 0.510 | 0.401 | 0.578 | 0.473 | 0.386 | 0.598 | 0.469 | 0.517 | 2471.136 |
| **SVM** | **HierMean** | 0.456 | 0.568 | 0.506 | 0.362 | 0.563 | 0.441 | 0.347 | 0.601 | 0.440 | 0.489 | 4029.166 |
| **GNB** | **NoResampling** | 0.557 | 0.489 | 0.521 | 0.734 | 0.579 | 0.648 | 0.706 | 0.706 | 0.706 | 0.706 | 50.790 |
| **GNB** | **RandomUnderSampler** | 0.605 | 0.407 | 0.487 | 0.587 | 0.416 | 0.487 | 0.506 | 0.506 | 0.506 | 0.506 | 366.194 |
| **GNB** | **RandomOverSampler** | 0.552 | 0.481 | 0.514 | 0.731 | 0.576 | 0.644 | 0.703 | 0.703 | 0.703 | 0.703 | 1008.661 |
| **GNB** | **Balancing_mean** | 0.571 | 0.454 | 0.506 | 0.696 | 0.534 | 0.604 | 0.667 | 0.667 | 0.667 | 0.667 | 896.942 |
| **GNB** | **HierMean** | 0.585 | 0.451 | 0.509 | 0.681 | 0.516 | 0.587 | 0.653 | 0.653 | 0.653 | 0.653 | 2511.271 |
| **XGB** | **NoResampling** | 0.736 | 0.689 | 0.712 | 0.860 | 0.786 | 0.821 | 0.851 | 0.869 | 0.860 | 0.863 | 4864.929 |
| **XGB** | **RandomUnderSampler** | 0.628 | 0.524 | 0.571 | 0.579 | 0.536 | 0.557 | 0.535 | 0.639 | 0.583 | 0.591 | 3944.032 |

| | | | | | | | | | | | |
|---|---|---|---|---|---|---|---|---|---|---|---|
| **XGB** | **RandomOverSampler** | 0.767 | 0.711 | 0.738 | 0.873 | 0.799 | 0.835 | 0.864 | 0.877 | 0.870 | 0.874 | 14097.291 |
| **XGB** | **Balancing_mean** | 0.762 | 0.661 | 0.708 | 0.816 | 0.735 | 0.773 | 0.801 | 0.829 | 0.815 | 0.813 | 5608.895 |
| **XGB** | **HierMean** | 0.746 | 0.641 | 0.689 | 0.769 | 0.685 | 0.725 | 0.749 | 0.791 | 0.769 | 0.761 | 7156.246 |

**Table S3.** The 2313 *S.* Enteritidis sample collection used for training and testing of the hML classifier presented in the main manuscript. All samples were collected by the UKHSA as a part of their genomic surveillance initiative. Recent recorded travel was collected as a part of the UKHSA's 'enhanced surveillance' programme. The columns include: SRA accession code, receipt date of isolate, SNP Address, reported country of travel, subregion and region that country lies within based on the UN M49 Standard for regional codes.

| SRA Accession | RECEIPT date | SNP address | Country | Region | Subregion |
|---|---|---|---|---|---|
| **SRR8667277** | 20/02/2019 | 1.2.3.18.180.180.12747 | Barbados | Americas | Latin America and the Caribbean |
| **SRR8691693** | 20/02/2019 | 1.2.3.18.180.180.12776 | Barbados | Americas | Latin America and the Caribbean |
| **SRR1967763** | 21/05/2014 | 1.2.3.18.365.1120.1889 | Barbados | Americas | Latin America and the Caribbean |
| **SRR8369264** | 31/07/2018 | 1.5.11.2045.2843.3591.10052 | Barbados | Americas | Latin America and the Caribbean |
| **SRR6922673** | 27/01/2017 | 1.5.11.2045.2843.3591.6906 | Barbados | Americas | Latin America and the Caribbean |
| **SRR5220509** | 13/05/2015 | 1.5.11.296.296.1951.2805 | Barbados | Americas | Latin America and the Caribbean |
| **SRR6901034** | 21/02/2017 | 1.5.11.296.296.3642.6998 | Barbados | Americas | Latin America and the Caribbean |
| **SRR7892287** | 04/01/2018 | 1.5.11.296.296.3642.9180 | Barbados | Americas | Latin America and the Caribbean |
| **SRR5632870** | 03/05/2017 | 1.5.11.296.296.3734.7233 | Barbados | Americas | Latin America and the Caribbean |
| **SRR3286628** | 08/04/2015 | 1.5.11.296.296.858.1093 | Barbados | Americas | Latin America and the Caribbean |
| **SRR1957747** | 10/09/2014 | 1.1.2.12.12.2866.810 | Bulgaria | Europe | Eastern Europe |
| **SRR1967706** | 01/08/2014 | 1.1.2.12.12.590.1520 | Bulgaria | Europe | Eastern Europe |
| **SRR3285355** | 20/10/2015 | 1.1.2.12.12.590.3896 | Bulgaria | Europe | Eastern Europe |
| **SRR5220458** | 12/10/2015 | 1.1.2.12.12.665.3750 | Bulgaria | Europe | Eastern Europe |
| **SRR7867061** | 25/07/2018 | 1.1.2.12.421.429.6692 | Bulgaria | Europe | Eastern Europe |
| **SRR8272525** | 10/09/2018 | 1.1.2.12.421.4553.11672 | Bulgaria | Europe | Eastern Europe |
| **SRR8249716** | 13/11/2018 | 1.1.2.1326.574.3711.12445 | Bulgaria | Europe | Eastern Europe |
| **SRR6918623** | 05/09/2016 | 1.1.2.1674.2421.3017.5471 | Bulgaria | Europe | Eastern Europe |
| **SRR7292885** | 08/07/2015 | 1.11.48.1059.1734.2035.3050 | Bulgaria | Europe | Eastern Europe |
| **SRR1958539** | 26/09/2014 | 1.11.48.154.1148.1301.1755 | Bulgaria | Europe | Eastern Europe |
| **SRR8711843** | 07/09/2015 | 1.11.48.154.1148.1301.3492 | Bulgaria | Europe | Eastern Europe |
| **SRR1958537** | 23/09/2014 | 1.11.48.154.154.154.154 | Bulgaria | Europe | Eastern Europe |
| **SRR8087178** | 27/09/2018 | 1.11.48.154.4007.5622.12040 | Bulgaria | Europe | Eastern Europe |
| **SRR6920194** | 13/09/2017 | 1.11.48.2338.3182.4113.8403 | Bulgaria | Europe | Eastern Europe |
| **SRR5220649** | 20/06/2016 | 1.11.48.68.2221.2740.4847 | Bulgaria | Europe | Eastern Europe |
| **SRR8548734** | 21/08/2017 | 1.11.660.2325.3119.4003.8046 | Bulgaria | Europe | Eastern Europe |
| **SRR6918310** | 13/09/2017 | 1.11.660.2325.3119.4003.8298 | Bulgaria | Europe | Eastern Europe |
| **SRR6919807** | 03/10/2017 | 1.11.660.2325.3119.4003.8556 | Bulgaria | Europe | Eastern Europe |
| **SRR6900742** | 20/09/2016 | 1.11.82.1708.2484.3104.5707 | Bulgaria | Europe | Eastern Europe |
| **SRR7828419** | 23/07/2018 | 1.2.3.18.175.175.10499 | Bulgaria | Europe | Eastern Europe |
| **SRR7841512** | 18/07/2018 | 1.2.3.18.175.175.3397 | Bulgaria | Europe | Eastern Europe |
| **SRR6900956** | 01/08/2017 | 1.2.3.18.175.175.7972 | Bulgaria | Europe | Eastern Europe |
| **SRR6920506** | 07/09/2017 | 1.2.3.18.175.175.8318 | Bulgaria | Europe | Eastern Europe |
| **SRR6900134** | 13/09/2017 | 1.2.3.18.175.175.8362 | Bulgaria | Europe | Eastern Europe |
| **SRR5220307** | 18/09/2015 | 1.2.3.18.3648.4903.10568 | Bulgaria | Europe | Eastern Europe |
| **SRR6919312** | 31/08/2017 | 1.2.3.18.3648.4903.10613 | Bulgaria | Europe | Eastern Europe |
| **SRR6919277** | 04/07/2017 | 1.2.3.18.3648.4903.10622 | Bulgaria | Europe | Eastern Europe |
| **SRR7527861** | 23/06/2017 | 1.2.3.18.3648.4903.10644 | Bulgaria | Europe | Eastern Europe |
| **SRR8711775** | 11/09/2015 | 1.5.69.1112.1843.2187.3438 | Bulgaria | Europe | Eastern Europe |
| **SRR6900850** | 02/10/2017 | 1.5.69.1112.3214.4170.8574 | Bulgaria | Europe | Eastern Europe |
| **SRR5194331** | 31/08/2016 | 1.5.69.531.2315.2884.5237 | Bulgaria | Europe | Eastern Europe |
| **SRR5216193** | 26/09/2016 | 1.5.69.531.2496.3123.5815 | Bulgaria | Europe | Eastern Europe |
| **SRR8514641** | 12/02/2018 | 1.1.2.80.100.100.7880 | Cape verde | Africa | Sub-Saharan Africa |

| | | | | | |
|---|---|---|---|---|---|
| **SRR6897875** | 13/10/2017 | 1.1.2.80.100.100.8659 | Cape verde | Africa | Sub-Saharan Africa |
| **SRR3284844** | 28/10/2015 | 1.2.3.151.1800.2130.3920 | Cape verde | Africa | Sub-Saharan Africa |
| **SRR8648228** | 12/02/2019 | 1.2.3.18.180.180.10985 | Cape verde | Africa | Sub-Saharan Africa |
| **SRR8585162** | 29/01/2019 | 1.2.3.18.180.180.12684 | Cape verde | Africa | Sub-Saharan Africa |
| **SRR8691506** | 18/02/2019 | 1.2.3.18.180.180.12767 | Cape verde | Africa | Sub-Saharan Africa |
| **SRR8239903** | 12/11/2018 | 1.2.3.18.3256.5644.12128 | Cape verde | Africa | Sub-Saharan Africa |
| **SRR7351348** | 19/11/2015 | 1.2.3.18.365.2452.4069 | Cape verde | Africa | Sub-Saharan Africa |
| **SRR8503738** | 04/04/2018 | 1.2.3.18.365.2876.9775 | Cape verde | Africa | Sub-Saharan Africa |
| **SRR5193507** | 02/08/2016 | 1.21.152.270.2270.2814.5072 | Cape verde | Africa | Sub-Saharan Africa |
| **SRR6901197** | 02/08/2016 | 1.21.152.270.2270.2814.5101 | Cape verde | Africa | Sub-Saharan Africa |
| **SRR1969985** | 11/11/2014 | 1.21.152.270.270.270.1265 | Cape verde | Africa | Sub-Saharan Africa |
| **SRR1959217** | 02/12/2014 | 1.21.152.270.270.270.2518 | Cape verde | Africa | Sub-Saharan Africa |
| **SRR1963361** | 06/01/2015 | 1.21.152.270.270.270.904 | Cape verde | Africa | Sub-Saharan Africa |
| **SRR3585356** | 01/09/2015 | 1.21.260.1114.1845.2190.3444 | Cape verde | Africa | Sub-Saharan Africa |
| **SRR7187213** | 04/09/2015 | 1.21.260.1114.1845.2190.3555 | Cape verde | Africa | Sub-Saharan Africa |
| **SRR5220622** | 18/10/2016 | 1.21.496.1858.2552.3201.6037 | Cape verde | Africa | Sub-Saharan Africa |
| **SRR5215795** | 17/10/2016 | 1.21.496.1858.2552.3201.6051 | Cape verde | Africa | Sub-Saharan Africa |
| **SRR5194243** | 20/10/2016 | 1.21.496.1858.2552.3201.6068 | Cape verde | Africa | Sub-Saharan Africa |
| **SRR7297982** | 26/10/2015 | 1.1.2.1181.2005.2408.3969 | China | Asia | Eastern Asia |
| **SRR5220108** | 04/11/2015 | 1.1.2.1183.2008.2414.3978 | China | Asia | Eastern Asia |
| **SRR6920165** | 08/09/2017 | 1.1.2.174.3169.4092.8348 | China | Asia | Eastern Asia |
| **SRR7962254** | 17/09/2018 | 1.1.2.174.3252.4262.11762 | China | Asia | Eastern Asia |
| **SRR7884713** | 17/07/2018 | 1.1.2.174.3631.4861.10458 | China | Asia | Eastern Asia |
| **SRR5216455** | 06/09/2016 | 1.1.2.206.2441.3043.5516 | China | Asia | Eastern Asia |
| **SRR1969341** | 03/11/2014 | 1.1.2.214.214.214.214 | China | Asia | Eastern Asia |
| **SRR1970088** | 12/11/2014 | 1.1.2.242.242.242.1680 | China | Asia | Eastern Asia |
| **SRR7359045** | 10/05/2018 | 1.1.2.242.3555.4719.9941 | China | Asia | Eastern Asia |
| **SRR8269077** | 13/11/2018 | 1.1.2.2656.4044.5703.12428 | China | Asia | Eastern Asia |
| **SRR1961769** | 18/11/2014 | 1.1.2.291.1436.1655.2275 | China | Asia | Eastern Asia |
| **SRR1968511** | 25/06/2014 | 1.1.2.47.47.47.47 | China | Asia | Eastern Asia |
| **SRR3285284** | 18/06/2015 | 1.1.2.7.1724.2015.2977 | China | Asia | Eastern Asia |
| **SRR6900695** | 04/07/2017 | 1.1.2.7.3085.3931.7722 | China | Asia | Eastern Asia |
| **SRR6922083** | 12/09/2017 | 1.1.2.7.3187.4120.8424 | China | Asia | Eastern Asia |
| **SRR6918629** | 05/10/2017 | 1.1.2.7.3216.4176.8587 | China | Asia | Eastern Asia |
| **SRR6900554** | 14/11/2017 | 1.1.2.7.3216.4260.8873 | China | Asia | Eastern Asia |
| **SRR8503650** | 03/05/2018 | 1.1.2.7.3550.4710.9909 | China | Asia | Eastern Asia |
| **SRR8499260** | 16/05/2018 | 1.1.2.7.3550.4710.9954 | China | Asia | Eastern Asia |
| **SRR7879587** | 07/08/2018 | 1.1.2.7.3550.5315.11345 | China | Asia | Eastern Asia |
| **SRR8515755** | 10/05/2018 | 1.1.2.7.3563.4730.9971 | China | Asia | Eastern Asia |
| **SRR8299424** | 24/08/2018 | 1.1.2.7.3860.5380.11512 | China | Asia | Eastern Asia |
| **SRR5193096** | 29/04/2016 | 1.1.2.7.426.2654.4622 | China | Asia | Eastern Asia |
| **SRR5583845** | 27/02/2017 | 1.1.2.7.426.3647.7005 | China | Asia | Eastern Asia |
| **SRR6918552** | 18/07/2017 | 1.1.2.7.426.4012.8074 | China | Asia | Eastern Asia |
| **SRR1970131** | 25/07/2014 | 1.1.52.75.75.75.75 | China | Asia | Eastern Asia |
| **SRR5193472** | 05/12/2016 | 1.21.92.248.2622.3310.6380 | China | Asia | Eastern Asia |
| **SRR5220952** | 02/06/2015 | 4.41.232.1043.1697.1969.2876 | China | Asia | Eastern Asia |
| **SRR1969195** | 07/05/2014 | 1.1.2.15.1015.1140.1518 | Cuba | Americas | Latin America and the Caribbean |

| | | | | | |
|---|---|---|---|---|---|
| **SRR1970066** | 13/03/2015 | 1.1.2.15.1015.1140.2431 | Cuba | Americas | Latin America and the Caribbean |
| **SRR3049703** | 30/04/2014 | 1.1.2.15.1015.1140.2595 | Cuba | Americas | Latin America and the Caribbean |
| **SRR3049597** | 28/04/2014 | 1.1.2.15.1015.1140.2612 | Cuba | Americas | Latin America and the Caribbean |
| **SRR1963081** | 13/11/2014 | 1.1.2.15.1217.1387.1883 | Cuba | Americas | Latin America and the Caribbean |
| **SRR1966663** | 12/05/2014 | 1.1.2.15.15.15.15 | Cuba | Americas | Latin America and the Caribbean |
| **SRR7884512** | 05/09/2018 | 1.1.2.15.15.3655.11659 | Cuba | Americas | Latin America and the Caribbean |
| **SRR8117007** | 08/10/2018 | 1.1.2.15.15.3655.12114 | Cuba | Americas | Latin America and the Caribbean |
| **SRR9287750** | 12/04/2019 | 1.1.2.15.15.3655.12974 | Cuba | Americas | Latin America and the Caribbean |
| **SRR6922785** | 12/09/2017 | 1.1.2.15.15.3655.8364 | Cuba | Americas | Latin America and the Caribbean |
| **SRR6922486** | 28/09/2017 | 1.1.2.15.15.3655.8596 | Cuba | Americas | Latin America and the Caribbean |
| **SRR8509074** | 12/03/2018 | 1.1.2.15.15.3655.9704 | Cuba | Americas | Latin America and the Caribbean |
| **SRR7285867** | 30/11/2015 | 1.1.2.15.2056.2487.4165 | Cuba | Americas | Latin America and the Caribbean |
| **SRR5216173** | 03/03/2016 | 1.1.2.15.2056.2487.4479 | Cuba | Americas | Latin America and the Caribbean |
| **SRR5193249** | 27/04/2016 | 1.1.2.15.2056.2487.4610 | Cuba | Americas | Latin America and the Caribbean |
| **SRR6900474** | 24/01/2017 | 1.1.2.15.2056.2487.6871 | Cuba | Americas | Latin America and the Caribbean |
| **SRR6918875** | 18/01/2017 | 1.1.2.15.2056.2487.6891 | Cuba | Americas | Latin America and the Caribbean |
| **SRR6921913** | 18/01/2017 | 1.1.2.15.2056.2487.6900 | Cuba | Americas | Latin America and the Caribbean |
| **SRR5220227** | 20/12/2016 | 1.1.2.15.2056.3343.6495 | Cuba | Americas | Latin America and the Caribbean |
| **SRR5194271** | 13/01/2016 | 1.1.2.15.2089.2535.4313 | Cuba | Americas | Latin America and the Caribbean |
| **SRR5215881** | 09/02/2016 | 1.1.2.15.2118.2577.4405 | Cuba | Americas | Latin America and the Caribbean |
| **SRR5193281** | 18/07/2016 | 1.1.2.15.2252.2789.4996 | Cuba | Americas | Latin America and the Caribbean |
| **SRR6922710** | 14/06/2017 | 1.1.2.15.2274.2823.7550 | Cuba | Americas | Latin America and the Caribbean |
| **SRR6898312** | 10/08/2016 | 1.1.2.15.2278.2827.5109 | Cuba | Americas | Latin America and the Caribbean |
| **SRR5193869** | 14/09/2016 | 1.1.2.15.2460.3066.5598 | Cuba | Americas | Latin America and the Caribbean |
| **SRR6919842** | 10/10/2016 | 1.1.2.15.2517.3152.5902 | Cuba | Americas | Latin America and the Caribbean |
| **SRR7506997** | 25/06/2018 | 1.1.2.15.2834.3578.10266 | Cuba | Americas | Latin America and the Caribbean |
| **SRR6918339** | 11/01/2017 | 1.1.2.15.2834.3578.6852 | Cuba | Americas | Latin America and the Caribbean |
| **SRR5585374** | 07/02/2017 | 1.1.2.15.2850.3603.6933 | Cuba | Americas | Latin America and the Caribbean |
| **SRR7298387** | 25/01/2018 | 1.1.2.15.2850.3603.9340 | Cuba | Americas | Latin America and the Caribbean |
| **SRR7439464** | 21/03/2018 | 1.1.2.15.2850.4634.9722 | Cuba | Americas | Latin America and the Caribbean |
| **SRR5633354** | 14/03/2017 | 1.1.2.15.2929.3683.7077 | Cuba | Americas | Latin America and the Caribbean |
| **SRR6919160** | 20/03/2017 | 1.1.2.15.2929.3683.7099 | Cuba | Americas | Latin America and the Caribbean |
| **SRR6920459** | 19/06/2017 | 1.1.2.15.3069.3898.7617 | Cuba | Americas | Latin America and the Caribbean |
| **SRR7842558** | 23/07/2018 | 1.1.2.15.3111.3985.10500 | Cuba | Americas | Latin America and the Caribbean |
| **SRR6919512** | 22/09/2017 | 1.1.2.15.3111.4133.8467 | Cuba | Americas | Latin America and the Caribbean |
| **SRR6922496** | 05/10/2017 | 1.1.2.15.3218.4179.8595 | Cuba | Americas | Latin America and the Caribbean |

| | | | | | |
|---|---|---|---|---|---|
| **SRR1965454** | 16/09/2014 | 1.1.2.15.361.1317.1778 | Cuba | Americas | Latin America and the Caribbean |
| **SRR7298362** | 30/05/2018 | 1.1.2.15.361.1950.10060 | Cuba | Americas | Latin America and the Caribbean |
| **SRR7998237** | 20/06/2018 | 1.1.2.15.361.1950.10160 | Cuba | Americas | Latin America and the Caribbean |
| **SRR7842568** | 22/08/2018 | 1.1.2.15.361.1950.11481 | Cuba | Americas | Latin America and the Caribbean |
| **SRR7841552** | 30/08/2018 | 1.1.2.15.361.1950.11571 | Cuba | Americas | Latin America and the Caribbean |
| **SRR8293824** | 06/09/2018 | 1.1.2.15.361.1950.11642 | Cuba | Americas | Latin America and the Caribbean |
| **SRR7885130** | 10/09/2018 | 1.1.2.15.361.1950.11658 | Cuba | Americas | Latin America and the Caribbean |
| **SRR7890428** | 05/09/2018 | 1.1.2.15.361.1950.11673 | Cuba | Americas | Latin America and the Caribbean |
| **SRR8427182** | 18/12/2018 | 1.1.2.15.361.1950.12578 | Cuba | Americas | Latin America and the Caribbean |
| **SRR8731039** | 12/05/2015 | 1.1.2.15.361.1950.2803 | Cuba | Americas | Latin America and the Caribbean |
| **SRR6900953** | 18/05/2017 | 1.1.2.15.361.1950.3179 | Cuba | Americas | Latin America and the Caribbean |
| **SRR7506784** | 11/09/2015 | 1.1.2.15.361.1950.3456 | Cuba | Americas | Latin America and the Caribbean |
| **SRR5194299** | 11/05/2016 | 1.1.2.15.361.1950.4656 | Cuba | Americas | Latin America and the Caribbean |
| **SRR5193167** | 15/06/2016 | 1.1.2.15.361.1950.4836 | Cuba | Americas | Latin America and the Caribbean |
| **SRR6900454** | 22/06/2016 | 1.1.2.15.361.1950.4867 | Cuba | Americas | Latin America and the Caribbean |
| **SRR5193749** | 21/07/2016 | 1.1.2.15.361.1950.5015 | Cuba | Americas | Latin America and the Caribbean |
| **SRR5193061** | 15/11/2016 | 1.1.2.15.361.1950.6275 | Cuba | Americas | Latin America and the Caribbean |
| **SRR6922611** | 28/11/2016 | 1.1.2.15.361.1950.6320 | Cuba | Americas | Latin America and the Caribbean |
| **SRR6900542** | 29/11/2016 | 1.1.2.15.361.1950.6327 | Cuba | Americas | Latin America and the Caribbean |
| **SRR7349226** | 13/12/2016 | 1.1.2.15.361.1950.6439 | Cuba | Americas | Latin America and the Caribbean |
| **SRR5585296** | 05/01/2017 | 1.1.2.15.361.1950.6832 | Cuba | Americas | Latin America and the Caribbean |
| **SRR6918566** | 26/01/2017 | 1.1.2.15.361.1950.6901 | Cuba | Americas | Latin America and the Caribbean |
| **SRR6922672** | 07/02/2017 | 1.1.2.15.361.1950.6926 | Cuba | Americas | Latin America and the Caribbean |
| **SRR6919260** | 23/02/2017 | 1.1.2.15.361.1950.7011 | Cuba | Americas | Latin America and the Caribbean |
| **SRR6897976** | 06/03/2017 | 1.1.2.15.361.1950.7045 | Cuba | Americas | Latin America and the Caribbean |
| **SRR5632027** | 08/03/2017 | 1.1.2.15.361.1950.7052 | Cuba | Americas | Latin America and the Caribbean |
| **SRR6922122** | 04/04/2017 | 1.1.2.15.361.1950.7133 | Cuba | Americas | Latin America and the Caribbean |
| **SRR6922016** | 05/04/2017 | 1.1.2.15.361.1950.7144 | Cuba | Americas | Latin America and the Caribbean |
| **SRR5584636** | 10/04/2017 | 1.1.2.15.361.1950.7158 | Cuba | Americas | Latin America and the Caribbean |
| **SRR5584554** | 12/04/2017 | 1.1.2.15.361.1950.7179 | Cuba | Americas | Latin America and the Caribbean |
| **SRR5585015** | 04/05/2017 | 1.1.2.15.361.1950.7238 | Cuba | Americas | Latin America and the Caribbean |
| **SRR6922487** | 04/05/2017 | 1.1.2.15.361.1950.7256 | Cuba | Americas | Latin America and the Caribbean |
| **SRR6922671** | 19/05/2017 | 1.1.2.15.361.1950.7319 | Cuba | Americas | Latin America and the Caribbean |
| **SRR6919030** | 04/07/2017 | 1.1.2.15.361.1950.7718 | Cuba | Americas | Latin America and the Caribbean |
| **SRR6900197** | 05/07/2017 | 1.1.2.15.361.1950.7720 | Cuba | Americas | Latin America and the Caribbean |
| **SRR6896965** | 12/07/2017 | 1.1.2.15.361.1950.7777 | Cuba | Americas | Latin America and the Caribbean |

| | | | | | |
|---|---|---|---|---|---|
| **SRR6920148** | 02/08/2017 | 1.1.2.15.361.1950.7978 | Cuba | Americas | Latin America and the Caribbean |
| **SRR6900373** | 11/09/2017 | 1.1.2.15.361.1950.8293 | Cuba | Americas | Latin America and the Caribbean |
| **SRR6898010** | 12/09/2017 | 1.1.2.15.361.1950.8350 | Cuba | Americas | Latin America and the Caribbean |
| **SRR6919920** | 12/09/2017 | 1.1.2.15.361.1950.8361 | Cuba | Americas | Latin America and the Caribbean |
| **SRR6898456** | 21/09/2017 | 1.1.2.15.361.1950.8466 | Cuba | Americas | Latin America and the Caribbean |
| **SRR8490746** | 23/07/2018 | 1.1.2.15.361.1950.8598 | Cuba | Americas | Latin America and the Caribbean |
| **SRR6897829** | 07/12/2017 | 1.1.2.15.361.1950.9001 | Cuba | Americas | Latin America and the Caribbean |
| **SRR7850599** | 12/07/2018 | 1.1.2.15.361.1950.9334 | Cuba | Americas | Latin America and the Caribbean |
| **SRR8508660** | 26/02/2018 | 1.1.2.15.361.1950.9558 | Cuba | Americas | Latin America and the Caribbean |
| **SRR8509031** | 27/02/2018 | 1.1.2.15.361.1950.9571 | Cuba | Americas | Latin America and the Caribbean |
| **SRR7188019** | 06/04/2018 | 1.1.2.15.361.1950.9769 | Cuba | Americas | Latin America and the Caribbean |
| **SRR7223157** | 19/04/2018 | 1.1.2.15.361.1950.9841 | Cuba | Americas | Latin America and the Caribbean |
| **SRR7527873** | 24/04/2018 | 1.1.2.15.361.1950.9865 | Cuba | Americas | Latin America and the Caribbean |
| **SRR8524718** | 26/04/2018 | 1.1.2.15.361.1950.9879 | Cuba | Americas | Latin America and the Caribbean |
| **SRR7343911** | 02/05/2018 | 1.1.2.15.361.1950.9899 | Cuba | Americas | Latin America and the Caribbean |
| **SRR7209521** | 11/05/2018 | 1.1.2.15.361.1950.9942 | Cuba | Americas | Latin America and the Caribbean |
| **SRR5216159** | 08/02/2016 | 1.1.2.15.361.2574.4401 | Cuba | Americas | Latin America and the Caribbean |
| **SRR6900108** | 12/04/2017 | 1.1.2.15.361.2574.7184 | Cuba | Americas | Latin America and the Caribbean |
| **SRR6901154** | 23/02/2016 | 1.1.2.15.361.2592.4466 | Cuba | Americas | Latin America and the Caribbean |
| **SRR6900082** | 05/09/2016 | 1.1.2.15.361.2592.5495 | Cuba | Americas | Latin America and the Caribbean |
| **SRR5193610** | 26/10/2016 | 1.1.2.15.361.2592.6083 | Cuba | Americas | Latin America and the Caribbean |
| **SRR6922516** | 11/03/2016 | 1.1.2.15.361.2613.4519 | Cuba | Americas | Latin America and the Caribbean |
| **SRR5216103** | 19/04/2016 | 1.1.2.15.361.2648.4603 | Cuba | Americas | Latin America and the Caribbean |
| **SRR5216479** | 02/06/2016 | 1.1.2.15.361.2705.4758 | Cuba | Americas | Latin America and the Caribbean |
| **SRR5216603** | 02/09/2016 | 1.1.2.15.361.3055.5553 | Cuba | Americas | Latin America and the Caribbean |
| **SRR6898102** | 15/09/2016 | 1.1.2.15.361.3095.5688 | Cuba | Americas | Latin America and the Caribbean |
| **SRR5193585** | 04/10/2016 | 1.1.2.15.361.3146.5883 | Cuba | Americas | Latin America and the Caribbean |
| **SRR8364471** | 23/08/2018 | 1.1.2.15.361.3202.11506 | Cuba | Americas | Latin America and the Caribbean |
| **SRR5193753** | 19/10/2016 | 1.1.2.15.361.3202.6038 | Cuba | Americas | Latin America and the Caribbean |
| **SRR6924057** | 30/01/2017 | 1.1.2.15.361.3202.6903 | Cuba | Americas | Latin America and the Caribbean |
| **SRR5633213** | 23/03/2017 | 1.1.2.15.361.3692.7103 | Cuba | Americas | Latin America and the Caribbean |
| **SRR5632815** | 21/04/2017 | 1.1.2.15.361.3716.7175 | Cuba | Americas | Latin America and the Caribbean |
| **SRR7523078** | 07/03/2018 | 1.1.2.15.361.4254.9696 | Cuba | Americas | Latin America and the Caribbean |
| **SRR6898378** | 15/11/2017 | 1.1.2.15.361.4261.8875 | Cuba | Americas | Latin America and the Caribbean |
| **SRR7368935** | 15/01/2018 | 1.1.2.15.361.4411.9230 | Cuba | Americas | Latin America and the Caribbean |
| **SRR8568766** | 31/01/2019 | 1.1.2.15.361.4646.12705 | Cuba | Americas | Latin America and the Caribbean |

| | | | | | |
|---|---|---|---|---|---|
| **SRR8492344** | 12/06/2018 | 1.1.2.15.361.4673.9803 | Cuba | Americas | Latin America and the Caribbean |
| **SRR8503886** | 27/04/2018 | 1.1.2.15.361.4702.9872 | Cuba | Americas | Latin America and the Caribbean |
| **SRR8509189** | 17/05/2018 | 1.1.2.15.361.4729.9965 | Cuba | Americas | Latin America and the Caribbean |
| **SRR9287421** | 12/04/2019 | 1.1.2.15.361.4868.12973 | Cuba | Americas | Latin America and the Caribbean |
| **SRR7842654** | 25/07/2018 | 1.1.2.15.361.4882.10524 | Cuba | Americas | Latin America and the Caribbean |
| **SRR8201844** | 02/11/2018 | 1.1.2.15.361.4882.12370 | Cuba | Americas | Latin America and the Caribbean |
| **SRR8514513** | 15/01/2019 | 1.1.2.15.361.4882.12659 | Cuba | Americas | Latin America and the Caribbean |
| **SRR7828238** | 21/08/2018 | 1.1.2.15.361.5354.11435 | Cuba | Americas | Latin America and the Caribbean |
| **SRR7903106** | 29/08/2018 | 1.1.2.15.361.5389.11535 | Cuba | Americas | Latin America and the Caribbean |
| **SRR8297297** | 30/08/2018 | 1.1.2.15.361.5395.11563 | Cuba | Americas | Latin America and the Caribbean |
| **SRR7911491** | 10/09/2018 | 1.1.2.15.361.5427.11671 | Cuba | Americas | Latin America and the Caribbean |
| **SRR7911437** | 10/09/2018 | 1.1.2.15.361.5437.11716 | Cuba | Americas | Latin America and the Caribbean |
| **SRR8084289** | 05/10/2018 | 1.1.2.15.361.5639.12109 | Cuba | Americas | Latin America and the Caribbean |
| **SRR8585128** | 05/02/2019 | 1.1.2.15.361.5809.12716 | Cuba | Americas | Latin America and the Caribbean |
| **SRR8703575** | 19/02/2019 | 1.1.2.15.361.5827.12761 | Cuba | Americas | Latin America and the Caribbean |
| **SRR8724658** | 26/02/2019 | 1.1.2.15.361.5843.12800 | Cuba | Americas | Latin America and the Caribbean |
| **SRR7458813** | 03/06/2015 | 1.1.2.15.383.1245.2883 | Cuba | Americas | Latin America and the Caribbean |
| **SRR5220517** | 11/09/2015 | 1.1.2.15.383.1245.4294 | Cuba | Americas | Latin America and the Caribbean |
| **SRR6919819** | 28/11/2017 | 1.1.2.15.383.1245.8847 | Cuba | Americas | Latin America and the Caribbean |
| **SRR6924069** | 15/06/2017 | 1.1.2.15.383.3918.7683 | Cuba | Americas | Latin America and the Caribbean |
| **SRR6900152** | 22/08/2017 | 1.1.2.15.383.4049.8169 | Cuba | Americas | Latin America and the Caribbean |
| **SRR7842674** | 04/09/2018 | 1.1.2.15.3870.5409.11611 | Cuba | Americas | Latin America and the Caribbean |
| **SRR8272603** | 10/09/2018 | 1.1.2.15.3880.5426.11666 | Cuba | Americas | Latin America and the Caribbean |
| **SRR8293602** | 28/11/2018 | 1.1.2.15.3880.5723.12493 | Cuba | Americas | Latin America and the Caribbean |
| **SRR1968811** | 01/05/2014 | 1.1.2.15.428.438.583 | Cuba | Americas | Latin America and the Caribbean |
| **SRR1962045** | 13/11/2014 | 1.1.2.15.428.709.2062 | Cuba | Americas | Latin America and the Caribbean |
| **SRR5220965** | 25/06/2015 | 1.1.2.15.729.2010.2959 | Cuba | Americas | Latin America and the Caribbean |
| **SRR5193683** | 19/07/2016 | 1.1.2.15.729.2010.5001 | Cuba | Americas | Latin America and the Caribbean |
| **SRR5193830** | 27/07/2016 | 1.1.2.15.729.2010.5049 | Cuba | Americas | Latin America and the Caribbean |
| **SRR5193950** | 29/12/2016 | 1.1.2.15.729.2010.6790 | Cuba | Americas | Latin America and the Caribbean |
| **SRR7903100** | 23/12/2015 | 1.1.2.15.729.2508.4236 | Cuba | Americas | Latin America and the Caribbean |
| **SRR1963097** | 02/12/2014 | 1.1.2.15.729.945.1213 | Cuba | Americas | Latin America and the Caribbean |
| **SRR6897790** | 31/05/2017 | 1.1.333.103.103.548.5035 | Cuba | Americas | Latin America and the Caribbean |
| **SRR3286681** | 05/05/2015 | 1.5.79.1029.1675.1939.2774 | Cuba | Americas | Latin America and the Caribbean |
| **SRR7172561** | 25/11/2015 | 1.1.2.1326.1918.2290.4088 | Cyprus | Asia | Western Asia |
| **SRR5193792** | 01/09/2016 | 1.1.2.1326.544.571.5369 | Cyprus | Asia | Western Asia |
| **SRR7842623** | 30/08/2018 | 1.1.2.1326.800.2147.11576 | Cyprus | Asia | Western Asia |

| | | | | | |
|---|---|---|---|---|---|
| **SRR8201827** | 06/11/2018 | 1.1.2.1326.800.2147.12382 | Cyprus | Asia | Western Asia |
| **SRR7350649** | 18/04/2018 | 1.1.2.1326.800.2147.9832 | Cyprus | Asia | Western Asia |
| **SRR5193714** | 18/10/2016 | 1.1.2.1326.800.3205.6044 | Cyprus | Asia | Western Asia |
| **SRR7297901** | 05/01/2018 | 1.1.2.1326.800.3205.9204 | Cyprus | Asia | Western Asia |
| **SRR7892273** | 10/05/2018 | 1.1.2.1326.800.3205.9935 | Cyprus | Asia | Western Asia |
| **SRR5216509** | 02/12/2015 | 1.1.2.1712.475.492.4084 | Cyprus | Asia | Western Asia |
| **SRR8281176** | 21/11/2018 | 1.1.2.237.3136.4038.11530 | Cyprus | Asia | Western Asia |
| **SRR8116959** | 16/10/2018 | 1.1.2.237.3136.4038.12192 | Cyprus | Asia | Western Asia |
| **SRR8182955** | 23/10/2018 | 1.1.2.2533.3617.4833.12299 | Cyprus | Asia | Western Asia |
| **SRR6898108** | 17/08/2017 | 1.1.2.28.195.195.8125 | Cyprus | Asia | Western Asia |
| **SRR8281265** | 17/10/2018 | 1.1.2.28.43.1319.12219 | Cyprus | Asia | Western Asia |
| **SRR5220283** | 03/10/2016 | 1.1.2.314.476.3137.5843 | Cyprus | Asia | Western Asia |
| **SRR5215526** | 25/08/2015 | 1.11.48.1103.1829.2168.3382 | Cyprus | Asia | Western Asia |
| **SRR5220909** | 22/10/2015 | 1.2.3.151.151.783.2809 | Cyprus | Asia | Western Asia |
| **SRR5193235** | 10/10/2016 | 1.2.3.151.362.363.5918 | Cyprus | Asia | Western Asia |
| **SRR7523809** | 19/06/2018 | 1.2.3.18.175.175.10143 | Cyprus | Asia | Western Asia |
| **SRR6921965** | 01/11/2016 | 1.2.3.18.175.175.3397 | Cyprus | Asia | Western Asia |
| **SRR8293559** | 27/11/2018 | 1.2.3.18.1895.2261.12482 | Cyprus | Asia | Western Asia |
| **SRR5193415** | 28/07/2016 | 1.2.3.18.62.62.5044 | Cyprus | Asia | Western Asia |
| **SRR5194027** | 29/11/2016 | 59.106.504.1891.2617.3299.6356 | Cyprus | Asia | Western Asia |
| **SRR7873970** | 22/08/2018 | 59.106.504.2552.3858.5371.11485 | Cyprus | Asia | Western Asia |
| **SRR1966691** | 04/08/2014 | 1.1.2.95.95.95.95 | Czech republic | Europe | Eastern Europe |
| **SRR1967811** | 05/03/2015 | 1.2.3.18.1005.1128.1502 | Czech republic | Europe | Eastern Europe |
| **SRR6920125** | 21/09/2016 | 1.2.3.18.180.180.11135 | Czech republic | Europe | Eastern Europe |
| **SRR7873898** | 07/09/2018 | 1.2.3.18.180.180.11644 | Czech republic | Europe | Eastern Europe |
| **SRR6919318** | 12/09/2017 | 1.2.3.18.180.5089.10806 | Czech republic | Europe | Eastern Europe |
| **SRR1966789** | 06/02/2015 | 1.2.3.18.180.5185.11022 | Czech republic | Europe | Eastern Europe |
| **SRR5193440** | 14/09/2016 | 1.2.3.18.2226.2749.5593 | Czech republic | Europe | Eastern Europe |
| **SRR7538806** | 05/07/2018 | 1.2.3.18.3616.4832.10337 | Czech republic | Europe | Eastern Europe |
| **SRR6899356** | 18/12/2017 | 1.2.3.18.377.4326.9039 | Czech republic | Europe | Eastern Europe |
| **SRR1965484** | 04/09/2014 | 1.2.3.18.38.38.458 | Czech republic | Europe | Eastern Europe |
| **SRR6919090** | 25/09/2017 | 1.1.2.61.3197.4141.8483 | Dominica | Americas | Latin America and the Caribbean |
| **SRR6899429** | 26/04/2016 | 1.1.2.61.61.2651.4616 | Dominica | Americas | Latin America and the Caribbean |
| **SRR6919834** | 07/08/2017 | 1.1.2.61.61.3974.7910 | Dominica | Americas | Latin America and the Caribbean |
| **SRR6919478** | 23/10/2017 | 1.1.2.61.61.4202.8722 | Dominica | Americas | Latin America and the Caribbean |
| **SRR8492331** | 14/06/2018 | 1.1.2.61.61.4693.10126 | Dominica | Americas | Latin America and the Caribbean |
| **SRR5193404** | 08/04/2016 | 1.1.2.61.61.561.4561 | Dominica | Americas | Latin America and the Caribbean |
| **SRR5220052** | 10/01/2017 | 1.1.2.61.61.561.4987 | Dominica | Americas | Latin America and the Caribbean |
| **SRR5193594** | 06/01/2017 | 1.1.2.61.61.561.6833 | Dominica | Americas | Latin America and the Caribbean |
| **SRR6900482** | 20/04/2017 | 1.1.2.61.61.561.7188 | Dominica | Americas | Latin America and the Caribbean |
| **SRR6900654** | 25/10/2017 | 1.1.2.61.61.561.8739 | Dominica | Americas | Latin America and the |

| | | | | | |
|---|---|---|---|---|---|
| | | | | | Caribbean |
| **SRR6922712** | 10/03/2017 | 1.1.2.61.61.61.7060 | Dominica | Americas | Latin America and the Caribbean |
| **SRR7417201** | 09/02/2018 | 1.1.2.61.61.61.9376 | Dominica | Americas | Latin America and the Caribbean |
| **SRR7343933** | 09/02/2018 | 1.1.2.61.61.61.9455 | Dominica | Americas | Latin America and the Caribbean |
| **SRR3049056** | 10/06/2014 | 1.1.2.1326.544.571.2083 | Dominican republic | Americas | Latin America and the Caribbean |
| **SRR6924131** | 23/03/2017 | 1.1.2.61.2935.3693.7105 | Dominican republic | Americas | Latin America and the Caribbean |
| **SRR8740776** | 18/06/2015 | 1.1.2.61.61.2011.2970 | Dominican republic | Americas | Latin America and the Caribbean |
| **SRR6919481** | 05/05/2016 | 1.1.2.61.61.2651.4616 | Dominican republic | Americas | Latin America and the Caribbean |
| **SRR5220528** | 23/05/2016 | 1.1.2.61.61.2651.4697 | Dominican republic | Americas | Latin America and the Caribbean |
| **SRR6919314** | 04/09/2017 | 1.1.2.61.61.4062.8263 | Dominican republic | Americas | Latin America and the Caribbean |
| **SRR6922519** | 17/10/2017 | 1.1.2.61.61.4062.8667 | Dominican republic | Americas | Latin America and the Caribbean |
| **SRR6901170** | 07/11/2017 | 1.1.2.61.61.4062.8810 | Dominican republic | Americas | Latin America and the Caribbean |
| **SRR6896967** | 07/11/2017 | 1.1.2.61.61.4062.8816 | Dominican republic | Americas | Latin America and the Caribbean |
| **SRR7351490** | 12/02/2018 | 1.1.2.61.61.4062.9457 | Dominican republic | Americas | Latin America and the Caribbean |
| **SRR6898884** | 18/10/2017 | 1.1.2.61.61.4202.8694 | Dominican republic | Americas | Latin America and the Caribbean |
| **SRR6920002** | 31/10/2017 | 1.1.2.61.61.4202.8722 | Dominican republic | Americas | Latin America and the Caribbean |
| **SRR6920150** | 01/11/2017 | 1.1.2.61.61.4202.8777 | Dominican republic | Americas | Latin America and the Caribbean |
| **SRR6924052** | 23/11/2017 | 1.1.2.61.61.4265.8890 | Dominican republic | Americas | Latin America and the Caribbean |
| **SRR6898439** | 04/12/2017 | 1.1.2.61.61.4265.8944 | Dominican republic | Americas | Latin America and the Caribbean |
| **SRR6898417** | 29/11/2017 | 1.1.2.61.61.4287.8943 | Dominican republic | Americas | Latin America and the Caribbean |
| **SRR7292896** | 10/01/2018 | 1.1.2.61.61.4398.9192 | Dominican republic | Americas | Latin America and the Caribbean |
| **SRR7439596** | 01/06/2018 | 1.1.2.61.61.4676.10059 | Dominican republic | Americas | Latin America and the Caribbean |
| **SRR7842415** | 17/07/2018 | 1.1.2.61.61.4676.10452 | Dominican republic | Americas | Latin America and the Caribbean |
| **SRR8499082** | 12/06/2018 | 1.1.2.61.61.4693.10106 | Dominican republic | Americas | Latin America and the Caribbean |
| **SRR7450823** | 20/06/2018 | 1.1.2.61.61.4693.10159 | Dominican republic | Americas | Latin America and the Caribbean |
| **SRR7850626** | 08/08/2018 | 1.1.2.61.61.4693.11366 | Dominican republic | Americas | Latin America and the Caribbean |
| **SRR8503968** | 24/04/2018 | 1.1.2.61.61.4693.9854 | Dominican republic | Americas | Latin America and the Caribbean |
| **SRR7842634** | 02/08/2018 | 1.1.2.61.61.4827.11353 | Dominican republic | Americas | Latin America and the Caribbean |
| **SRR8142752** | 18/10/2018 | 1.1.2.61.61.4827.12221 | Dominican republic | Americas | Latin America and the Caribbean |
| **SRR8380061** | 23/07/2018 | 1.1.2.61.61.4849.10425 | Dominican republic | Americas | Latin America and the Caribbean |
| **SRR7962209** | 18/09/2018 | 1.1.2.61.61.5384.11795 | Dominican republic | Americas | Latin America and the Caribbean |
| **SRR8325500** | 02/10/2018 | 1.1.2.61.61.5384.12065 | Dominican republic | Americas | Latin America and the Caribbean |
| **SRR7469074** | 21/06/2018 | 1.1.2.61.61.561.10188 | Dominican republic | Americas | Latin America and the Caribbean |
| **SRR7885126** | 10/07/2018 | 1.1.2.61.61.561.10402 | Dominican republic | Americas | Latin America and the Caribbean |
| **SRR8350280** | 21/08/2018 | 1.1.2.61.61.561.11444 | Dominican republic | Americas | Latin America and the Caribbean |
| **SRR8204753** | 18/10/2018 | 1.1.2.61.61.561.12222 | Dominican republic | Americas | Latin America and the Caribbean |

| | | | | | |
|---|---|---|---|---|---|
| **SRR3049358** | 22/12/2014 | 1.1.2.61.61.561.1567 | Dominican republic | Americas | Latin America and the Caribbean |
| **SRR8730397** | 17/06/2015 | 1.1.2.61.61.561.2839 | Dominican republic | Americas | Latin America and the Caribbean |
| **SRR7284514** | 04/06/2015 | 1.1.2.61.61.561.2881 | Dominican republic | Americas | Latin America and the Caribbean |
| **SRR5216581** | 22/06/2015 | 1.1.2.61.61.561.3008 | Dominican republic | Americas | Latin America and the Caribbean |
| **SRR5215841** | 02/07/2015 | 1.1.2.61.61.561.3048 | Dominican republic | Americas | Latin America and the Caribbean |
| **SRR5194143** | 09/07/2015 | 1.1.2.61.61.561.3092 | Dominican republic | Americas | Latin America and the Caribbean |
| **SRR3284835** | 11/08/2015 | 1.1.2.61.61.561.3302 | Dominican republic | Americas | Latin America and the Caribbean |
| **SRR7286867** | 09/09/2015 | 1.1.2.61.61.561.3448 | Dominican republic | Americas | Latin America and the Caribbean |
| **SRR5220877** | 09/10/2015 | 1.1.2.61.61.561.3740 | Dominican republic | Americas | Latin America and the Caribbean |
| **SRR7443897** | 14/10/2015 | 1.1.2.61.61.561.3884 | Dominican republic | Americas | Latin America and the Caribbean |
| **SRR7417302** | 27/11/2015 | 1.1.2.61.61.561.4105 | Dominican republic | Americas | Latin America and the Caribbean |
| **SRR8704751** | 04/12/2015 | 1.1.2.61.61.561.4172 | Dominican republic | Americas | Latin America and the Caribbean |
| **SRR5193398** | 15/12/2015 | 1.1.2.61.61.561.4214 | Dominican republic | Americas | Latin America and the Caribbean |
| **SRR5216237** | 02/02/2016 | 1.1.2.61.61.561.4393 | Dominican republic | Americas | Latin America and the Caribbean |
| **SRR5216577** | 16/09/2016 | 1.1.2.61.61.561.4856 | Dominican republic | Americas | Latin America and the Caribbean |
| **SRR6918334** | 21/10/2016 | 1.1.2.61.61.561.6057 | Dominican republic | Americas | Latin America and the Caribbean |
| **SRR5194328** | 24/10/2016 | 1.1.2.61.61.561.6118 | Dominican republic | Americas | Latin America and the Caribbean |
| **SRR6922492** | 30/01/2017 | 1.1.2.61.61.561.6905 | Dominican republic | Americas | Latin America and the Caribbean |
| **SRR6919178** | 04/10/2017 | 1.1.2.61.61.561.7188 | Dominican republic | Americas | Latin America and the Caribbean |
| **SRR5633116** | 25/04/2017 | 1.1.2.61.61.561.7209 | Dominican republic | Americas | Latin America and the Caribbean |
| **SRR6918586** | 12/05/2017 | 1.1.2.61.61.561.7279 | Dominican republic | Americas | Latin America and the Caribbean |
| **SRR6922651** | 19/06/2017 | 1.1.2.61.61.561.7628 | Dominican republic | Americas | Latin America and the Caribbean |
| **SRR6900192** | 31/10/2017 | 1.1.2.61.61.561.7714 | Dominican republic | Americas | Latin America and the Caribbean |
| **SRR8166212** | 22/10/2018 | 1.1.2.61.61.5674.12262 | Dominican republic | Americas | Latin America and the Caribbean |
| **SRR8258163** | 13/11/2018 | 1.1.2.61.61.5708.12440 | Dominican republic | Americas | Latin America and the Caribbean |
| **SRR8492423** | 18/06/2018 | 1.1.2.61.61.61.10190 | Dominican republic | Americas | Latin America and the Caribbean |
| **SRR7867159** | 13/07/2018 | 1.1.2.61.61.61.10424 | Dominican republic | Americas | Latin America and the Caribbean |
| **SRR7873873** | 17/07/2018 | 1.1.2.61.61.61.10501 | Dominican republic | Americas | Latin America and the Caribbean |
| **SRR7828267** | 16/08/2018 | 1.1.2.61.61.61.11432 | Dominican republic | Americas | Latin America and the Caribbean |
| **SRR8258143** | 13/11/2018 | 1.1.2.61.61.61.12424 | Dominican republic | Americas | Latin America and the Caribbean |
| **SRR3284817** | 10/06/2015 | 1.1.2.61.61.61.2919 | Dominican republic | Americas | Latin America and the Caribbean |
| **SRR5193062** | 30/09/2015 | 1.1.2.61.61.61.3663 | Dominican republic | Americas | Latin America and the Caribbean |
| **SRR7465079** | 29/09/2015 | 1.1.2.61.61.61.3680 | Dominican republic | Americas | Latin America and the Caribbean |
| **SRR5216056** | 02/12/2015 | 1.1.2.61.61.61.4111 | Dominican republic | Americas | Latin America and the Caribbean |
| **SRR5193239** | 30/12/2015 | 1.1.2.61.61.61.4242 | Dominican republic | Americas | Latin America and the Caribbean |
| **SRR8701072** | 08/02/2016 | 1.1.2.61.61.61.4410 | Dominican republic | Americas | Latin America and the Caribbean |

| | | | | | |
|---|---|---|---|---|---|
| **SRR6901174** | 22/03/2016 | 1.1.2.61.61.61.4532 | Dominican republic | Americas | Latin America and the Caribbean |
| **SRR5194246** | 30/03/2016 | 1.1.2.61.61.61.4539 | Dominican republic | Americas | Latin America and the Caribbean |
| **SRR5216551** | 07/04/2016 | 1.1.2.61.61.61.4567 | Dominican republic | Americas | Latin America and the Caribbean |
| **SRR1969189** | 08/07/2014 | 1.1.2.61.61.61.61 | Dominican republic | Americas | Latin America and the Caribbean |
| **SRR6896814** | 05/07/2017 | 1.1.2.61.61.61.7060 | Dominican republic | Americas | Latin America and the Caribbean |
| **SRR5583245** | 05/04/2017 | 1.1.2.61.61.61.7135 | Dominican republic | Americas | Latin America and the Caribbean |
| **SRR6899411** | 03/07/2017 | 1.1.2.61.61.61.7699 | Dominican republic | Americas | Latin America and the Caribbean |
| **SRR6919286** | 04/07/2017 | 1.1.2.61.61.61.7723 | Dominican republic | Americas | Latin America and the Caribbean |
| **SRR6898455** | 05/07/2017 | 1.1.2.61.61.61.7724 | Dominican republic | Americas | Latin America and the Caribbean |
| **SRR6898386** | 24/07/2017 | 1.1.2.61.61.61.7860 | Dominican republic | Americas | Latin America and the Caribbean |
| **SRR6921911** | 02/08/2017 | 1.1.2.61.61.61.7871 | Dominican republic | Americas | Latin America and the Caribbean |
| **SRR6900179** | 02/08/2017 | 1.1.2.61.61.61.7881 | Dominican republic | Americas | Latin America and the Caribbean |
| **SRR7426962** | 01/08/2017 | 1.1.2.61.61.61.7989 | Dominican republic | Americas | Latin America and the Caribbean |
| **SRR6922686** | 22/08/2017 | 1.1.2.61.61.61.8188 | Dominican republic | Americas | Latin America and the Caribbean |
| **SRR6920147** | 29/08/2017 | 1.1.2.61.61.61.8240 | Dominican republic | Americas | Latin America and the Caribbean |
| **SRR6900156** | 30/08/2017 | 1.1.2.61.61.61.8290 | Dominican republic | Americas | Latin America and the Caribbean |
| **SRR6922683** | 12/10/2017 | 1.1.2.61.61.61.8643 | Dominican republic | Americas | Latin America and the Caribbean |
| **SRR6897948** | 18/10/2017 | 1.1.2.61.61.61.8691 | Dominican republic | Americas | Latin America and the Caribbean |
| **SRR6919515** | 04/12/2017 | 1.1.2.61.61.61.8970 | Dominican republic | Americas | Latin America and the Caribbean |
| **SRR6919769** | 29/08/2017 | 1.1.2.61.61.61.9649 | Dominican republic | Americas | Latin America and the Caribbean |
| **SRR7533446** | 15/05/2018 | 1.1.2.61.61.61.9959 | Dominican republic | Americas | Latin America and the Caribbean |
| **SRR1958103** | 10/10/2014 | 1.1.2.61.61.729.905 | Dominican republic | Americas | Latin America and the Caribbean |
| **SRR3585348** | 25/08/2015 | 1.1.256.1101.1827.2165.3375 | Dominican republic | Americas | Latin America and the Caribbean |
| **SRR8711366** | 17/09/2015 | 1.1.256.1101.1827.2165.3530 | Dominican republic | Americas | Latin America and the Caribbean |
| **SRR8704615** | 06/11/2015 | 1.1.256.1101.1827.2165.3980 | Dominican republic | Americas | Latin America and the Caribbean |
| **SRR5193948** | 09/06/2016 | 1.1.256.1101.1827.2165.4781 | Dominican republic | Americas | Latin America and the Caribbean |
| **SRR7475501** | 04/09/2015 | 1.1.256.1101.1827.2516.4250 | Dominican republic | Americas | Latin America and the Caribbean |
| **SRR6918592** | 20/12/2017 | 1.1.256.1101.1827.4290.9050 | Dominican republic | Americas | Latin America and the Caribbean |
| **SRR7350732** | 16/11/2015 | 1.5.11.13.2030.2451.4068 | Dominican republic | Americas | Latin America and the Caribbean |
| **SRR7416263** | 29/01/2018 | 1.5.11.13.3364.4438.9310 | Dominican republic | Americas | Latin America and the Caribbean |
| **SRR5194051** | 24/07/2015 | 1.1.2.1074.1763.2074.3174 | Egypt | Africa | Northern Africa |
| **SRR5194069** | 26/08/2015 | 1.1.2.1100.1826.2164.3374 | Egypt | Africa | Northern Africa |
| **SRR5220411** | 01/10/2015 | 1.1.2.1100.1826.2164.3668 | Egypt | Africa | Northern Africa |
| **SRR1969115** | 30/09/2014 | 1.1.2.1326.544.571.2616 | Egypt | Africa | Northern Africa |
| **SRR8114907** | 04/10/2018 | 1.1.2.147.4014.5634.12083 | Egypt | Africa | Northern Africa |
| **SRR6922722** | 02/12/2016 | 1.1.2.1893.2623.3311.6382 | Egypt | Africa | Northern Africa |
| **SRR5216604** | 23/09/2015 | 1.1.2.2073.1887.2253.3599 | Egypt | Africa | Northern Africa |
| **SRR3285345** | 13/11/2015 | 1.1.2.2073.1887.2429.4019 | Egypt | Africa | Northern Africa |

| | | | | | |
|---|---|---|---|---|---|
| **SRR5215696** | 10/03/2016 | 1.1.2.2073.1887.2429.4499 | Egypt | Africa | Northern Africa |
| **SRR5193799** | 01/06/2016 | 1.1.2.2073.1887.2707.4761 | Egypt | Africa | Northern Africa |
| **SRR5216460** | 25/10/2016 | 1.1.2.2073.1887.3212.6091 | Egypt | Africa | Northern Africa |
| **SRR6901021** | 03/10/2017 | 1.1.2.2073.1887.4181.8600 | Egypt | Africa | Northern Africa |
| **SRR8377931** | 24/07/2018 | 1.1.2.2073.1887.4877.10512 | Egypt | Africa | Northern Africa |
| **SRR7439344** | 07/10/2015 | 1.1.2.2073.1933.2316.3755 | Egypt | Africa | Northern Africa |
| **SRR5216476** | 18/10/2016 | 1.1.2.2073.2557.3210.6074 | Egypt | Africa | Northern Africa |
| **SRR7892278** | 09/08/2017 | 1.1.2.2073.2557.3210.7914 | Egypt | Africa | Northern Africa |
| **SRR7842656** | 04/07/2018 | 1.1.2.2073.2557.4842.10379 | Egypt | Africa | Northern Africa |
| **SRR7873932** | 22/08/2018 | 1.1.2.2073.2557.5373.11491 | Egypt | Africa | Northern Africa |
| **SRR7997043** | 26/09/2018 | 1.1.2.2073.2557.5525.11899 | Egypt | Africa | Northern Africa |
| **SRR8172835** | 25/09/2018 | 1.1.2.2073.2557.5614.12017 | Egypt | Africa | Northern Africa |
| **SRR3286669** | 06/05/2015 | 1.1.2.2073.389.1688.2773 | Egypt | Africa | Northern Africa |
| **SRR3048808** | 03/06/2014 | 1.1.2.2073.389.1834.2560 | Egypt | Africa | Northern Africa |
| **SRR5193716** | 03/08/2015 | 1.1.2.2073.389.2101.3242 | Egypt | Africa | Northern Africa |
| **SRR3322381** | 17/09/2015 | 1.1.2.2073.389.2188.3440 | Egypt | Africa | Northern Africa |
| **SRR5193598** | 09/09/2015 | 1.1.2.2073.389.2224.3523 | Egypt | Africa | Northern Africa |
| **SRR3285350** | 30/09/2015 | 1.1.2.2073.389.2224.3655 | Egypt | Africa | Northern Africa |
| **SRR8704764** | 04/12/2015 | 1.1.2.2073.389.2224.4173 | Egypt | Africa | Northern Africa |
| **SRR6900954** | 25/05/2016 | 1.1.2.2073.389.2690.4708 | Egypt | Africa | Northern Africa |
| **SRR7879572** | 25/07/2018 | 1.1.2.2073.389.2854.10523 | Egypt | Africa | Northern Africa |
| **SRR8389679** | 14/12/2018 | 1.1.2.2073.389.2854.12576 | Egypt | Africa | Northern Africa |
| **SRR8479012** | 09/01/2019 | 1.1.2.2073.389.2854.12638 | Egypt | Africa | Northern Africa |
| **SRR8731027** | 28/05/2015 | 1.1.2.2073.389.2854.2854 | Egypt | Africa | Northern Africa |
| **SRR8724947** | 02/07/2015 | 1.1.2.2073.389.2854.3029 | Egypt | Africa | Northern Africa |
| **SRR5216497** | 23/09/2015 | 1.1.2.2073.389.2854.3614 | Egypt | Africa | Northern Africa |
| **SRR6918546** | 04/07/2017 | 1.1.2.2073.389.2854.7717 | Egypt | Africa | Northern Africa |
| **SRR6899347** | 28/07/2017 | 1.1.2.2073.389.2854.7909 | Egypt | Africa | Northern Africa |
| **SRR6897977** | 20/07/2017 | 1.1.2.2073.389.2854.7979 | Egypt | Africa | Northern Africa |
| **SRR6898832** | 17/08/2017 | 1.1.2.2073.389.2854.8132 | Egypt | Africa | Northern Africa |
| **SRR6899331** | 07/09/2017 | 1.1.2.2073.389.2854.8309 | Egypt | Africa | Northern Africa |
| **SRR6897877** | 26/09/2017 | 1.1.2.2073.389.2854.8495 | Egypt | Africa | Northern Africa |
| **SRR6897793** | 27/09/2017 | 1.1.2.2073.389.2854.8518 | Egypt | Africa | Northern Africa |
| **SRR6901067** | 10/10/2017 | 1.1.2.2073.389.2854.8656 | Egypt | Africa | Northern Africa |
| **SRR6900690** | 16/10/2017 | 1.1.2.2073.389.2854.8660 | Egypt | Africa | Northern Africa |
| **SRR6900651** | 07/11/2017 | 1.1.2.2073.389.2854.8812 | Egypt | Africa | Northern Africa |
| **SRR6922729** | 09/11/2017 | 1.1.2.2073.389.2854.8838 | Egypt | Africa | Northern Africa |
| **SRR6900270** | 13/11/2017 | 1.1.2.2073.389.2854.8860 | Egypt | Africa | Northern Africa |
| **SRR6899398** | 16/11/2017 | 1.1.2.2073.389.2854.8876 | Egypt | Africa | Northern Africa |
| **SRR6898851** | 15/11/2017 | 1.1.2.2073.389.2854.8880 | Egypt | Africa | Northern Africa |
| **SRR6921922** | 28/11/2017 | 1.1.2.2073.389.2854.8923 | Egypt | Africa | Northern Africa |
| **SRR6919299** | 29/11/2017 | 1.1.2.2073.389.2854.8939 | Egypt | Africa | Northern Africa |
| **SRR6920503** | 01/12/2017 | 1.1.2.2073.389.2854.8962 | Egypt | Africa | Northern Africa |
| **SRR6900922** | 01/12/2017 | 1.1.2.2073.389.2854.8963 | Egypt | Africa | Northern Africa |
| **SRR7285634** | 16/01/2018 | 1.1.2.2073.389.2854.9259 | Egypt | Africa | Northern Africa |
| **SRR8524434** | 16/01/2018 | 1.1.2.2073.389.2854.9260 | Egypt | Africa | Northern Africa |
| **SRR5220678** | 29/06/2016 | 1.1.2.2073.389.393.4897 | Egypt | Africa | Northern Africa |
| **SRR7209569** | 10/04/2018 | 1.1.2.2073.389.4677.9811 | Egypt | Africa | Northern Africa |

| | | | | | |
|---|---|---|---|---|---|
| **SRR7439423** | 13/04/2018 | 1.1.2.2073.389.4683.9819 | Egypt | Africa | Northern Africa |
| **SRR1963485** | 14/01/2015 | 1.1.2.2073.389.551.640 | Egypt | Africa | Northern Africa |
| **SRR1969451** | 14/11/2014 | 1.1.2.2073.624.667.813 | Egypt | Africa | Northern Africa |
| **SRR3048719** | 03/10/2014 | 1.1.2.2073.776.851.1082 | Egypt | Africa | Northern Africa |
| **SRR1970042** | 06/10/2014 | 1.1.2.2073.776.851.1224 | Egypt | Africa | Northern Africa |
| **SRR1968869** | 09/09/2014 | 1.1.2.219.219.219.1041 | Egypt | Africa | Northern Africa |
| **SRR1965547** | 18/12/2014 | 1.1.2.219.219.219.1385 | Egypt | Africa | Northern Africa |
| **SRR1966193** | 05/11/2014 | 1.1.2.219.219.219.219 | Egypt | Africa | Northern Africa |
| **SRR8998262** | 17/04/2019 | 1.1.2.219.4123.5897.12982 | Egypt | Africa | Northern Africa |
| **SRR8438037** | 09/08/2018 | 1.1.2.2545.3839.5331.11386 | Egypt | Africa | Northern Africa |
| **SRR7873982** | 31/08/2018 | 1.1.2.2545.3839.5331.11571 | Egypt | Africa | Northern Africa |
| **SRR8204778** | 07/11/2018 | 1.1.2.2652.4037.5692.12383 | Egypt | Africa | Northern Africa |
| **SRR8936167** | 02/04/2019 | 1.1.2.2678.4117.5884.12931 | Egypt | Africa | Northern Africa |
| **SRR1968551** | 06/08/2014 | 1.1.2.273.1454.1676.2308 | Egypt | Africa | Northern Africa |
| **SRR1968653** | 18/08/2014 | 1.1.2.273.273.273.2454 | Egypt | Africa | Northern Africa |
| **SRR1969078** | 02/07/2014 | 1.1.2.273.273.273.2614 | Egypt | Africa | Northern Africa |
| **SRR1967821** | 07/05/2014 | 1.1.2.273.273.273.273 | Egypt | Africa | Northern Africa |
| **SRR5220556** | 29/04/2015 | 1.1.2.273.273.273.2756 | Egypt | Africa | Northern Africa |
| **SRR5193181** | 07/07/2015 | 1.1.2.273.273.273.3024 | Egypt | Africa | Northern Africa |
| **SRR5216189** | 10/07/2015 | 1.1.2.273.273.273.3072 | Egypt | Africa | Northern Africa |
| **SRR5193483** | 23/07/2015 | 1.1.2.273.273.273.3149 | Egypt | Africa | Northern Africa |
| **SRR3285474** | 06/08/2015 | 1.1.2.273.273.273.3262 | Egypt | Africa | Northern Africa |
| **SRR3048817** | 10/03/2015 | 1.1.2.273.602.798.1403 | Egypt | Africa | Northern Africa |
| **SRR1963118** | 22/12/2014 | 1.1.2.273.602.798.1752 | Egypt | Africa | Northern Africa |
| **SRR5216290** | 26/08/2015 | 1.1.2.273.602.798.3366 | Egypt | Africa | Northern Africa |
| **SRR1966488** | 05/11/2014 | 1.1.2.273.602.798.996 | Egypt | Africa | Northern Africa |
| **SRR1957904** | 31/10/2014 | 1.1.2.273.602.820.1033 | Egypt | Africa | Northern Africa |
| **SRR6919255** | 20/06/2017 | 1.1.2.274.274.3900.7622 | Egypt | Africa | Northern Africa |
| **SRR8873669** | 26/03/2019 | 1.1.2.28.149.149.12887 | Egypt | Africa | Northern Africa |
| **SRR3286855** | 17/06/2015 | 1.1.2.28.149.149.149 | Egypt | Africa | Northern Africa |
| **SRR3284740** | 21/04/2015 | 1.1.2.28.1666.1924.2739 | Egypt | Africa | Northern Africa |
| **SRR7842481** | 22/08/2018 | 1.1.2.28.2230.2759.11489 | Egypt | Africa | Northern Africa |
| **SRR7879324** | 04/09/2018 | 1.1.2.28.2230.2759.11620 | Egypt | Africa | Northern Africa |
| **SRR7890396** | 07/09/2018 | 1.1.2.28.2230.2759.11670 | Egypt | Africa | Northern Africa |
| **SRR8292220** | 27/11/2018 | 1.1.2.28.2230.2759.12488 | Egypt | Africa | Northern Africa |
| **SRR8648353** | 04/02/2019 | 1.1.2.28.2230.2759.12710 | Egypt | Africa | Northern Africa |
| **SRR5194022** | 28/06/2016 | 1.1.2.28.2230.2759.4901 | Egypt | Africa | Northern Africa |
| **SRR5194216** | 08/07/2016 | 1.1.2.28.2230.2759.4948 | Egypt | Africa | Northern Africa |
| **SRR5216483** | 21/07/2016 | 1.1.2.28.2230.2759.5014 | Egypt | Africa | Northern Africa |
| **SRR6919482** | 10/08/2016 | 1.1.2.28.2230.2759.5148 | Egypt | Africa | Northern Africa |
| **SRR5193939** | 07/11/2016 | 1.1.2.28.2230.2759.6231 | Egypt | Africa | Northern Africa |
| **SRR5194166** | 14/09/2016 | 1.1.2.28.2461.3067.5599 | Egypt | Africa | Northern Africa |
| **SRR5216044** | 03/11/2016 | 1.1.2.28.2596.3266.6247 | Egypt | Africa | Northern Africa |
| **SRR6898815** | 29/03/2017 | 1.1.2.28.2936.3694.7109 | Egypt | Africa | Northern Africa |
| **SRR8375229** | 19/07/2018 | 1.1.2.28.2953.3726.10463 | Egypt | Africa | Northern Africa |
| **SRR8370541** | 30/07/2018 | 1.1.2.28.2953.3726.10540 | Egypt | Africa | Northern Africa |
| **SRR8333794** | 14/08/2018 | 1.1.2.28.2953.3726.11404 | Egypt | Africa | Northern Africa |
| **SRR7867053** | 06/07/2017 | 1.1.2.28.2953.3726.7733 | Egypt | Africa | Northern Africa |

| | | | | | |
|---|---|---|---|---|---|
| **SRR8648291** | 11/08/2017 | 1.1.2.28.2972.3765.7932 | Egypt | Africa | Northern Africa |
| **SRR1967433** | 24/03/2015 | 1.1.2.28.355.355.356 | Egypt | Africa | Northern Africa |
| **SRR1967878** | 29/10/2014 | 1.1.2.28.413.1419.1924 | Egypt | Africa | Northern Africa |
| **SRR7475472** | 23/06/2015 | 1.1.2.28.413.474.519 | Egypt | Africa | Northern Africa |
| **SRR3049397** | 10/06/2014 | 1.1.2.28.415.1126.1500 | Egypt | Africa | Northern Africa |
| **SRR8704849** | 04/11/2015 | 1.1.2.28.415.1126.4017 | Egypt | Africa | Northern Africa |
| **SRR1970278** | 06/08/2014 | 1.1.2.28.415.1171.1563 | Egypt | Africa | Northern Africa |
| **SRR1957907** | 10/07/2014 | 1.1.2.28.415.1326.1787 | Egypt | Africa | Northern Africa |
| **SRR1959285** | 31/12/2014 | 1.1.2.28.415.422.1208 | Egypt | Africa | Northern Africa |
| **SRR1957993** | 03/09/2014 | 1.1.2.28.415.422.1356 | Egypt | Africa | Northern Africa |
| **SRR1968658** | 19/08/2014 | 1.1.2.28.415.422.1806 | Egypt | Africa | Northern Africa |
| **SRR7465131** | 30/05/2018 | 1.1.2.28.43.1319.10043 | Egypt | Africa | Northern Africa |
| **SRR7457147** | 30/05/2018 | 1.1.2.28.43.1319.10070 | Egypt | Africa | Northern Africa |
| **SRR7480125** | 25/06/2018 | 1.1.2.28.43.1319.10128 | Egypt | Africa | Northern Africa |
| **SRR7828410** | 18/07/2018 | 1.1.2.28.43.1319.10473 | Egypt | Africa | Northern Africa |
| **SRR7873801** | 23/07/2018 | 1.1.2.28.43.1319.10511 | Egypt | Africa | Northern Africa |
| **SRR7884499** | 10/09/2018 | 1.1.2.28.43.1319.11346 | Egypt | Africa | Northern Africa |
| **SRR8166254** | 12/09/2018 | 1.1.2.28.43.1319.11696 | Egypt | Africa | Northern Africa |
| **SRR8184522** | 11/10/2018 | 1.1.2.28.43.1319.11747 | Egypt | Africa | Northern Africa |
| **SRR7962275** | 20/09/2018 | 1.1.2.28.43.1319.11805 | Egypt | Africa | Northern Africa |
| **SRR7996980** | 26/09/2018 | 1.1.2.28.43.1319.11868 | Egypt | Africa | Northern Africa |
| **SRR7997115** | 24/09/2018 | 1.1.2.28.43.1319.12004 | Egypt | Africa | Northern Africa |
| **SRR8098325** | 27/09/2018 | 1.1.2.28.43.1319.12039 | Egypt | Africa | Northern Africa |
| **SRR8484193** | 02/10/2018 | 1.1.2.28.43.1319.12058 | Egypt | Africa | Northern Africa |
| **SRR8098353** | 03/10/2018 | 1.1.2.28.43.1319.12110 | Egypt | Africa | Northern Africa |
| **SRR8087140** | 09/10/2018 | 1.1.2.28.43.1319.12120 | Egypt | Africa | Northern Africa |
| **SRR8182732** | 23/10/2018 | 1.1.2.28.43.1319.12219 | Egypt | Africa | Northern Africa |
| **SRR8490773** | 23/10/2018 | 1.1.2.28.43.1319.12294 | Egypt | Africa | Northern Africa |
| **SRR8281280** | 20/11/2018 | 1.1.2.28.43.1319.12454 | Egypt | Africa | Northern Africa |
| **SRR8380678** | 17/12/2018 | 1.1.2.28.43.1319.12563 | Egypt | Africa | Northern Africa |
| **SRR8401416** | 21/12/2018 | 1.1.2.28.43.1319.12569 | Egypt | Africa | Northern Africa |
| **SRR8514503** | 22/02/2018 | 1.1.2.28.43.1319.9508 | Egypt | Africa | Northern Africa |
| **SRR7359079** | 22/02/2018 | 1.1.2.28.43.1319.9531 | Egypt | Africa | Northern Africa |
| **SRR3286680** | 23/04/2015 | 1.1.2.28.43.1923.2735 | Egypt | Africa | Northern Africa |
| **SRR7187234** | 30/04/2015 | 1.1.2.28.43.1923.2762 | Egypt | Africa | Northern Africa |
| **SRR3048944** | 19/12/2014 | 1.1.2.28.43.380.1195 | Egypt | Africa | Northern Africa |
| **SRR8281255** | 22/11/2018 | 1.1.2.28.43.380.12469 | Egypt | Africa | Northern Africa |
| **SRR5220947** | 08/04/2015 | 1.1.2.28.43.380.1376 | Egypt | Africa | Northern Africa |
| **SRR1968195** | 09/03/2015 | 1.1.2.28.43.380.1524 | Egypt | Africa | Northern Africa |
| **SRR1966497** | 26/11/2014 | 1.1.2.28.43.380.1779 | Egypt | Africa | Northern Africa |
| **SRR1968505** | 08/12/2014 | 1.1.2.28.43.380.1910 | Egypt | Africa | Northern Africa |
| **SRR1968917** | 03/02/2015 | 1.1.2.28.43.380.2120 | Egypt | Africa | Northern Africa |
| **SRR7474903** | 07/07/2015 | 1.1.2.28.43.380.3019 | Egypt | Africa | Northern Africa |
| **SRR7350790** | 14/08/2015 | 1.1.2.28.43.380.3338 | Egypt | Africa | Northern Africa |
| **SRR7414984** | 15/09/2015 | 1.1.2.28.43.380.3621 | Egypt | Africa | Northern Africa |
| **SRR5220109** | 19/10/2015 | 1.1.2.28.43.380.3796 | Egypt | Africa | Northern Africa |
| **SRR1957949** | 14/10/2014 | 1.1.2.28.43.380.387 | Egypt | Africa | Northern Africa |
| **SRR1958409** | 01/09/2014 | 1.1.2.28.43.380.403 | Egypt | Africa | Northern Africa |

| | | | | | |
|---|---|---|---|---|---|
| **SRR5220465** | 24/11/2015 | 1.1.2.28.43.380.4102 | Egypt | Africa | Northern Africa |
| **SRR3286649** | 08/05/2015 | 1.1.2.28.43.380.464 | Egypt | Africa | Northern Africa |
| **SRR6922681** | 07/11/2017 | 1.1.2.28.43.4242.8813 | Egypt | Africa | Northern Africa |
| **SRR7902690** | 16/01/2018 | 1.1.2.28.43.4242.9269 | Egypt | Africa | Northern Africa |
| **SRR1969419** | 13/05/2014 | 1.1.2.28.43.43.1407 | Egypt | Africa | Northern Africa |
| **SRR3049098** | 10/03/2015 | 1.1.2.28.43.43.1841 | Egypt | Africa | Northern Africa |
| **SRR1968428** | 08/07/2014 | 1.1.2.28.43.43.425 | Egypt | Africa | Northern Africa |
| **SRR1967719** | 23/06/2014 | 1.1.2.28.43.43.43 | Egypt | Africa | Northern Africa |
| **SRR8492397** | 19/06/2018 | 1.1.2.28.43.4782.10140 | Egypt | Africa | Northern Africa |
| **SRR8377940** | 12/12/2018 | 1.1.2.28.43.5747.12554 | Egypt | Africa | Northern Africa |
| **SRR8375206** | 01/08/2018 | 1.1.2.28.43.883.10573 | Egypt | Africa | Northern Africa |
| **SRR1959398** | 12/11/2014 | 1.1.2.28.43.883.1132 | Egypt | Africa | Northern Africa |
| **SRR8098380** | 26/09/2018 | 1.1.2.28.43.883.11873 | Egypt | Africa | Northern Africa |
| **SRR8269033** | 13/11/2018 | 1.1.2.28.43.883.12443 | Egypt | Africa | Northern Africa |
| **SRR8325628** | 03/12/2018 | 1.1.2.28.43.883.12501 | Egypt | Africa | Northern Africa |
| **SRR8492301** | 10/01/2019 | 1.1.2.28.43.883.12644 | Egypt | Africa | Northern Africa |
| **SRR8552361** | 22/01/2019 | 1.1.2.28.43.883.12675 | Egypt | Africa | Northern Africa |
| **SRR8724960** | 28/02/2019 | 1.1.2.28.43.883.12798 | Egypt | Africa | Northern Africa |
| **SRR5216488** | 12/01/2016 | 1.1.2.28.43.883.4317 | Egypt | Africa | Northern Africa |
| **SRR6898954** | 17/03/2016 | 1.1.2.28.43.883.4527 | Egypt | Africa | Northern Africa |
| **SRR6918298** | 08/06/2017 | 1.1.2.28.43.883.7442 | Egypt | Africa | Northern Africa |
| **SRR6918612** | 11/10/2017 | 1.1.2.28.43.883.8649 | Egypt | Africa | Northern Africa |
| **SRR1968985** | 08/05/2014 | 1.1.2.28.469.486.709 | Egypt | Africa | Northern Africa |
| **SRR1969736** | 05/02/2015 | 1.1.2.28.469.486.858 | Egypt | Africa | Northern Africa |
| **SRR1965408** | 15/08/2014 | 1.1.2.28.490.1718.2376 | Egypt | Africa | Northern Africa |
| **SRR6900115** | 05/12/2016 | 1.1.2.28.490.3307.6377 | Egypt | Africa | Northern Africa |
| **SRR1960070** | 07/01/2015 | 1.1.2.28.490.508.1057 | Egypt | Africa | Northern Africa |
| **SRR1960353** | 11/12/2014 | 1.1.2.28.490.508.1521 | Egypt | Africa | Northern Africa |
| **SRR1969444** | 05/12/2014 | 1.1.2.28.490.508.1707 | Egypt | Africa | Northern Africa |
| **SRR7457877** | 17/06/2015 | 1.1.2.28.490.508.2956 | Egypt | Africa | Northern Africa |
| **SRR1969196** | 01/08/2014 | 1.1.2.28.490.508.573 | Egypt | Africa | Northern Africa |
| **SRR1968083** | 04/08/2014 | 1.1.2.28.520.541.623 | Egypt | Africa | Northern Africa |
| **SRR3284692** | 13/07/2015 | 1.1.2.28.535.1168.3057 | Egypt | Africa | Northern Africa |
| **SRR1968065** | 27/08/2014 | 1.1.2.28.535.558.1014 | Egypt | Africa | Northern Africa |
| **SRR1968003** | 26/03/2015 | 1.1.2.28.535.558.2061 | Egypt | Africa | Northern Africa |
| **SRR1966775** | 16/03/2015 | 1.1.2.28.535.558.648 | Egypt | Africa | Northern Africa |
| **SRR8419406** | 21/12/2018 | 1.1.2.28.535.5758.12596 | Egypt | Africa | Northern Africa |
| **SRR1968156** | 07/10/2014 | 1.1.2.28.535.692.2095 | Egypt | Africa | Northern Africa |
| **SRR1969120** | 29/09/2014 | 1.1.2.28.535.692.847 | Egypt | Africa | Northern Africa |
| **SRR1966995** | 18/09/2014 | 1.1.2.28.535.692.954 | Egypt | Africa | Northern Africa |
| **SRR5220922** | 27/07/2015 | 1.1.2.28.538.2073.3171 | Egypt | Africa | Northern Africa |
| **SRR6918524** | 04/05/2017 | 1.1.2.28.538.3736.7241 | Egypt | Africa | Northern Africa |
| **SRR7480287** | 18/05/2015 | 1.1.2.28.538.562.2802 | Egypt | Africa | Northern Africa |
| **SRR8706125** | 28/10/2015 | 1.1.2.28.538.562.3934 | Egypt | Africa | Northern Africa |
| **SRR1965165** | 07/10/2014 | 1.1.2.28.870.1151.1532 | Egypt | Africa | Northern Africa |
| **SRR5220832** | 25/09/2015 | 1.1.2.28.870.1996.3682 | Egypt | Africa | Northern Africa |
| **SRR5216242** | 14/10/2015 | 1.1.2.28.870.1996.3817 | Egypt | Africa | Northern Africa |
| **SRR8704627** | 26/11/2015 | 1.1.2.28.870.1996.4108 | Egypt | Africa | Northern Africa |

| | | | | | |
|---|---|---|---|---|---|
| **SRR8333792** | 14/08/2018 | 1.1.2.28.870.3930.11398 | Egypt | Africa | Northern Africa |
| **SRR6922714** | 05/07/2017 | 1.1.2.28.870.3930.7719 | Egypt | Africa | Northern Africa |
| **SRR1968151** | 03/09/2014 | 1.1.2.28.870.969.1243 | Egypt | Africa | Northern Africa |
| **SRR7251497** | 03/09/2014 | 1.1.2.28.870.969.1644 | Egypt | Africa | Northern Africa |
| **SRR7501695** | 01/05/2015 | 1.1.2.32.1671.1932.2758 | Egypt | Africa | Northern Africa |
| **SRR7351421** | 01/09/2015 | 1.1.2.32.1671.1932.3458 | Egypt | Africa | Northern Africa |
| **SRR7962207** | 19/09/2018 | 1.1.2.32.1671.4117.11820 | Egypt | Africa | Northern Africa |
| **SRR8149435** | 04/10/2018 | 1.1.2.32.1671.4117.12087 | Egypt | Africa | Northern Africa |
| **SRR8106802** | 04/10/2018 | 1.1.2.32.1671.4117.12088 | Egypt | Africa | Northern Africa |
| **SRR8116974** | 16/10/2018 | 1.1.2.32.1671.4117.12189 | Egypt | Africa | Northern Africa |
| **SRR6900952** | 27/09/2017 | 1.1.2.32.1671.4157.8531 | Egypt | Africa | Northern Africa |
| **SRR7369081** | 29/07/2015 | 1.1.2.32.1766.2086.3208 | Egypt | Africa | Northern Africa |
| **SRR1957746** | 24/06/2014 | 1.1.2.359.1229.1400.1900 | Egypt | Africa | Northern Africa |
| **SRR1965767** | 26/06/2014 | 1.1.2.359.1232.1403.1904 | Egypt | Africa | Northern Africa |
| **SRR1965773** | 03/07/2014 | 1.1.2.359.1468.1694.2335 | Egypt | Africa | Northern Africa |
| **SRR1967012** | 25/07/2014 | 1.1.2.359.367.368.1085 | Egypt | Africa | Northern Africa |
| **SRR1967970** | 16/02/2015 | 1.1.2.359.367.368.2471 | Egypt | Africa | Northern Africa |
| **SRR1966966** | 18/11/2014 | 1.1.2.359.367.368.372 | Egypt | Africa | Northern Africa |
| **SRR1966337** | 23/04/2014 | 1.1.2.359.380.383.1343 | Egypt | Africa | Northern Africa |
| **SRR3049444** | 18/06/2014 | 1.1.2.359.380.383.390 | Egypt | Africa | Northern Africa |
| **SRR7850497** | 13/08/2018 | 1.1.2.359.616.5343.11416 | Egypt | Africa | Northern Africa |
| **SRR7172614** | 08/04/2015 | 1.1.2.393.425.435.2162 | Egypt | Africa | Northern Africa |
| **SRR1965349** | 03/03/2015 | 1.1.2.393.425.435.773 | Egypt | Africa | Northern Africa |
| **SRR7414772** | 10/05/2018 | 1.1.2.64.3557.4721.9944 | Egypt | Africa | Northern Africa |
| **SRR7842682** | 09/08/2018 | 1.1.2.64.64.1930.11382 | Egypt | Africa | Northern Africa |
| **SRR8754466** | 07/03/2019 | 1.1.2.64.64.1930.12820 | Egypt | Africa | Northern Africa |
| **SRR8758321** | 01/03/2019 | 1.1.2.64.64.1930.12826 | Egypt | Africa | Northern Africa |
| **SRR5194131** | 05/10/2015 | 1.1.2.64.64.1930.3714 | Egypt | Africa | Northern Africa |
| **SRR8503760** | 16/04/2018 | 1.1.2.64.64.4684.9820 | Egypt | Africa | Northern Africa |
| **SRR8484232** | 07/11/2018 | 1.1.2.64.64.5693.12387 | Egypt | Africa | Northern Africa |
| **SRR8325633** | 04/12/2018 | 1.1.2.64.64.5693.12508 | Egypt | Africa | Northern Africa |
| **SRR1966277** | 06/08/2014 | 1.1.2.64.64.64.942 | Egypt | Africa | Northern Africa |
| **SRR6920130** | 11/08/2016 | 1.1.329.1318.2284.2836.5130 | Egypt | Africa | Northern Africa |
| **SRR5215489** | 26/08/2015 | 1.11.48.154.1148.1301.3349 | Egypt | Africa | Northern Africa |
| **SRR5193782** | 13/01/2016 | 1.2.3.1229.2090.2538.4316 | Egypt | Africa | Northern Africa |
| **SRR1968543** | 31/03/2015 | 1.2.3.18.1207.1370.1858 | Egypt | Africa | Northern Africa |
| **SRR4063716** | 05/05/2015 | 1.2.3.18.359.360.2409 | Egypt | Africa | Northern Africa |
| **SRR7523772** | 07/09/2015 | 1.2.3.18.458.472.3037 | Egypt | Africa | Northern Africa |
| **SRR7528166** | 09/10/2015 | 1.2.3.18.599.639.3757 | Egypt | Africa | Northern Africa |
| **SRR1959270** | 30/12/2014 | 1.2.3.18.599.639.772 | Egypt | Africa | Northern Africa |
| **SRR5220545** | 01/09/2015 | 1.1.2.1117.1849.2197.3457 | France | Europe | Western Europe |
| **SRR6900142** | 09/08/2017 | 1.1.2.2073.2557.3210.7868 | France | Europe | Western Europe |
| **SRR6922615** | 08/09/2017 | 1.11.48.2344.3174.4098.8359 | France | Europe | Western Europe |
| **SRR8272649** | 10/09/2018 | 1.2.3.151.151.783.11676 | France | Europe | Western Europe |
| **SRR6920446** | 28/04/2016 | 1.2.3.151.151.783.4627 | France | Europe | Western Europe |
| **SRR8084193** | 09/10/2018 | 1.5.757.2642.4018.5647.12152 | France | Europe | Western Europe |
| **SRR8711633** | 20/10/2015 | 1.7.14.1169.1973.2359.3829 | France | Europe | Western Europe |
| **SRR5194224** | 01/09/2016 | 3.4.49.1671.2418.3014.5461 | France | Europe | Western Europe |

| | | | | | |
|---|---|---|---|---|---|
| **SRR1958370** | 08/10/2014 | 3.4.8.307.677.731.907 | France | Europe | Western Europe |
| **SRR5216596** | 02/06/2016 | 8.14.59.1281.2200.2701.4747 | France | Europe | Western Europe |
| **SRR5194088** | 28/09/2016 | 8.14.59.1828.2495.3122.5814 | France | Europe | Western Europe |
| **SRR5216409** | 07/09/2016 | 8.23.121.1693.2450.3054.5551 | France | Europe | Western Europe |
| **SRR6900354** | 28/09/2017 | 1.1.2.12.12.4034.8538 | Greece | Europe | Southern Europe |
| **SRR7465102** | 25/04/2018 | 1.1.2.12.421.429.9840 | Greece | Europe | Southern Europe |
| **SRR7892290** | 11/09/2018 | 1.1.2.237.3136.4038.11530 | Greece | Europe | Southern Europe |
| **SRR5632086** | 11/01/2017 | 1.1.2.28.195.2365.6857 | Greece | Europe | Southern Europe |
| **SRR6898077** | 11/10/2016 | 1.1.2.80.100.100.5932 | Greece | Europe | Southern Europe |
| **SRR6918631** | 03/10/2017 | 1.1.2.80.1903.2273.8568 | Greece | Europe | Southern Europe |
| **SRR5215537** | 15/03/2016 | 1.11.306.1254.2142.2609.4511 | Greece | Europe | Southern Europe |
| **SRR7221366** | 04/09/2015 | 1.11.48.1093.1814.2150.3348 | Greece | Europe | Southern Europe |
| **SRR7456899** | 07/09/2015 | 1.11.48.1093.1814.2150.3471 | Greece | Europe | Southern Europe |
| **SRR5216440** | 09/09/2015 | 1.11.48.1093.1814.2150.3486 | Greece | Europe | Southern Europe |
| **SRR6898447** | 15/05/2017 | 1.11.48.1309.2967.3759.7301 | Greece | Europe | Southern Europe |
| **SRR1958269** | 12/09/2014 | 1.11.48.148.148.148.148 | Greece | Europe | Southern Europe |
| **SRR8490666** | 04/07/2018 | 1.11.48.2530.3611.4822.10296 | Greece | Europe | Southern Europe |
| **SRR8490645** | 04/07/2018 | 1.11.48.2530.3611.4822.10320 | Greece | Europe | Southern Europe |
| **SRR8441124** | 02/07/2018 | 1.11.48.2532.3614.4828.10312 | Greece | Europe | Southern Europe |
| **SRR7874119** | 05/09/2018 | 1.11.48.256.3872.5411.11622 | Greece | Europe | Southern Europe |
| **SRR7841575** | 07/08/2018 | 1.11.67.104.2359.5314.11343 | Greece | Europe | Southern Europe |
| **SRR5194111** | 08/03/2016 | 1.2.3.151.1707.1980.2896 | Greece | Europe | Southern Europe |
| **SRR7841546** | 31/08/2018 | 1.2.3.18.175.175.11564 | Greece | Europe | Southern Europe |
| **SRR4063757** | 04/09/2015 | 1.2.3.18.1859.2211.3495 | Greece | Europe | Southern Europe |
| **SRR7350636** | 29/10/2015 | 1.2.3.18.323.2409.3971 | Greece | Europe | Southern Europe |
| **SRR6919936** | 19/07/2016 | 1.2.3.18.323.2409.4964 | Greece | Europe | Southern Europe |
| **SRR6920167** | 12/07/2016 | 1.2.3.18.323.2409.4977 | Greece | Europe | Southern Europe |
| **SRR5216151** | 11/08/2016 | 1.2.3.18.323.2409.5133 | Greece | Europe | Southern Europe |
| **SRR6918595** | 26/09/2016 | 1.2.3.18.323.2409.5713 | Greece | Europe | Southern Europe |
| **SRR5193595** | 26/10/2016 | 1.2.3.18.323.2409.6123 | Greece | Europe | Southern Europe |
| **SRR5194064** | 11/10/2016 | 1.2.3.18.359.360.5929 | Greece | Europe | Southern Europe |
| **SRR6920454** | 26/09/2017 | 1.2.3.18.359.360.5969 | Greece | Europe | Southern Europe |
| **SRR6897715** | 20/09/2017 | 1.2.3.18.359.360.8444 | Greece | Europe | Southern Europe |
| **SRR1957820** | 23/09/2014 | 1.2.3.18.38.38.2163 | Greece | Europe | Southern Europe |
| **SRR6919756** | 16/10/2017 | 1.2.3.18.62.4193.8661 | Greece | Europe | Southern Europe |
| **SRR7962184** | 13/09/2018 | 1.2.3.18.62.62.11720 | Greece | Europe | Southern Europe |
| **SRR1969962** | 15/08/2014 | 1.2.3.18.62.62.1640 | Greece | Europe | Southern Europe |
| **SRR1969384** | 17/09/2014 | 1.2.3.18.62.62.2072 | Greece | Europe | Southern Europe |
| **SRR6920149** | 20/09/2016 | 1.2.3.18.62.62.4841 | Greece | Europe | Southern Europe |
| **SRR6919213** | 19/07/2016 | 1.2.3.18.62.62.4994 | Greece | Europe | Southern Europe |
| **SRR5220565** | 26/08/2016 | 1.2.3.18.62.62.5044 | Greece | Europe | Southern Europe |
| **SRR6922024** | 02/09/2016 | 1.2.3.18.62.62.5089 | Greece | Europe | Southern Europe |
| **SRR5193409** | 15/08/2016 | 1.2.3.18.62.62.5137 | Greece | Europe | Southern Europe |
| **SRR5193860** | 24/08/2016 | 1.2.3.18.62.62.5232 | Greece | Europe | Southern Europe |
| **SRR5215772** | 14/09/2016 | 1.2.3.18.62.62.5250 | Greece | Europe | Southern Europe |
| **SRR6919784** | 23/08/2016 | 1.2.3.18.62.62.5252 | Greece | Europe | Southern Europe |
| **SRR5194263** | 23/08/2016 | 1.2.3.18.62.62.5275 | Greece | Europe | Southern Europe |
| **SRR5220106** | 02/09/2016 | 1.2.3.18.62.62.5464 | Greece | Europe | Southern Europe |

| | | | | | |
|---|---|---|---|---|---|
| **SRR5220670** | 06/09/2016 | 1.2.3.18.62.62.5522 | Greece | Europe | Southern Europe |
| **SRR5194019** | 13/09/2016 | 1.2.3.18.62.62.5654 | Greece | Europe | Southern Europe |
| **SRR5194258** | 31/08/2016 | 1.2.3.18.62.62.5671 | Greece | Europe | Southern Europe |
| **SRR6920477** | 20/09/2016 | 1.2.3.18.62.62.5721 | Greece | Europe | Southern Europe |
| **SRR5216326** | 04/10/2016 | 1.2.3.18.62.62.5879 | Greece | Europe | Southern Europe |
| **SRR5216187** | 13/10/2016 | 1.2.3.18.62.62.5954 | Greece | Europe | Southern Europe |
| **SRR6919764** | 16/06/2017 | 1.2.3.18.62.62.7689 | Greece | Europe | Southern Europe |
| **SRR5220314** | 15/10/2015 | 12.28.160.283.283.283.2726 | Greece | Europe | Southern Europe |
| **SRR5757214** | 07/07/2015 | 19.43.240.1058.1733.2034.3047 | Greece | Europe | Southern Europe |
| **SRR7408396** | 12/06/2018 | 69.136.711.2525.3590.4770.10105 | Greece | Europe | Southern Europe |
| **SRR1965781** | 01/10/2014 | 8.14.59.168.168.168.168 | Greece | Europe | Southern Europe |
| **SRR6900081** | 09/11/2017 | 1.1.2.174.3252.4248.8839 | China | Asia | Eastern Asia |
| **SRR7426509** | 01/05/2018 | 1.1.2.174.3252.4262.9884 | China | Asia | Eastern Asia |
| **SRR6900748** | 06/12/2017 | 1.1.2.206.3277.4302.8987 | China | Asia | Eastern Asia |
| **SRR1960611** | 02/12/2014 | 1.1.2.4.4.1165.1771 | China | Asia | Eastern Asia |
| **SRR8367072** | 11/12/2018 | 1.1.2.7.4063.5743.12544 | China | Asia | Eastern Asia |
| **SRR5193864** | 10/05/2016 | 1.1.2.7.426.2668.4649 | China | Asia | Eastern Asia |
| **SRR5220656** | 08/07/2015 | 1.1.200.491.1742.2046.3103 | China | Asia | Eastern Asia |
| **SRR6922653** | 05/05/2016 | 1.2.3.151.362.363.4662 | China | Asia | Eastern Asia |
| **SRR6919039** | 09/05/2016 | 4.70.312.1272.2185.2674.4661 | China | Asia | Eastern Asia |
| **SRR7495566** | 22/05/2018 | 1.1.2.152.152.2430.9992 | Hungary | Europe | Eastern Europe |
| **SRR6898331** | 24/10/2017 | 1.1.2.1705.3234.4212.8718 | Hungary | Europe | Eastern Europe |
| **SRR5194236** | 24/06/2016 | 1.1.2.26.26.2722.4896 | Hungary | Europe | Eastern Europe |
| **SRR1969627** | 14/01/2015 | 1.11.48.146.1630.1882.2633 | Hungary | Europe | Eastern Europe |
| **SRR7298404** | 03/12/2015 | 1.2.3.1204.2044.2473.4131 | Hungary | Europe | Eastern Europe |
| **SRR5216317** | 21/09/2016 | 1.2.3.18.175.175.5732 | Hungary | Europe | Eastern Europe |
| **SRR5193466** | 30/09/2016 | 1.2.3.18.175.175.5855 | Hungary | Europe | Eastern Europe |
| **SRR8490667** | 03/07/2018 | 1.2.3.18.175.175.9937 | Hungary | Europe | Eastern Europe |
| **SRR5220917** | 16/09/2016 | 1.2.3.18.180.180.11080 | Hungary | Europe | Eastern Europe |
| **SRR6924097** | 18/10/2017 | 1.2.3.18.2634.3335.8678 | Hungary | Europe | Eastern Europe |
| **SRR5193980** | 31/08/2016 | 1.2.3.18.3698.5100.10908 | Hungary | Europe | Eastern Europe |
| **SRR1967905** | 21/05/2014 | 1.2.3.18.458.472.517 | Hungary | Europe | Eastern Europe |
| **SRR3049341** | 28/04/2014 | 1.2.3.18.642.689.843 | Hungary | Europe | Eastern Europe |
| **SRR4063708** | 03/06/2015 | 1.2.3.22.1705.1978.2892 | Hungary | Europe | Eastern Europe |
| **SRR8201796** | 02/10/2018 | 1.2.3.22.22.3334.12052 | Hungary | Europe | Eastern Europe |
| **SRR8283330** | 19/11/2018 | 1.2.3.22.22.3334.12459 | Hungary | Europe | Eastern Europe |
| **SRR4063735** | 04/09/2015 | 1.2.3.3.1851.2200.3466 | Hungary | Europe | Eastern Europe |
| **SRR5194245** | 14/09/2016 | 1.5.464.1699.2459.3065.5596 | Hungary | Europe | Eastern Europe |
| **SRR1965831** | 15/08/2014 | 1.5.69.111.111.111.111 | Hungary | Europe | Eastern Europe |
| **SRR3321873** | 24/04/2015 | 1.1.2.1025.1663.1919.2720 | India | Asia | Southern Asia |
| **SRR5215635** | 07/10/2016 | 1.1.2.1028.1673.3149.5893 | India | Asia | Southern Asia |
| **SRR6918351** | 19/12/2017 | 1.1.2.1028.1673.4329.9049 | India | Asia | Southern Asia |
| **SRR8114870** | 27/09/2018 | 1.1.2.1028.1673.5620.12037 | India | Asia | Southern Asia |
| **SRR8704717** | 27/11/2015 | 1.1.2.1028.2048.2479.4144 | India | Asia | Southern Asia |
| **SRR8474028** | 03/01/2019 | 1.1.2.1030.4075.5768.12620 | India | Asia | Southern Asia |
| **SRR5193821** | 18/09/2015 | 1.1.2.1125.1868.2220.3511 | India | Asia | Southern Asia |
| **SRR6901074** | 23/12/2015 | 1.1.2.1218.2065.2497.4210 | India | Asia | Southern Asia |

| | | | | | |
|---|---|---|---|---|---|
| **SRR5194199** | 01/06/2016 | 1.1.2.1284.2204.2709.4766 | India | Asia | Southern Asia |
| **SRR8730996** | 08/05/2015 | 1.1.2.1326.447.461.1171 | India | Asia | Southern Asia |
| **SRR5194002** | 18/10/2016 | 1.1.2.165.165.165.5198 | India | Asia | Southern Asia |
| **SRR7274982** | 23/02/2018 | 1.1.2.178.1855.4562.9526 | India | Asia | Southern Asia |
| **SRR8325576** | 05/12/2018 | 1.1.2.178.3157.4073.12517 | India | Asia | Southern Asia |
| **SRR1966874** | 23/02/2015 | 1.1.2.2.2.2.1198 | India | Asia | Southern Asia |
| **SRR1960954** | 07/01/2015 | 1.1.2.2.2.2.1962 | India | Asia | Southern Asia |
| **SRR1966183** | 08/05/2014 | 1.1.2.2.2.2.2 | India | Asia | Southern Asia |
| **SRR1969064** | 15/05/2014 | 1.1.2.2.2.2.2443 | India | Asia | Southern Asia |
| **SRR1960057** | 09/01/2015 | 1.1.2.2.2.2.891 | India | Asia | Southern Asia |
| **SRR7458178** | 23/01/2018 | 1.1.2.2073.389.2854.8939 | India | Asia | Southern Asia |
| **SRR7286964** | 06/02/2018 | 1.1.2.2073.389.2854.9357 | India | Asia | Southern Asia |
| **SRR1969411** | 26/11/2014 | 1.1.2.214.1049.1181.1577 | India | Asia | Southern Asia |
| **SRR6924049** | 14/08/2017 | 1.1.2.2315.3095.3950.7801 | India | Asia | Southern Asia |
| **SRR8503983** | 06/04/2018 | 1.1.2.2508.3526.4658.9770 | India | Asia | Southern Asia |
| **SRR8893154** | 02/04/2019 | 1.1.2.274.274.5879.12916 | India | Asia | Southern Asia |
| **SRR5194193** | 23/11/2016 | 1.1.2.28.149.149.6341 | India | Asia | Southern Asia |
| **SRR5220291** | 16/12/2015 | 1.1.2.284.284.791.4219 | India | Asia | Southern Asia |
| **SRR1969852** | 16/09/2014 | 1.1.2.350.762.835.1061 | India | Asia | Southern Asia |
| **SRR1959229** | 24/10/2014 | 1.1.2.66.1231.1402.1902 | India | Asia | Southern Asia |
| **SRR3048821** | 21/07/2014 | 1.1.2.66.66.66.66 | India | Asia | Southern Asia |
| **SRR7426496** | 18/11/2015 | 1.1.2.994.1576.1817.4054 | India | Asia | Southern Asia |
| **SRR5220931** | 07/12/2015 | 1.1.2.994.2058.2489.4169 | India | Asia | Southern Asia |
| **SRR5193601** | 31/10/2016 | 1.1.2.994.2563.3221.6137 | India | Asia | Southern Asia |
| **SRR6897818** | 01/11/2017 | 1.1.2.994.3242.4228.8769 | India | Asia | Southern Asia |
| **SRR7443943** | 12/04/2018 | 1.1.2.994.3242.4228.9810 | India | Asia | Southern Asia |
| **SRR8114918** | 05/10/2018 | 1.2.3.18.175.175.9845 | India | Asia | Southern Asia |
| **SRR8503801** | 11/04/2018 | 1.2.3.18.180.180.10861 | India | Asia | Southern Asia |
| **SRR5216050** | 30/12/2016 | 1.2.3.18.2769.3503.6731 | India | Asia | Southern Asia |
| **SRR8131563** | 05/10/2018 | 1.2.3.18.353.1560.12090 | India | Asia | Southern Asia |
| **SRR6919272** | 16/05/2017 | 1.3.154.1021.2971.3764.7309 | India | Asia | Southern Asia |
| **SRR5193849** | 16/05/2016 | 1.3.310.1267.2177.2666.4645 | India | Asia | Southern Asia |
| **SRR8735155** | 07/05/2015 | 1.39.227.1032.1680.1945.2784 | India | Asia | Southern Asia |
| **SRR5215676** | 28/11/2016 | 1.5.159.280.280.280.6321 | India | Asia | Southern Asia |
| **SRR6901120** | 23/05/2017 | 20.53.283.2249.2939.3700.7322 | India | Asia | Southern Asia |
| **SRR8249768** | 15/11/2018 | 20.53.283.2657.4045.5705.12432 | India | Asia | Southern Asia |
| **SRR1965963** | 21/01/2015 | 4.31.193.423.488.506.568 | India | Asia | Southern Asia |
| **SRR6900943** | 08/06/2016 | 4.31.316.1287.2208.2714.4778 | India | Asia | Southern Asia |
| **SRR5193960** | 16/08/2016 | 4.31.331.1321.2291.2844.5155 | India | Asia | Southern Asia |
| **SRR6919768** | 23/10/2017 | 4.44.672.2365.3236.4215.8724 | India | Asia | Southern Asia |
| **SRR5216337** | 29/02/2016 | 56.68.304.1248.2131.2594.4470 | India | Asia | Southern Asia |
| **SRR7879567** | 17/07/2018 | 56.68.717.2536.3629.4859.10455 | India | Asia | Southern Asia |
| **SRR3049240** | 06/06/2014 | 56.9.21.24.24.24.24 | India | Asia | Southern Asia |
| **SRR8114913** | 25/09/2018 | 1.1.2.116.2151.2624.11997 | Indonesia | Asia | South-eastern Asia |
| **SRR5216407** | 30/03/2016 | 1.1.2.116.2151.2624.4542 | Indonesia | Asia | South-eastern Asia |
| **SRR5216531** | 28/09/2016 | 1.1.2.116.2151.2624.5792 | Indonesia | Asia | South-eastern Asia |

| | | | | | |
|---|---|---|---|---|---|
| **SRR3284684** | 06/11/2015 | 1.1.2.244.2013.2422.4006 | Indonesia | Asia | South-eastern Asia |
| **SRR1966512** | 12/08/2014 | 1.1.2.28.1489.1719.2377 | Indonesia | Asia | South-eastern Asia |
| **SRR5220827** | 25/06/2015 | 1.1.2.28.1725.2018.2983 | Indonesia | Asia | South-eastern Asia |
| **SRR1965651** | 05/03/2015 | 1.1.2.28.195.195.1128 | Indonesia | Asia | South-eastern Asia |
| **SRR8172801** | 09/10/2018 | 1.1.2.28.195.195.12134 | Indonesia | Asia | South-eastern Asia |
| **SRR6899360** | 06/06/2017 | 1.1.2.28.195.195.279 | Indonesia | Asia | South-eastern Asia |
| **SRR7475352** | 25/09/2015 | 1.1.2.28.195.195.3651 | Indonesia | Asia | South-eastern Asia |
| **SRR1969366** | 23/07/2014 | 1.1.2.28.195.195.658 | Indonesia | Asia | South-eastern Asia |
| **SRR3049526** | 26/01/2015 | 1.1.2.28.195.195.761 | Indonesia | Asia | South-eastern Asia |
| **SRR6919027** | 26/07/2017 | 1.1.2.28.195.195.7886 | Indonesia | Asia | South-eastern Asia |
| **SRR6922560** | 26/09/2017 | 1.1.2.28.195.195.8493 | Indonesia | Asia | South-eastern Asia |
| **SRR8724933** | 18/06/2015 | 1.1.2.28.195.2020.2986 | Indonesia | Asia | South-eastern Asia |
| **SRR7450697** | 17/08/2015 | 1.1.2.28.195.2139.3325 | Indonesia | Asia | South-eastern Asia |
| **SRR6918305** | 01/06/2016 | 1.1.2.28.195.2702.4749 | Indonesia | Asia | South-eastern Asia |
| **SRR5633142** | 24/01/2017 | 1.1.2.28.195.3041.6888 | Indonesia | Asia | South-eastern Asia |
| **SRR6918892** | 20/09/2017 | 1.1.2.28.195.3041.8446 | Indonesia | Asia | South-eastern Asia |
| **SRR7828441** | 17/07/2018 | 1.1.2.28.195.4863.10461 | Indonesia | Asia | South-eastern Asia |
| **SRR7885285** | 23/08/2018 | 1.1.2.28.195.5381.11513 | Indonesia | Asia | South-eastern Asia |
| **SRR8084281** | 02/10/2018 | 1.1.2.28.195.5629.12064 | Indonesia | Asia | South-eastern Asia |
| **SRR8758307** | 06/03/2019 | 1.1.2.28.195.5704.12823 | Indonesia | Asia | South-eastern Asia |
| **SRR8419410** | 28/12/2018 | 1.1.2.28.195.5757.12594 | Indonesia | Asia | South-eastern Asia |
| **SRR6919163** | 26/07/2016 | 1.1.2.28.2261.2800.5025 | Indonesia | Asia | South-eastern Asia |
| **SRR8509435** | 15/03/2018 | 1.1.2.28.3508.4626.9711 | Indonesia | Asia | South-eastern Asia |
| **SRR7439309** | 06/06/2018 | 1.1.2.28.3510.4630.10075 | Indonesia | Asia | South-eastern Asia |
| **SRR8149140** | 03/10/2018 | 1.1.2.28.3510.4630.12100 | Indonesia | Asia | South-eastern Asia |
| **SRR8435771** | 09/10/2018 | 1.1.2.28.3510.4630.12125 | Indonesia | Asia | South-eastern Asia |
| **SRR7480444** | 22/06/2018 | 1.1.2.28.3510.4799.10223 | Indonesia | Asia | South-eastern Asia |
| **SRR8172814** | 24/10/2018 | 1.1.2.28.3510.5680.12308 | Indonesia | Asia | South-eastern Asia |
| **SRR5194016** | 02/09/2016 | 1.1.2.28.354.2047.5331 | Indonesia | Asia | South-eastern Asia |
| **SRR5193424** | 06/01/2017 | 1.1.2.28.354.2047.6831 | Indonesia | Asia | South-eastern Asia |
| **SRR6920135** | 11/09/2017 | 1.1.2.28.354.2047.8327 | Indonesia | Asia | South-eastern Asia |
| **SRR6919994** | 27/10/2017 | 1.1.2.28.354.3302.8727 | Indonesia | Asia | South-eastern Asia |
| **SRR7415019** | 14/08/2015 | 1.1.2.28.354.354.3335 | Indonesia | Asia | South-eastern Asia |
| **SRR6900724** | 29/09/2017 | 1.1.2.28.354.3855.8562 | Indonesia | Asia | South-eastern Asia |
| **SRR7468920** | 18/06/2018 | 1.1.2.28.354.4788.10168 | Indonesia | Asia | South-eastern Asia |
| **SRR7841623** | 18/07/2018 | 1.1.2.28.354.4871.10482 | Indonesia | Asia | South-eastern Asia |
| **SRR8106794** | 08/10/2018 | 1.1.2.28.354.5640.12115 | Indonesia | Asia | South-eastern Asia |
| **SRR1969261** | 03/03/2015 | 1.1.2.28.354.574.679 | Indonesia | Asia | South-eastern Asia |
| **SRR8738279** | 04/03/2019 | 1.1.2.28.4103.5845.12811 | Indonesia | Asia | South-eastern Asia |
| **SRR6919115** | 18/07/2017 | 1.1.2.30.2558.3971.7902 | Indonesia | Asia | South-eastern Asia |
| **SRR7528052** | 04/08/2015 | 1.1.2.35.1779.2100.3240 | Indonesia | Asia | South-eastern Asia |
| **SRR5220902** | 12/08/2015 | 1.1.2.35.1789.2114.3279 | Indonesia | Asia | South-eastern Asia |
| **SRR6922564** | 23/10/2017 | 1.1.2.35.1789.4196.8725 | Indonesia | Asia | South-eastern Asia |
| **SRR8361477** | 07/08/2018 | 1.1.2.35.3832.5318.11358 | Indonesia | Asia | South-eastern Asia |
| **SRR7457892** | 20/06/2018 | 1.1.2.4.3525.4785.10154 | Indonesia | Asia | South-eastern Asia |
| **SRR6922515** | 05/04/2017 | 1.1.226.1027.1668.3704.7145 | Indonesia | Asia | South-eastern Asia |
| **SRR8717173** | 12/08/2015 | 1.1.99.172.1796.2126.3300 | Indonesia | Asia | South-eastern Asia |
| **SRR6919716** | 11/10/2017 | 4.125.670.2360.3224.4190.865 | Indonesia | Asia | South-eastern Asia |

| | | | | | |
|---|---|---|---|---|---|
| | | 0 | | | |
| **SRR1952912** | 14/10/2014 | 1.1.2.2069.368.369.617 | Italy | Europe | Southern Europe |
| **SRR6919765** | 02/08/2016 | 1.1.2.28.2281.2830.5112 | Italy | Europe | Southern Europe |
| **SRR6924105** | 15/06/2016 | 1.1.2.313.313.1470.4832 | Italy | Europe | Southern Europe |
| **SRR4063740** | 08/07/2015 | 1.2.3.18.1747.2052.3117 | Italy | Europe | Southern Europe |
| **SRR5216576** | 28/10/2016 | 1.2.3.18.175.175.6133 | Italy | Europe | Southern Europe |
| **SRR6900985** | 19/09/2017 | 1.2.3.18.175.175.8436 | Italy | Europe | Southern Europe |
| **SRR6918527** | 26/09/2017 | 1.2.3.18.175.175.8491 | Italy | Europe | Southern Europe |
| **SRR8492291** | 07/06/2018 | 1.2.3.18.180.180.10985 | Italy | Europe | Southern Europe |
| **SRR6897019** | 06/06/2017 | 1.2.3.18.180.180.11098 | Italy | Europe | Southern Europe |
| **SRR8293798** | 05/09/2018 | 1.2.3.18.180.180.11639 | Italy | Europe | Southern Europe |
| **SRR8293774** | 04/09/2018 | 1.2.3.18.1836.5419.11640 | Italy | Europe | Southern Europe |
| **SRR6922495** | 27/09/2017 | 1.2.3.18.455.4150.8511 | Italy | Europe | Southern Europe |
| **SRR5216517** | 21/08/2015 | 3.4.49.1113.1844.2189.3443 | Italy | Europe | Southern Europe |
| **SRR8333801** | 15/08/2018 | 1.2.3.18.180.180.11457 | Jamaica | Americas | Latin America and the Caribbean |
| **SRR1967222** | 08/07/2014 | 1.5.11.13.13.13.1032 | Jamaica | Americas | Latin America and the Caribbean |
| **SRR7533376** | 03/07/2018 | 1.5.11.13.13.13.10364 | Jamaica | Americas | Latin America and the Caribbean |
| **SRR7841506** | 13/07/2018 | 1.5.11.13.13.13.10432 | Jamaica | Americas | Latin America and the Caribbean |
| **SRR1959498** | 23/10/2014 | 1.5.11.13.13.13.1322 | Jamaica | Americas | Latin America and the Caribbean |
| **SRR5215733** | 13/09/2016 | 1.5.11.13.13.13.1872 | Jamaica | Americas | Latin America and the Caribbean |
| **SRR1968827** | 05/03/2015 | 1.5.11.13.13.13.2608 | Jamaica | Americas | Latin America and the Caribbean |
| **SRR7237453** | 16/07/2015 | 1.5.11.13.13.13.3109 | Jamaica | Americas | Latin America and the Caribbean |
| **SRR6901020** | 21/07/2016 | 1.5.11.13.13.13.5007 | Jamaica | Americas | Latin America and the Caribbean |
| **SRR5194172** | 17/10/2016 | 1.5.11.13.13.13.5962 | Jamaica | Americas | Latin America and the Caribbean |
| **SRR5633183** | 23/01/2017 | 1.5.11.13.13.13.6902 | Jamaica | Americas | Latin America and the Caribbean |
| **SRR7890247** | 01/02/2018 | 1.5.11.13.13.13.9346 | Jamaica | Americas | Latin America and the Caribbean |
| **SRR8509106** | 11/04/2018 | 1.5.11.13.13.13.9427 | Jamaica | Americas | Latin America and the Caribbean |
| **SRR6901050** | 31/08/2017 | 1.5.11.13.13.13.9594 | Jamaica | Americas | Latin America and the Caribbean |
| **SRR7410304** | 27/03/2018 | 1.5.11.13.13.13.9787 | Jamaica | Americas | Latin America and the Caribbean |
| **SRR7414404** | 23/04/2018 | 1.5.11.13.13.13.9846 | Jamaica | Americas | Latin America and the Caribbean |
| **SRR1958571** | 31/10/2014 | 1.5.11.13.130.130.1276 | Jamaica | Americas | Latin America and the Caribbean |
| **SRR3049698** | 30/05/2014 | 1.5.11.13.130.130.911 | Jamaica | Americas | Latin America and the Caribbean |
| **SRR3048722** | 16/06/2014 | 1.5.11.13.333.333.1455 | Jamaica | Americas | Latin America and the Caribbean |
| **SRR3049742** | 06/06/2014 | 1.5.11.13.333.333.376 | Jamaica | Americas | Latin America and the Caribbean |
| **SRR5193131** | 02/06/2016 | 1.5.11.13.333.991.4763 | Jamaica | Americas | Latin America and the Caribbean |
| **SRR6898461** | 06/03/2017 | 1.5.11.13.333.991.7041 | Jamaica | Americas | Latin America and the Caribbean |
| **SRR5632019** | 07/03/2017 | 1.5.11.13.333.991.7050 | Jamaica | Americas | Latin America and the Caribbean |
| **SRR6899500** | 16/05/2017 | 1.5.11.13.333.991.7302 | Jamaica | Americas | Latin America and the Caribbean |
| **SRR7879668** | 31/08/2018 | 1.5.11.2554.3864.5394.11560 | Jamaica | Americas | Latin America and the Caribbean |

| | | | | | |
|---|---|---|---|---|---|
| **SRR5215544** | 23/11/2015 | 1.5.61.92.92.92.4151 | Jamaica | Americas | Latin America and the Caribbean |
| **SRR5220418** | 04/08/2015 | 1.5.79.1029.1784.2106.3251 | Jamaica | Americas | Latin America and the Caribbean |
| **SRR6922513** | 01/12/2017 | 1.5.79.1857.3270.4294.8967 | Jamaica | Americas | Latin America and the Caribbean |
| **SRR1967178** | 04/11/2014 | 1.1.2.216.216.216.216 | Kenya | Africa | Sub-Saharan Africa |
| **SRR1966781** | 16/02/2015 | 1.1.2.249.1596.1841.2571 | Kenya | Africa | Sub-Saharan Africa |
| **SRR1966972** | 28/01/2015 | 1.1.2.249.1596.1841.2671 | Kenya | Africa | Sub-Saharan Africa |
| **SRR8499343** | 09/01/2019 | 1.1.2.2666.4079.5776.12637 | Kenya | Africa | Sub-Saharan Africa |
| **SRR6921944** | 16/08/2017 | 1.1.2.28.3135.4037.8142 | Kenya | Africa | Sub-Saharan Africa |
| **SRR6897048** | 05/10/2017 | 1.2.3.18.3219.4180.8597 | Kenya | Africa | Sub-Saharan Africa |
| **SRR8504934** | 21/03/2018 | 1.2.3.18.3256.4252.9740 | Kenya | Africa | Sub-Saharan Africa |
| **SRR8097936** | 09/10/2018 | 1.2.3.18.3256.5644.12128 | Kenya | Africa | Sub-Saharan Africa |
| **SRR5193300** | 09/02/2016 | 1.2.3.18.365.1854.4417 | Kenya | Africa | Sub-Saharan Africa |
| **SRR5193056** | 10/03/2016 | 1.2.3.18.365.2545.4503 | Kenya | Africa | Sub-Saharan Africa |
| **SRR4063733** | 02/03/2016 | 1.2.3.18.365.2591.4461 | Kenya | Africa | Sub-Saharan Africa |
| **SRR5216153** | 08/03/2016 | 1.2.3.18.365.2598.4495 | Kenya | Africa | Sub-Saharan Africa |
| **SRR5194164** | 21/04/2016 | 1.2.3.18.365.2646.4594 | Kenya | Africa | Sub-Saharan Africa |
| **SRR5216523** | 18/04/2016 | 1.2.3.18.365.2646.4602 | Kenya | Africa | Sub-Saharan Africa |
| **SRR5193047** | 23/08/2016 | 1.2.3.18.365.2890.5271 | Kenya | Africa | Sub-Saharan Africa |
| **SRR7538796** | 29/06/2018 | 1.2.3.18.365.3680.10311 | Kenya | Africa | Sub-Saharan Africa |
| **SRR8366142** | 11/12/2018 | 1.2.3.18.365.384.12550 | Kenya | Africa | Sub-Saharan Africa |
| **SRR4063745** | 30/04/2015 | 1.2.3.18.365.384.2769 | Kenya | Africa | Sub-Saharan Africa |
| **SRR1963280** | 23/10/2014 | 1.2.3.18.365.384.391 | Kenya | Africa | Sub-Saharan Africa |
| **SRR1967682** | 17/02/2015 | 1.2.3.18.365.384.475 | Kenya | Africa | Sub-Saharan Africa |
| **SRR1967747** | 07/05/2014 | 1.2.3.18.365.384.622 | Kenya | Africa | Sub-Saharan Africa |
| **SRR8524502** | 18/01/2018 | 1.2.3.18.365.384.9283 | Kenya | Africa | Sub-Saharan Africa |
| **SRR8499356** | 01/05/2018 | 1.2.3.18.365.384.9885 | Kenya | Africa | Sub-Saharan Africa |
| **SRR6898380** | 27/11/2017 | 1.2.3.18.365.4278.8927 | Kenya | Africa | Sub-Saharan Africa |
| **SRR8503747** | 04/04/2018 | 1.2.3.18.365.4660.9778 | Kenya | Africa | Sub-Saharan Africa |
| **SRR5216492** | 11/02/2016 | 1.2.3.18.365.543.4421 | Kenya | Africa | Sub-Saharan Africa |
| **SRR8735126** | 26/02/2019 | 1.2.3.18.365.5842.12799 | Kenya | Africa | Sub-Saharan Africa |
| **SRR8754453** | 08/03/2019 | 1.2.3.18.365.5847.12815 | Kenya | Africa | Sub-Saharan Africa |
| **SRR8717212** | 27/02/2019 | 1.2.3.18.4049.5718.12787 | Kenya | Africa | Sub-Saharan Africa |
| **SRR5193277** | 08/11/2016 | 1.1.2.80.349.1012.6248 | China | Asia | Eastern Asia |
| **SRR5216527** | 28/10/2016 | 1.1.2.80.80.80.6128 | China | Asia | Eastern Asia |
| **SRR8711780** | 05/11/2015 | 1.1.185.1185.2011.2418.3987 | Malaysia | Asia | South-eastern Asia |
| **SRR5215538** | 25/08/2015 | 1.1.2.1094.1816.2152.3353 | Malaysia | Asia | South-eastern Asia |
| **SRR5583891** | 11/04/2017 | 1.1.2.1094.2516.3706.7148 | Malaysia | Asia | South-eastern Asia |
| **SRR6900936** | 06/09/2016 | 1.1.2.1690.2443.3045.5530 | Malaysia | Asia | South-eastern Asia |
| **SRR6898400** | 31/03/2016 | 1.1.2.206.1728.2022.4550 | Malaysia | Asia | South-eastern Asia |
| **SRR6921946** | 03/05/2017 | 1.1.2.206.1980.3725.7216 | Malaysia | Asia | South-eastern Asia |
| **SRR5193720** | 05/07/2016 | 1.1.2.206.2240.2770.4930 | Malaysia | Asia | South-eastern Asia |
| **SRR6900307** | 03/05/2017 | 1.1.2.206.2960.3739.7250 | Malaysia | Asia | South-eastern Asia |
| **SRR6922109** | 02/02/2017 | 1.1.2.206.508.3595.6921 | Malaysia | Asia | South-eastern Asia |
| **SRR1969665** | 05/12/2014 | 1.1.2.206.508.529.603 | Malaysia | Asia | South-eastern Asia |
| **SRR6900362** | 29/09/2017 | 1.1.2.206.508.529.8527 | Malaysia | Asia | South-eastern Asia |
| **SRR1968949** | 16/05/2014 | 1.1.2.206.939.1054.1390 | Malaysia | Asia | South-eastern Asia |
| **SRR7533390** | 12/12/2017 | 1.1.2.2381.3287.4319.9020 | Malaysia | Asia | South-eastern Asia |

| | | | | | |
|---|---|---|---|---|---|
| **SRR5194233** | 02/09/2016 | 1.1.2.28.2255.2793.5450 | Malaysia | Asia | South-eastern Asia |
| **SRR6920562** | 14/09/2017 | 1.1.2.28.3180.4108.8390 | Malaysia | Asia | South-eastern Asia |
| **SRR7841571** | 13/08/2018 | 1.1.2.30.2558.3211.11412 | Malaysia | Asia | South-eastern Asia |
| **SRR8503771** | 05/04/2018 | 1.1.2.4.3525.4657.9767 | Malaysia | Asia | South-eastern Asia |
| **SRR8503649** | 09/04/2018 | 1.1.2.4.3525.4662.9783 | Malaysia | Asia | South-eastern Asia |
| **SRR7867042** | 24/07/2018 | 1.1.2.4.3635.4879.10516 | Malaysia | Asia | South-eastern Asia |
| **SRR1970240** | 26/03/2015 | 1.1.2.4.4.1909.2688 | Malaysia | Asia | South-eastern Asia |
| **SRR5220846** | 04/11/2015 | 1.1.2.4.4.2384.4000 | Malaysia | Asia | South-eastern Asia |
| **SRR8272295** | 13/11/2018 | 1.1.2.4.4.5320.11362 | Malaysia | Asia | South-eastern Asia |
| **SRR8401399** | 20/12/2018 | 1.1.2.4.4.5755.12589 | Malaysia | Asia | South-eastern Asia |
| **SRR1966804** | 18/06/2014 | 1.1.2.44.44.44.44 | Malaysia | Asia | South-eastern Asia |
| **SRR6897651** | 06/09/2017 | 1.1.200.491.2948.3718.8312 | Malaysia | Asia | South-eastern Asia |
| **SRR6920005** | 24/01/2017 | 1.1.226.1700.2840.3588.6890 | Malaysia | Asia | South-eastern Asia |
| **SRR8509131** | 16/03/2018 | 1.1.226.2505.3509.4627.9712 | Malaysia | Asia | South-eastern Asia |
| **SRR9287742** | 17/04/2019 | 1.1.226.2681.4122.5896.12979 | Malaysia | Asia | South-eastern Asia |
| **SRR7962116** | 14/09/2018 | 4.111.631.2564.3888.5442.117 32 | Malaysia | Asia | South-eastern Asia |
| **SRR1969696** | 09/09/2014 | 1.1.2.138.138.138.138 | Malta | Europe | Southern Europe |
| **SRR8166151** | 11/09/2018 | 1.1.2.2073.3883.5430.11688 | Malta | Europe | Southern Europe |
| **SRR1966179** | 21/10/2014 | 1.1.2.210.210.210.210 | Malta | Europe | Southern Europe |
| **SRR3286605** | 16/04/2015 | 1.1.2.229.229.229.1804 | Malta | Europe | Southern Europe |
| **SRR8492317** | 05/06/2018 | 1.1.2.229.3578.4757.10078 | Malta | Europe | Southern Europe |
| **SRR8300441** | 30/08/2018 | 1.2.3.18.180.180.11577 | Malta | Europe | Southern Europe |
| **SRR7962258** | 21/08/2018 | 1.2.3.18.183.183.11448 | Malta | Europe | Southern Europe |
| **SRR8489346** | 14/01/2019 | 1.2.3.18.183.183.12649 | Malta | Europe | Southern Europe |
| **SRR1963285** | 31/12/2014 | 1.2.3.18.183.183.183 | Malta | Europe | Southern Europe |
| **SRR5193751** | 29/06/2015 | 1.2.3.18.183.183.3005 | Malta | Europe | Southern Europe |
| **SRR7348943** | 14/09/2015 | 1.2.3.18.183.183.3454 | Malta | Europe | Southern Europe |
| **SRR5193807** | 22/10/2015 | 1.2.3.18.183.183.3902 | Malta | Europe | Southern Europe |
| **SRR7516698** | 01/12/2015 | 1.2.3.18.183.183.4157 | Malta | Europe | Southern Europe |
| **SRR5193544** | 10/02/2016 | 1.2.3.18.183.183.4414 | Malta | Europe | Southern Europe |
| **SRR6900486** | 15/02/2016 | 1.2.3.18.183.183.4422 | Malta | Europe | Southern Europe |
| **SRR5215503** | 15/04/2016 | 1.2.3.18.183.183.4579 | Malta | Europe | Southern Europe |
| **SRR5193070** | 13/04/2016 | 1.2.3.18.183.183.4581 | Malta | Europe | Southern Europe |
| **SRR5194125** | 19/04/2016 | 1.2.3.18.183.183.4590 | Malta | Europe | Southern Europe |
| **SRR6900356** | 26/04/2016 | 1.2.3.18.183.183.4614 | Malta | Europe | Southern Europe |
| **SRR5215648** | 28/04/2016 | 1.2.3.18.183.183.4630 | Malta | Europe | Southern Europe |
| **SRR5215726** | 17/05/2016 | 1.2.3.18.183.183.4681 | Malta | Europe | Southern Europe |
| **SRR5194146** | 31/08/2016 | 1.2.3.18.183.183.5306 | Malta | Europe | Southern Europe |
| **SRR6924051** | 03/08/2017 | 1.2.3.18.183.183.7918 | Malta | Europe | Southern Europe |
| **SRR8509046** | 15/03/2018 | 1.2.3.18.183.183.9713 | Malta | Europe | Southern Europe |
| **SRR6919322** | 10/10/2017 | 1.2.3.18.183.4178.8593 | Malta | Europe | Southern Europe |
| **SRR9287472** | 09/04/2019 | 1.2.3.18.211.211.12956 | Malta | Europe | Southern Europe |
| **SRR1958089** | 23/09/2014 | 1.2.3.18.408.415.434 | Malta | Europe | Southern Europe |
| **SRR1963439** | 24/12/2014 | 1.2.3.18.408.616.738 | Malta | Europe | Southern Europe |
| **SRR3585355** | 23/10/2015 | 1.1.2.1177.1995.2391.3912 | Mexico | Americas | Latin America and the Caribbean |
| **SRR7495739** | 29/10/2015 | 1.1.2.1182.2007.2412.3976 | Mexico | Americas | Latin America and the Caribbean |
| **SRR6919304** | 06/09/2017 | 1.1.2.1182.3168.4091.8342 | Mexico | Americas | Latin America and the |

| | | | | | |
|---|---|---|---|---|---|
| | | | | | Caribbean |
| **SRR5193116** | 02/03/2016 | 1.1.2.12.12.590.3896 | Mexico | Americas | Latin America and the Caribbean |
| **SRR7465049** | 28/04/2015 | 1.1.2.80.349.1012.2768 | Mexico | Americas | Latin America and the Caribbean |
| **SRR1965636** | 27/03/2015 | 1.1.204.580.786.862.1101 | Mexico | Americas | Latin America and the Caribbean |
| **SRR3286863** | 15/06/2015 | 1.1.237.1051.1716.2001.2940 | Mexico | Americas | Latin America and the Caribbean |
| **SRR5193245** | 11/02/2016 | 1.1.333.103.103.2090.4416 | Mexico | Americas | Latin America and the Caribbean |
| **SRR7275014** | 23/05/2018 | 1.1.333.103.103.2119.10014 | Mexico | Americas | Latin America and the Caribbean |
| **SRR6900640** | 03/05/2016 | 1.1.333.103.103.2119.4731 | Mexico | Americas | Latin America and the Caribbean |
| **SRR6900341** | 28/03/2017 | 1.1.333.103.103.2119.7106 | Mexico | Americas | Latin America and the Caribbean |
| **SRR6897949** | 09/05/2017 | 1.1.333.103.103.2119.7289 | Mexico | Americas | Latin America and the Caribbean |
| **SRR6924127** | 28/09/2017 | 1.1.333.103.103.2119.8530 | Mexico | Americas | Latin America and the Caribbean |
| **SRR8524667** | 15/11/2017 | 1.1.333.103.103.2119.9016 | Mexico | Americas | Latin America and the Caribbean |
| **SRR6897821** | 15/12/2017 | 1.1.333.103.103.2119.9038 | Mexico | Americas | Latin America and the Caribbean |
| **SRR7456781** | 14/02/2018 | 1.1.333.103.103.2119.9479 | Mexico | Americas | Latin America and the Caribbean |
| **SRR7204609** | 05/04/2018 | 1.1.333.103.103.2119.9766 | Mexico | Americas | Latin America and the Caribbean |
| **SRR7495458** | 20/04/2018 | 1.1.333.103.103.2119.9847 | Mexico | Americas | Latin America and the Caribbean |
| **SRR6900084** | 26/09/2017 | 1.1.333.103.103.4147.8503 | Mexico | Americas | Latin America and the Caribbean |
| **SRR7350602** | 13/04/2018 | 1.1.333.103.103.4682.9818 | Mexico | Americas | Latin America and the Caribbean |
| **SRR7439504** | 14/06/2018 | 1.1.333.103.103.4777.10125 | Mexico | Americas | Latin America and the Caribbean |
| **SRR1966216** | 08/07/2014 | 1.1.333.103.103.548.1391 | Mexico | Americas | Latin America and the Caribbean |
| **SRR1965348** | 04/08/2014 | 1.1.333.103.103.548.1738 | Mexico | Americas | Latin America and the Caribbean |
| **SRR1966970** | 03/02/2015 | 1.1.333.103.103.548.1813 | Mexico | Americas | Latin America and the Caribbean |
| **SRR1960181** | 09/12/2014 | 1.1.333.103.103.548.2165 | Mexico | Americas | Latin America and the Caribbean |
| **SRR1966236** | 01/08/2014 | 1.1.333.103.103.548.2639 | Mexico | Americas | Latin America and the Caribbean |
| **SRR1968641** | 29/01/2015 | 1.1.333.103.103.548.2648 | Mexico | Americas | Latin America and the Caribbean |
| **SRR3286852** | 09/06/2015 | 1.1.333.103.103.548.2917 | Mexico | Americas | Latin America and the Caribbean |
| **SRR7469064** | 16/06/2015 | 1.1.333.103.103.548.2943 | Mexico | Americas | Latin America and the Caribbean |
| **SRR3286930** | 18/06/2015 | 1.1.333.103.103.548.2968 | Mexico | Americas | Latin America and the Caribbean |
| **SRR3284840** | 23/09/2015 | 1.1.333.103.103.548.3600 | Mexico | Americas | Latin America and the Caribbean |
| **SRR5193896** | 15/09/2015 | 1.1.333.103.103.548.3617 | Mexico | Americas | Latin America and the Caribbean |
| **SRR5220460** | 23/09/2015 | 1.1.333.103.103.548.3632 | Mexico | Americas | Latin America and the Caribbean |
| **SRR7426628** | 07/10/2015 | 1.1.333.103.103.548.3720 | Mexico | Americas | Latin America and the Caribbean |
| **SRR5220888** | 08/10/2015 | 1.1.333.103.103.548.3793 | Mexico | Americas | Latin America and the Caribbean |
| **SRR5193303** | 14/10/2015 | 1.1.333.103.103.548.3823 | Mexico | Americas | Latin America and the Caribbean |
| **SRR5193389** | 16/10/2015 | 1.1.333.103.103.548.3828 | Mexico | Americas | Latin America and the Caribbean |
| **SRR5215512** | 20/10/2015 | 1.1.333.103.103.548.3871 | Mexico | Americas | Latin America and the Caribbean |

| | | | | | |
|---|---|---|---|---|---|
| **SRR8704853** | 02/12/2015 | 1.1.333.103.103.548.4121 | Mexico | Americas | Latin America and the Caribbean |
| **SRR5194034** | 31/12/2015 | 1.1.333.103.103.548.4239 | Mexico | Americas | Latin America and the Caribbean |
| **SRR5193814** | 05/02/2016 | 1.1.333.103.103.548.4407 | Mexico | Americas | Latin America and the Caribbean |
| **SRR5216562** | 03/03/2016 | 1.1.333.103.103.548.4481 | Mexico | Americas | Latin America and the Caribbean |
| **SRR5194137** | 11/04/2016 | 1.1.333.103.103.548.4577 | Mexico | Americas | Latin America and the Caribbean |
| **SRR7122569** | 07/06/2016 | 1.1.333.103.103.548.4815 | Mexico | Americas | Latin America and the Caribbean |
| **SRR5194318** | 12/07/2016 | 1.1.333.103.103.548.4980 | Mexico | Americas | Latin America and the Caribbean |
| **SRR5193620** | 25/07/2016 | 1.1.333.103.103.548.5035 | Mexico | Americas | Latin America and the Caribbean |
| **SRR3286685** | 12/05/2015 | 1.1.333.103.103.548.5188 | Mexico | Americas | Latin America and the Caribbean |
| **SRR6919816** | 01/09/2016 | 1.1.333.103.103.548.5415 | Mexico | Americas | Latin America and the Caribbean |
| **SRR5220699** | 14/09/2016 | 1.1.333.103.103.548.5600 | Mexico | Americas | Latin America and the Caribbean |
| **SRR5193787** | 23/09/2016 | 1.1.333.103.103.548.5744 | Mexico | Americas | Latin America and the Caribbean |
| **SRR6900181** | 20/09/2016 | 1.1.333.103.103.548.5757 | Mexico | Americas | Latin America and the Caribbean |
| **SRR6922558** | 04/10/2016 | 1.1.333.103.103.548.5878 | Mexico | Americas | Latin America and the Caribbean |
| **SRR7351319** | 01/04/2015 | 1.1.333.103.103.548.637 | Mexico | Americas | Latin America and the Caribbean |
| **SRR5632726** | 24/04/2017 | 1.1.333.103.103.548.7204 | Mexico | Americas | Latin America and the Caribbean |
| **SRR6919918** | 15/05/2017 | 1.1.333.103.103.548.7296 | Mexico | Americas | Latin America and the Caribbean |
| **SRR6922073** | 18/08/2017 | 1.1.333.103.103.548.8186 | Mexico | Americas | Latin America and the Caribbean |
| **SRR7163836** | 04/01/2018 | 1.1.333.103.103.548.9181 | Mexico | Americas | Latin America and the Caribbean |
| **SRR8137395** | 17/10/2018 | 1.1.333.103.103.5669.12230 | Mexico | Americas | Latin America and the Caribbean |
| **SRR1966960** | 26/02/2015 | 1.1.333.103.103.670.1058 | Mexico | Americas | Latin America and the Caribbean |
| **SRR7998240** | 26/09/2018 | 1.1.333.103.103.670.11844 | Mexico | Americas | Latin America and the Caribbean |
| **SRR8731025** | 28/05/2015 | 1.1.333.103.103.670.2850 | Mexico | Americas | Latin America and the Caribbean |
| **SRR7349202** | 09/06/2015 | 1.1.333.103.103.670.2906 | Mexico | Americas | Latin America and the Caribbean |
| **SRR8705999** | 05/10/2015 | 1.1.333.103.103.670.3717 | Mexico | Americas | Latin America and the Caribbean |
| **SRR6900342** | 04/05/2016 | 1.1.333.103.103.670.4652 | Mexico | Americas | Latin America and the Caribbean |
| **SRR6922090** | 30/06/2016 | 1.1.333.103.103.670.4904 | Mexico | Americas | Latin America and the Caribbean |
| **SRR6918882** | 07/09/2016 | 1.1.333.103.103.670.5546 | Mexico | Americas | Latin America and the Caribbean |
| **SRR6922075** | 05/10/2016 | 1.1.333.103.103.670.5882 | Mexico | Americas | Latin America and the Caribbean |
| **SRR5193925** | 29/11/2016 | 1.1.333.103.103.670.5940 | Mexico | Americas | Latin America and the Caribbean |
| **SRR5220696** | 16/12/2016 | 1.1.333.103.103.670.6523 | Mexico | Americas | Latin America and the Caribbean |
| **SRR6900311** | 02/05/2017 | 1.1.333.103.103.670.7217 | Mexico | Americas | Latin America and the Caribbean |
| **SRR6898867** | 13/06/2017 | 1.1.333.103.103.670.7538 | Mexico | Americas | Latin America and the Caribbean |
| **SRR7458549** | 14/02/2018 | 1.1.333.103.103.670.9474 | Mexico | Americas | Latin America and the Caribbean |
| **SRR7216057** | 08/03/2018 | 1.1.333.103.103.670.9684 | Mexico | Americas | Latin America and the Caribbean |
| **SRR3285386** | 23/10/2015 | 1.1.333.103.1992.2387.3906 | Mexico | Americas | Latin America and the Caribbean |

| | | | | | |
|---|---|---|---|---|---|
| **SRR6919279** | 21/07/2016 | 1.1.333.103.1992.2387.5002 | Mexico | Americas | Latin America and the Caribbean |
| **SRR5215808** | 23/09/2016 | 1.1.333.103.1992.2387.5788 | Mexico | Americas | Latin America and the Caribbean |
| **SRR6920106** | 22/09/2016 | 1.1.333.103.1992.2387.5790 | Mexico | Americas | Latin America and the Caribbean |
| **SRR6901088** | 15/05/2017 | 1.1.333.103.1992.2387.7292 | Mexico | Americas | Latin America and the Caribbean |
| **SRR6898885** | 27/06/2016 | 1.1.333.103.1992.2757.4891 | Mexico | Americas | Latin America and the Caribbean |
| **SRR5220664** | 11/01/2017 | 1.1.333.103.1992.2757.6854 | Mexico | Americas | Latin America and the Caribbean |
| **SRR5193656** | 27/10/2016 | 1.1.333.103.2561.3217.6126 | Mexico | Americas | Latin America and the Caribbean |
| **SRR6900868** | 23/05/2017 | 1.1.333.103.2977.3770.7334 | Mexico | Americas | Latin America and the Caribbean |
| **SRR8087208** | 08/10/2018 | 1.1.333.103.4016.5636.12091 | Mexico | Americas | Latin America and the Caribbean |
| **SRR6898847** | 28/09/2017 | 1.1.669.2356.3206.4160.8539 | Mexico | Americas | Latin America and the Caribbean |
| **SRR5633347** | 24/04/2017 | 1.2.3.18.2950.3720.7211 | Mexico | Americas | Latin America and the Caribbean |
| **SRR7343913** | 04/09/2015 | 1.1.2.1023.1657.2192.3447 | Morocco | Africa | Northern Africa |
| **SRR8503796** | 03/05/2018 | 1.1.2.12.12.590.9897 | Morocco | Africa | Northern Africa |
| **SRR1962286** | 31/12/2014 | 1.1.2.150.1196.1357.1837 | Morocco | Africa | Northern Africa |
| **SRR1958651** | 12/09/2014 | 1.1.2.150.150.150.150 | Morocco | Africa | Northern Africa |
| **SRR6919751** | 16/11/2016 | 1.1.2.1886.2604.3278.6278 | Morocco | Africa | Northern Africa |
| **SRR7358931** | 17/11/2015 | 1.1.2.27.27.2456.4075 | Morocco | Africa | Northern Africa |
| **SRR1968172** | 19/06/2014 | 1.1.2.27.27.27.27 | Morocco | Africa | Northern Africa |
| **SRR8706113** | 01/10/2015 | 1.1.2.61.61.561.3653 | Morocco | Africa | Northern Africa |
| **SRR5193140** | 27/04/2016 | 1.1.2.80.2167.2650.4611 | Morocco | Africa | Northern Africa |
| **SRR7885280** | 21/08/2018 | 1.1.2.80.2167.3333.11449 | Morocco | Africa | Northern Africa |
| **SRR6898011** | 01/11/2017 | 1.1.2.80.2167.3333.8768 | Morocco | Africa | Northern Africa |
| **SRR6897044** | 05/04/2017 | 1.1.2.80.2942.3705.7147 | Morocco | Africa | Northern Africa |
| **SRR6899269** | 12/04/2017 | 1.1.2.80.2942.3705.7182 | Morocco | Africa | Northern Africa |
| **SRR3286624** | 20/04/2015 | 1.1.2.80.349.1012.2676 | Morocco | Africa | Northern Africa |
| **SRR7538384** | 28/04/2015 | 1.1.2.80.349.1012.2768 | Morocco | Africa | Northern Africa |
| **SRR5194294** | 14/04/2016 | 1.1.2.80.349.1012.4596 | Morocco | Africa | Northern Africa |
| **SRR5193731** | 14/04/2016 | 1.1.2.80.349.1012.4597 | Morocco | Africa | Northern Africa |
| **SRR6922066** | 19/04/2016 | 1.1.2.80.349.1012.4604 | Morocco | Africa | Northern Africa |
| **SRR5193482** | 24/08/2016 | 1.1.2.80.349.1012.5208 | Morocco | Africa | Northern Africa |
| **SRR8549079** | 15/08/2017 | 1.1.2.80.349.1012.5795 | Morocco | Africa | Northern Africa |
| **SRR5220631** | 27/09/2016 | 1.1.2.80.349.1012.5798 | Morocco | Africa | Northern Africa |
| **SRR5220671** | 26/09/2016 | 1.1.2.80.349.1012.5817 | Morocco | Africa | Northern Africa |
| **SRR5193538** | 04/10/2016 | 1.1.2.80.349.1012.5870 | Morocco | Africa | Northern Africa |
| **SRR5194176** | 11/10/2016 | 1.1.2.80.349.1012.5919 | Morocco | Africa | Northern Africa |
| **SRR6900118** | 10/10/2017 | 1.1.2.80.349.1012.8606 | Morocco | Africa | Northern Africa |
| **SRR6920171** | 30/10/2017 | 1.1.2.80.349.1012.8756 | Morocco | Africa | Northern Africa |
| **SRR6922664** | 30/10/2017 | 1.1.2.80.349.1012.8762 | Morocco | Africa | Northern Africa |
| **SRR7495741** | 05/12/2017 | 1.1.2.80.349.1012.8979 | Morocco | Africa | Northern Africa |
| **SRR7090673** | 19/04/2018 | 1.1.2.80.349.1012.9839 | Morocco | Africa | Northern Africa |
| **SRR7495456** | 01/05/2018 | 1.1.2.80.349.1012.9887 | Morocco | Africa | Northern Africa |
| **SRR8509200** | 17/01/2019 | 1.1.2.80.349.2639.12657 | Morocco | Africa | Northern Africa |
| **SRR8142771** | 18/10/2018 | 1.1.2.80.349.4328.12228 | Morocco | Africa | Northern Africa |
| **SRR7890380** | 31/08/2018 | 1.1.2.80.3560.4725.11558 | Morocco | Africa | Northern Africa |

| | | | | | |
|---|---|---|---|---|---|
| **SRR6918651** | 16/10/2017 | 1.1.2.80.728.4192.8655 | Morocco | Africa | Northern Africa |
| **SRR1963528** | 27/10/2014 | 1.1.2.80.728.792.1742 | Morocco | Africa | Northern Africa |
| **SRR1963335** | 13/11/2014 | 1.1.2.80.80.80.2035 | Morocco | Africa | Northern Africa |
| **SRR3049307** | 09/06/2014 | 1.1.2.80.833.921.1183 | Morocco | Africa | Northern Africa |
| **SRR8503748** | 20/03/2018 | 1.1.333.103.103.2119.9727 | Morocco | Africa | Northern Africa |
| **SRR5215638** | 26/07/2016 | 1.2.3.151.151.783.5031 | Morocco | Africa | Northern Africa |
| **SRR5194297** | 11/07/2016 | 1.2.3.151.362.363.4934 | Morocco | Africa | Northern Africa |
| **SRR8116993** | 16/10/2018 | 1.2.3.18.180.180.12194 | Morocco | Africa | Northern Africa |
| **SRR8839538** | 21/03/2019 | 1.2.3.18.2950.3720.12871 | Morocco | Africa | Northern Africa |
| **SRR8878999** | 26/03/2019 | 1.2.3.18.2950.3720.12888 | Morocco | Africa | Northern Africa |
| **SRR6919302** | 24/04/2017 | 1.2.3.18.2950.3720.7203 | Morocco | Africa | Northern Africa |
| **SRR6898008** | 25/10/2017 | 1.2.3.18.2950.3720.8742 | Morocco | Africa | Northern Africa |
| **SRR8717069** | 06/08/2015 | 1.1.127.233.1785.2109.3263 | Pakistan | Asia | Southern Asia |
| **SRR5220674** | 15/11/2016 | 1.1.127.233.2608.3286.6299 | Pakistan | Asia | Southern Asia |
| **SRR3286718** | 28/05/2015 | 1.1.2.147.1047.1962.2852 | Pakistan | Asia | Southern Asia |
| **SRR7516241** | 13/07/2017 | 1.1.2.147.1047.2286.7756 | Pakistan | Asia | Southern Asia |
| **SRR6898938** | 17/01/2017 | 1.1.2.147.1047.3586.6885 | Pakistan | Asia | Southern Asia |
| **SRR6897795** | 29/11/2017 | 1.1.2.147.1047.4285.8941 | Pakistan | Asia | Southern Asia |
| **SRR6324230** | 15/04/2015 | 1.1.2.147.855.951.1221 | Pakistan | Asia | Southern Asia |
| **SRR5216334** | 13/09/2016 | 1.1.2.28.2471.3082.5659 | Pakistan | Asia | Southern Asia |
| **SRR6918616** | 04/10/2016 | 1.1.2.504.2340.2914.5876 | Pakistan | Asia | Southern Asia |
| **SRR5220203** | 17/09/2015 | 1.2.3.151.151.783.2809 | Pakistan | Asia | Southern Asia |
| **SRR8711352** | 16/09/2015 | 1.1.2.1132.1880.2241.3559 | Poland | Europe | Eastern Europe |
| **SRR8385673** | 19/12/2018 | 1.1.2.12.12.12.12585 | Poland | Europe | Eastern Europe |
| **SRR8936125** | 01/04/2019 | 1.1.2.12.12.12.12910 | Poland | Europe | Eastern Europe |
| **SRR3049635** | 21/08/2014 | 1.1.2.12.12.12.2106 | Poland | Europe | Eastern Europe |
| **SRR1958144** | 10/07/2014 | 1.1.2.12.12.12.496 | Poland | Europe | Eastern Europe |
| **SRR6899359** | 31/08/2017 | 1.1.2.12.12.12.7700 | Poland | Europe | Eastern Europe |
| **SRR7465069** | 12/06/2018 | 1.1.2.12.12.12.8795 | Poland | Europe | Eastern Europe |
| **SRR6918540** | 04/04/2017 | 1.1.2.12.12.2681.7138 | Poland | Europe | Eastern Europe |
| **SRR6900483** | 25/05/2017 | 1.1.2.12.12.3778.7365 | Poland | Europe | Eastern Europe |
| **SRR6918646** | 10/10/2017 | 1.1.2.12.12.4185.8627 | Poland | Europe | Eastern Europe |
| **SRR8484201** | 29/08/2018 | 1.1.2.12.12.590.11526 | Poland | Europe | Eastern Europe |
| **SRR7997119** | 17/09/2018 | 1.1.2.12.12.590.11753 | Poland | Europe | Eastern Europe |
| **SRR8717226** | 03/09/2015 | 1.1.2.12.12.590.3413 | Poland | Europe | Eastern Europe |
| **SRR5193887** | 14/09/2016 | 1.1.2.12.12.590.3988 | Poland | Europe | Eastern Europe |
| **SRR7458149** | 04/04/2018 | 1.1.2.12.12.590.9773 | Poland | Europe | Eastern Europe |
| **SRR8293834** | 04/09/2018 | 1.1.2.12.12.590.9897 | Poland | Europe | Eastern Europe |
| **SRR6898868** | 12/09/2016 | 1.1.2.12.12.890.5626 | Poland | Europe | Eastern Europe |
| **SRR7230717** | 11/09/2015 | 1.1.2.12.421.2201.4263 | Poland | Europe | Eastern Europe |
| **SRR6922080** | 02/08/2017 | 1.1.2.12.421.3957.7825 | Poland | Europe | Eastern Europe |
| **SRR6922721** | 30/12/2015 | 1.1.2.12.421.429.4225 | Poland | Europe | Eastern Europe |
| **SRR3049013** | 16/07/2014 | 1.1.2.127.127.1273.2151 | Poland | Europe | Eastern Europe |
| **SRR8958155** | 08/04/2019 | 1.1.2.1293.2392.5478.12939 | Poland | Europe | Eastern Europe |
| **SRR5220966** | 12/11/2015 | 1.1.2.152.152.2430.4020 | Poland | Europe | Eastern Europe |
| **SRR5216195** | 27/10/2016 | 1.1.2.26.26.3220.6136 | Poland | Europe | Eastern Europe |
| **SRR1959269** | 19/09/2014 | 1.1.2.265.265.265.265 | Poland | Europe | Eastern Europe |
| **SRR6924056** | 04/09/2017 | 1.1.2.265.265.265.8264 | Poland | Europe | Eastern Europe |

| | | | | | |
|---|---|---|---|---|---|
| **SRR6900824** | 19/04/2016 | 1.1.2.28.195.195.4593 | Poland | Europe | Eastern Europe |
| **SRR5194338** | 18/09/2015 | 1.1.2.28.195.358.3453 | Poland | Europe | Eastern Europe |
| **SRR1958262** | 14/10/2014 | 1.1.2.28.195.358.360 | Poland | Europe | Eastern Europe |
| **SRR5216569** | 10/11/2015 | 1.1.2.28.195.358.4034 | Poland | Europe | Eastern Europe |
| **SRR5193497** | 15/09/2016 | 1.1.2.28.195.358.5686 | Poland | Europe | Eastern Europe |
| **SRR6900863** | 03/07/2017 | 1.1.2.28.195.3922.7693 | Poland | Europe | Eastern Europe |
| **SRR5216435** | 23/11/2015 | 1.1.2.28.2034.2460.4086 | Poland | Europe | Eastern Europe |
| **SRR6899403** | 16/06/2017 | 1.1.658.2313.3082.3923.7696 | Poland | Europe | Eastern Europe |
| **SRR3286856** | 17/06/2015 | 1.2.3.18.1691.1961.2954 | Poland | Europe | Eastern Europe |
| **SRR5216439** | 26/04/2016 | 1.2.3.18.175.1305.4621 | Poland | Europe | Eastern Europe |
| **SRR8490776** | 30/08/2018 | 1.2.3.18.175.175.11564 | Poland | Europe | Eastern Europe |
| **SRR6898818** | 26/07/2016 | 1.2.3.18.175.175.5018 | Poland | Europe | Eastern Europe |
| **SRR5216426** | 05/01/2017 | 1.2.3.18.175.175.6842 | Poland | Europe | Eastern Europe |
| **SRR7480620** | 12/06/2018 | 1.2.3.18.175.175.9937 | Poland | Europe | Eastern Europe |
| **SRR6922674** | 04/04/2017 | 1.2.3.18.175.2769.7131 | Poland | Europe | Eastern Europe |
| **SRR8503953** | 04/04/2018 | 1.2.3.18.180.180.10861 | Poland | Europe | Eastern Europe |
| **SRR5216266** | 13/09/2016 | 1.2.3.18.180.180.11246 | Poland | Europe | Eastern Europe |
| **SRR7998226** | 13/09/2018 | 1.2.3.18.180.180.11711 | Poland | Europe | Eastern Europe |
| **SRR6900941** | 25/09/2017 | 1.2.3.18.180.5091.11123 | Poland | Europe | Eastern Europe |
| **SRR6900699** | 15/09/2016 | 1.2.3.18.180.5120.10980 | Poland | Europe | Eastern Europe |
| **SRR6922695** | 08/08/2017 | 1.2.3.18.180.5238.11170 | Poland | Europe | Eastern Europe |
| **SRR7867150** | 31/08/2018 | 1.2.3.18.180.5402.11564 | Poland | Europe | Eastern Europe |
| **SRR4063713** | 11/09/2015 | 1.2.3.18.1895.2261.4287 | Poland | Europe | Eastern Europe |
| **SRR8382004** | 17/07/2018 | 1.2.3.18.3626.4855.10443 | Poland | Europe | Eastern Europe |
| **SRR7286763** | 01/09/2015 | 1.2.3.18.3698.5090.11222 | Poland | Europe | Eastern Europe |
| **SRR3286629** | 08/05/2015 | 1.2.3.18.3698.5100.10870 | Poland | Europe | Eastern Europe |
| **SRR6924135** | 24/08/2016 | 1.2.3.18.3698.5100.10882 | Poland | Europe | Eastern Europe |
| **SRR3286944** | 30/06/2015 | 1.2.3.18.458.472.3037 | Poland | Europe | Eastern Europe |
| **SRR5193031** | 08/07/2015 | 1.2.3.18.458.472.3082 | Poland | Europe | Eastern Europe |
| **SRR5194231** | 08/07/2015 | 1.2.3.18.458.472.3110 | Poland | Europe | Eastern Europe |
| **SRR6922676** | 07/07/2016 | 1.2.3.18.458.472.4947 | Poland | Europe | Eastern Europe |
| **SRR4063704** | 04/01/2016 | 1.2.3.18.779.854.4238 | Poland | Europe | Eastern Europe |
| **SRR6922743** | 07/07/2017 | 1.2.3.18.816.896.7772 | Poland | Europe | Eastern Europe |
| **SRR4063712** | 28/09/2015 | 1.2.3.18.860.2248.3590 | Poland | Europe | Eastern Europe |
| **SRR7506914** | 26/06/2018 | 1.2.3.421.3577.4756.10216 | Poland | Europe | Eastern Europe |
| **SRR6919850** | 13/06/2017 | 1.1.2.1037.3028.3853.7552 | Portugal | Europe | Southern Europe |
| **SRR6900457** | 14/11/2017 | 1.1.2.1037.3028.3853.8874 | Portugal | Europe | Southern Europe |
| **SRR6920473** | 26/06/2017 | 1.1.2.1076.3075.3909.7650 | Portugal | Europe | Southern Europe |
| **SRR7538811** | 05/07/2018 | 1.1.2.12.421.4553.10336 | Portugal | Europe | Southern Europe |
| **SRR5215656** | 02/06/2016 | 1.1.2.1283.2203.2706.4759 | Portugal | Europe | Southern Europe |
| **SRR5631728** | 26/04/2017 | 1.1.2.1283.2203.3729.7225 | Portugal | Europe | Southern Europe |
| **SRR1969548** | 27/08/2014 | 1.1.2.131.456.1040.1377 | Portugal | Europe | Southern Europe |
| **SRR5220918** | 10/11/2015 | 1.1.2.1326.373.2419.3997 | Portugal | Europe | Southern Europe |
| **SRR7122587** | 29/10/2015 | 1.1.2.28.195.2087.3210 | Portugal | Europe | Southern Europe |
| **SRR1965366** | 01/10/2014 | 1.2.3.151.151.151.798 | Portugal | Europe | Southern Europe |
| **SRR1969684** | 09/09/2014 | 1.2.3.151.151.151.974 | Portugal | Europe | Southern Europe |
| **SRR7879555** | 16/07/2018 | 1.2.3.151.151.783.10410 | Portugal | Europe | Southern Europe |
| **SRR8711755** | 28/09/2015 | 1.2.3.151.151.783.2809 | Portugal | Europe | Southern Europe |

| | | | | | |
|---|---|---|---|---|---|
| **SRR5216418** | 18/10/2016 | 1.2.3.18.175.175.6033 | Portugal | Europe | Southern Europe |
| **SRR8137458** | 19/10/2018 | 1.2.3.18.180.180.10985 | Portugal | Europe | Southern Europe |
| **SRR6899303** | 18/07/2017 | 1.2.3.18.180.5093.10912 | Portugal | Europe | Southern Europe |
| **SRR7298410** | 18/04/2018 | 1.2.3.18.180.5093.11146 | Portugal | Europe | Southern Europe |
| **SRR7417282** | 05/06/2018 | 1.2.3.18.3101.3966.10085 | Portugal | Europe | Southern Europe |
| **SRR8293561** | 10/09/2018 | 1.2.3.18.3101.3966.11667 | Portugal | Europe | Southern Europe |
| **SRR5583250** | 27/04/2017 | 1.2.3.18.359.360.7159 | Portugal | Europe | Southern Europe |
| **SRR6897968** | 15/06/2017 | 1.2.3.18.359.360.7553 | Portugal | Europe | Southern Europe |
| **SRR6897974** | 07/09/2017 | 1.2.3.18.359.360.8308 | Portugal | Europe | Southern Europe |
| **SRR5216116** | 15/09/2016 | 1.2.3.203.2182.2671.4922 | Portugal | Europe | Southern Europe |
| **SRR5193912** | 24/08/2016 | 1.2.3.203.2182.2671.5203 | Portugal | Europe | Southern Europe |
| **SRR6901013** | 14/07/2017 | 1.21.92.248.248.4017.8095 | Portugal | Europe | Southern Europe |
| **SRR6900823** | 07/09/2017 | 1.5.159.280.280.280.8319 | Portugal | Europe | Southern Europe |
| **SRR5194118** | 24/06/2016 | 1.5.321.1298.2231.2760.4903 | Portugal | Europe | Southern Europe |
| **SRR5215793** | 20/09/2016 | 1.5.321.1298.2231.2760.5726 | Portugal | Europe | Southern Europe |
| **SRR7310322** | 27/08/2015 | 1.1.2.1107.1835.2175.3401 | Russian federation | Europe | Eastern Europe |
| **SRR7469039** | 30/09/2015 | 1.1.2.1151.1908.2278.3666 | Russian federation | Europe | Eastern Europe |
| **SRR1968370** | 24/06/2014 | 1.1.2.2073.834.922.1185 | Russian federation | Europe | Eastern Europe |
| **SRR6920538** | 11/09/2017 | 1.1.2.2342.3167.4090.8340 | Russian federation | Europe | Eastern Europe |
| **SRR7961932** | 13/09/2018 | 1.1.2.2562.3886.5436.11715 | Russian federation | Europe | Eastern Europe |
| **SRR1958142** | 07/11/2014 | 1.1.2.28.1428.1647.2265 | Russian federation | Europe | Eastern Europe |
| **SRR8380066** | 18/07/2018 | 1.1.2.28.1428.4865.10465 | Russian federation | Europe | Eastern Europe |
| **SRR1969359** | 04/07/2014 | 1.1.2.51.51.51.51 | Russian federation | Europe | Eastern Europe |
| **SRR5216384** | 09/08/2016 | 1.1.2.78.2277.2826.5108 | Russian federation | Europe | Eastern Europe |
| **SRR5220667** | 21/06/2016 | 1.1.320.1296.2225.2746.4865 | Russian federation | Europe | Eastern Europe |
| **SRR7850546** | 20/07/2018 | 1.1.320.1296.3634.4875.10506 | Russian federation | Europe | Eastern Europe |
| **SRR7351414** | 23/09/2015 | 1.2.270.1145.1898.2264.3626 | Russian federation | Europe | Eastern Europe |
| **SRR6901127** | 18/10/2016 | 1.1.2.1326.544.2643.6036 | Saudi arabia | Asia | Western Asia |
| **SRR5633338** | 25/04/2017 | 1.1.2.1326.544.3723.7212 | Saudi arabia | Asia | Western Asia |
| **SRR1958427** | 29/08/2014 | 1.1.2.1326.544.571.930 | Saudi arabia | Asia | Western Asia |
| **SRR7468936** | 06/02/2018 | 1.1.2.147.1047.4448.9361 | Saudi arabia | Asia | Western Asia |
| **SRR8137285** | 13/09/2018 | 1.1.2.274.274.4281.11714 | Saudi arabia | Asia | Western Asia |
| **SRR7456980** | 27/04/2018 | 1.1.2.274.274.4281.9878 | Saudi arabia | Asia | Western Asia |
| **SRR7458633** | 23/01/2018 | 1.1.2.274.3362.4435.9302 | Saudi arabia | Asia | Western Asia |
| **SRR5632175** | 06/02/2017 | 1.1.2.28.195.195.6922 | Saudi arabia | Asia | Western Asia |
| **SRR5193675** | 28/04/2016 | 1.1.2.28.535.2659.4633 | Saudi arabia | Asia | Western Asia |
| **SRR6898859** | 21/09/2017 | 1.1.2.359.983.1104.8457 | Saudi arabia | Asia | Western Asia |
| **SRR1968728** | 09/03/2015 | 1.1.2.504.1236.1407.1908 | Saudi arabia | Asia | Western Asia |
| **SRR7439550** | 30/09/2015 | 1.1.2.504.1236.2272.3652 | Saudi arabia | Asia | Western Asia |
| **SRR6919308** | 15/11/2016 | 1.1.2.504.2607.3284.6290 | Saudi | Asia | Western Asia |

| | | | | | |
|---|---|---|---|---|---|
| | | | arabia | | |
| **SRR6924059** | 19/09/2017 | 1.1.2.504.3191.4127.8447 | Saudi arabia | Asia | Western Asia |
| **SRR8833187** | 19/03/2019 | 1.1.2.504.4111.5863.12858 | Saudi arabia | Asia | Western Asia |
| **SRR7349374** | 15/06/2015 | 1.1.2.504.633.1999.2935 | Saudi arabia | Asia | Western Asia |
| **SRR3286572** | 15/04/2015 | 1.1.2.504.967.1440.1955 | Saudi arabia | Asia | Western Asia |
| **SRR1961770** | 31/12/2014 | 1.1.2.504.967.1440.1970 | Saudi arabia | Asia | Western Asia |
| **SRR6900611** | 04/05/2017 | 1.1.2.504.967.1440.7248 | Saudi arabia | Asia | Western Asia |
| **SRR6919740** | 29/11/2017 | 1.1.2.504.967.4284.8936 | Saudi arabia | Asia | Western Asia |
| **SRR8397154** | 18/12/2018 | 1.2.3.18.180.180.12575 | Saudi arabia | Asia | Western Asia |
| **SRR8724678** | 27/02/2019 | 1.2.3.18.365.1120.12788 | Saudi arabia | Asia | Western Asia |
| **SRR1967526** | 07/05/2014 | 1.3.154.272.272.272.272 | Saudi arabia | Asia | Western Asia |
| **SRR8172826** | 23/10/2018 | 1.1.185.1138.3091.3940.12298 | Singapore | Asia | South-eastern Asia |
| **SRR1957918** | 23/09/2014 | 1.1.2.157.157.157.157 | Singapore | Asia | South-eastern Asia |
| **SRR5220466** | 27/04/2015 | 1.1.2.206.1660.1916.2716 | Singapore | Asia | South-eastern Asia |
| **SRR5584871** | 03/05/2017 | 1.1.2.206.1980.3725.7216 | Singapore | Asia | South-eastern Asia |
| **SRR1966294** | 05/11/2014 | 1.1.2.206.227.227.227 | Singapore | Asia | South-eastern Asia |
| **SRR6924082** | 28/04/2017 | 1.1.2.206.2466.3074.7236 | Singapore | Asia | South-eastern Asia |
| **SRR1963443** | 01/12/2014 | 1.1.2.206.508.679.830 | Singapore | Asia | South-eastern Asia |
| **SRR1958005** | 29/08/2014 | 1.1.2.206.758.829.1051 | Singapore | Asia | South-eastern Asia |
| **SRR7358374** | 11/05/2018 | 1.1.2.2306.3558.4722.9945 | Singapore | Asia | South-eastern Asia |
| **SRR1958385** | 25/09/2014 | 1.1.2.28.195.1454.1973 | Singapore | Asia | South-eastern Asia |
| **SRR6920162** | 04/12/2017 | 1.1.2.28.1998.4301.8984 | Singapore | Asia | South-eastern Asia |
| **SRR5220867** | 09/11/2015 | 1.1.2.28.2010.2417.3984 | Singapore | Asia | South-eastern Asia |
| **SRR8553928** | 25/07/2017 | 1.1.2.28.3100.3965.7869 | Singapore | Asia | South-eastern Asia |
| **SRR3285441** | 17/12/2015 | 1.1.2.28.354.354.4201 | Singapore | Asia | South-eastern Asia |
| **SRR7251107** | 17/04/2015 | 1.1.2.4.1654.1908.2685 | Singapore | Asia | South-eastern Asia |
| **SRR8509052** | 16/04/2018 | 1.1.2.4.3525.4662.9783 | Singapore | Asia | South-eastern Asia |
| **SRR1960144** | 05/01/2015 | 1.1.2.4.4.1165.1553 | Singapore | Asia | South-eastern Asia |
| **SRR7457938** | 08/04/2015 | 1.1.2.4.4.1909.2701 | Singapore | Asia | South-eastern Asia |
| **SRR8288221** | 26/11/2018 | 1.1.2.4.4.5719.12478 | Singapore | Asia | South-eastern Asia |
| **SRR5215737** | 01/12/2015 | 1.1.2.7.2046.2476.4136 | Singapore | Asia | South-eastern Asia |
| **SRR7841326** | 17/07/2018 | 1.1.2.7.3625.4854.10442 | Singapore | Asia | South-eastern Asia |
| **SRR1965938** | 16/03/2015 | 1.1.208.623.859.955.1226 | Singapore | Asia | South-eastern Asia |
| **SRR8503781** | 17/04/2018 | 1.1.226.1700.3540.4688.9831 | Singapore | Asia | South-eastern Asia |
| **SRR4063710** | 17/02/2016 | 1.2.3.18.2123.2584.4434 | South africa | Africa | Sub-Saharan Africa |
| **SRR8509103** | 26/04/2018 | 1.2.3.18.3538.4681.9817 | South africa | Africa | Sub-Saharan Africa |
| **SRR5584680** | 10/01/2017 | 1.2.3.18.365.1120.6862 | South africa | Africa | Sub-Saharan Africa |
| **SRR5583841** | 24/02/2017 | 1.2.3.18.365.1120.7023 | South africa | Africa | Sub-Saharan Africa |
| **SRR8508560** | 08/03/2018 | 1.2.3.18.365.3656.9676 | South africa | Africa | Sub-Saharan Africa |
| **SRR7359619** | 08/03/2018 | 1.2.3.18.365.3656.9698 | South africa | Africa | Sub-Saharan Africa |
| **SRR6900962** | 09/08/2017 | 1.2.3.18.365.3946.7788 | South africa | Africa | Sub-Saharan Africa |
| **SRR1965486** | 27/06/2014 | 1.2.3.18.365.476.521 | South africa | Africa | Sub-Saharan Africa |

| | | | | | |
|---|---|---|---|---|---|
| **SRR7892248** | 11/09/2018 | 1.2.3.18.365.5431.11689 | South africa | Africa | Sub-Saharan Africa |
| **SRR8490750** | 27/09/2018 | 3.4.8.1691.4008.5624.12043 | South africa | Africa | Sub-Saharan Africa |
| **SRR3284689** | 11/11/2015 | 1.1.2.1065.1746.2051.3585 | Spain | Europe | Southern Europe |
| **SRR5220516** | 27/11/2015 | 1.1.2.1065.1746.2051.4107 | Spain | Europe | Southern Europe |
| **SRR5193307** | 24/07/2015 | 1.1.2.1072.1759.2066.3148 | Spain | Europe | Southern Europe |
| **SRR6918330** | 15/06/2016 | 1.1.2.1072.2219.2735.4837 | Spain | Europe | Southern Europe |
| **SRR8142765** | 18/10/2018 | 1.1.2.12.421.429.12206 | Spain | Europe | Southern Europe |
| **SRR1957812** | 10/09/2014 | 1.1.2.131.131.131.131 | Spain | Europe | Southern Europe |
| **SRR1968831** | 01/10/2014 | 1.1.2.131.456.1040.1441 | Spain | Europe | Southern Europe |
| **SRR3286812** | 10/06/2015 | 1.1.2.131.456.1040.2926 | Spain | Europe | Southern Europe |
| **SRR6901064** | 02/10/2017 | 1.1.2.131.456.1040.5082 | Spain | Europe | Southern Europe |
| **SRR5193699** | 04/10/2016 | 1.1.2.131.456.1040.5862 | Spain | Europe | Southern Europe |
| **SRR6900363** | 10/01/2017 | 1.1.2.131.456.1040.6850 | Spain | Europe | Southern Europe |
| **SRR3285361** | 29/09/2015 | 1.1.2.131.456.2311.3745 | Spain | Europe | Southern Europe |
| **SRR6919114** | 17/02/2016 | 1.1.2.131.456.470.4429 | Spain | Europe | Southern Europe |
| **SRR5220873** | 19/07/2016 | 1.1.2.131.456.470.5013 | Spain | Europe | Southern Europe |
| **SRR5216146** | 29/07/2016 | 1.1.2.131.456.470.5063 | Spain | Europe | Southern Europe |
| **SRR6919287** | 21/09/2017 | 1.1.2.131.456.470.8459 | Spain | Europe | Southern Europe |
| **SRR5193600** | 28/10/2015 | 1.1.2.1326.373.2393.3918 | Spain | Europe | Southern Europe |
| **SRR5193208** | 03/10/2016 | 1.1.2.1326.373.2419.5828 | Spain | Europe | Southern Europe |
| **SRR1968969** | 05/11/2014 | 1.1.2.138.138.138.832 | Spain | Europe | Southern Europe |
| **SRR1970135** | 17/09/2014 | 1.1.2.166.166.166.166 | Spain | Europe | Southern Europe |
| **SRR6900452** | 22/08/2017 | 1.1.2.167.167.167.8145 | Spain | Europe | Southern Europe |
| **SRR6919718** | 19/09/2017 | 1.1.2.167.167.167.8437 | Spain | Europe | Southern Europe |
| **SRR5583221** | 07/03/2017 | 1.1.2.1709.2485.3678.7059 | Spain | Europe | Southern Europe |
| **SRR7456732** | 28/02/2018 | 1.1.2.1709.2485.4623.9707 | Spain | Europe | Southern Europe |
| **SRR8183061** | 30/10/2018 | 1.1.2.2066.411.3654.12339 | Spain | Europe | Southern Europe |
| **SRR8249759** | 09/11/2018 | 1.1.2.2066.411.3654.12419 | Spain | Europe | Southern Europe |
| **SRR7286907** | 14/08/2015 | 1.1.2.2066.411.3654.3341 | Spain | Europe | Southern Europe |
| **SRR5194262** | 11/07/2016 | 1.1.2.2066.411.3654.4941 | Spain | Europe | Southern Europe |
| **SRR5216399** | 24/08/2016 | 1.1.2.2066.411.3654.5212 | Spain | Europe | Southern Europe |
| **SRR5193844** | 02/09/2016 | 1.1.2.2066.411.3654.5531 | Spain | Europe | Southern Europe |
| **SRR6918647** | 01/11/2016 | 1.1.2.2066.411.3654.5910 | Spain | Europe | Southern Europe |
| **SRR6920022** | 05/10/2017 | 1.1.2.2066.411.3654.8540 | Spain | Europe | Southern Europe |
| **SRR6898875** | 16/10/2017 | 1.1.2.2066.411.3654.8658 | Spain | Europe | Southern Europe |
| **SRR3285352** | 20/07/2015 | 1.1.2.208.208.1938.2875 | Spain | Europe | Southern Europe |
| **SRR5220932** | 14/07/2015 | 1.1.2.208.208.1938.3120 | Spain | Europe | Southern Europe |
| **SRR6919747** | 10/05/2016 | 1.1.2.208.208.1938.4667 | Spain | Europe | Southern Europe |
| **SRR5193498** | 26/07/2016 | 1.1.2.208.208.1938.5033 | Spain | Europe | Southern Europe |
| **SRR1968443** | 21/10/2014 | 1.1.2.208.208.208.208 | Spain | Europe | Southern Europe |
| **SRR1967416** | 04/11/2014 | 1.1.2.208.208.430.460 | Spain | Europe | Southern Europe |
| **SRR5220531** | 06/05/2015 | 1.1.2.208.208.560.2770 | Spain | Europe | Southern Europe |
| **SRR8724686** | 28/07/2015 | 1.1.2.208.208.560.3216 | Spain | Europe | Southern Europe |
| **SRR3323006** | 15/10/2015 | 1.1.2.208.208.560.3831 | Spain | Europe | Southern Europe |
| **SRR7286819** | 26/10/2015 | 1.1.2.208.208.560.3955 | Spain | Europe | Southern Europe |
| **SRR5216298** | 25/05/2016 | 1.1.2.208.208.560.4706 | Spain | Europe | Southern Europe |
| **SRR6900843** | 27/07/2016 | 1.1.2.208.208.560.5046 | Spain | Europe | Southern Europe |

| | | | | | |
|---|---|---|---|---|---|
| **SRR3048852** | 08/10/2014 | 1.1.2.208.208.560.650 | Spain | Europe | Southern Europe |
| **SRR1965182** | 04/11/2014 | 1.1.2.208.208.786.981 | Spain | Europe | Southern Europe |
| **SRR1958398** | 15/10/2014 | 1.1.2.208.614.655.2208 | Spain | Europe | Southern Europe |
| **SRR6922127** | 17/10/2017 | 1.1.2.230.230.2078.8677 | Spain | Europe | Southern Europe |
| **SRR1965681** | 07/10/2014 | 1.1.2.230.230.230.230 | Spain | Europe | Southern Europe |
| **SRR8893102** | 02/04/2019 | 1.1.2.2309.3074.3907.12912 | Spain | Europe | Southern Europe |
| **SRR8936128** | 01/04/2019 | 1.1.2.2309.3074.3907.12928 | Spain | Europe | Southern Europe |
| **SRR8568744** | 23/06/2017 | 1.1.2.2309.3074.3907.7644 | Spain | Europe | Southern Europe |
| **SRR6900049** | 03/07/2017 | 1.1.2.2309.3074.3907.7652 | Spain | Europe | Southern Europe |
| **SRR7495449** | 13/07/2017 | 1.1.2.2309.3074.3907.7682 | Spain | Europe | Southern Europe |
| **SRR6900693** | 10/07/2017 | 1.1.2.2309.3074.3907.7740 | Spain | Europe | Southern Europe |
| **SRR6919840** | 10/08/2017 | 1.1.2.2309.3074.3907.7947 | Spain | Europe | Southern Europe |
| **SRR6900274** | 25/08/2017 | 1.1.2.2309.3074.3907.8191 | Spain | Europe | Southern Europe |
| **SRR6899328** | 29/09/2017 | 1.1.2.2309.3074.3907.8552 | Spain | Europe | Southern Europe |
| **SRR6920599** | 03/11/2017 | 1.1.2.2309.3074.3907.8782 | Spain | Europe | Southern Europe |
| **SRR6924142** | 10/11/2017 | 1.1.2.2309.3074.3907.8866 | Spain | Europe | Southern Europe |
| **SRR7890377** | 07/02/2018 | 1.1.2.2309.3074.3907.9369 | Spain | Europe | Southern Europe |
| **SRR8304788** | 05/10/2018 | 1.1.2.2546.3840.5332.12105 | Spain | Europe | Southern Europe |
| **SRR5220321** | 26/07/2016 | 1.1.2.274.2260.2798.5022 | Spain | Europe | Southern Europe |
| **SRR8490787** | 03/12/2018 | 1.1.2.28.195.195.12511 | Spain | Europe | Southern Europe |
| **SRR5632731** | 17/01/2017 | 1.1.2.28.195.195.6883 | Spain | Europe | Southern Europe |
| **SRR8325468** | 03/12/2018 | 1.1.2.28.195.195.8125 | Spain | Europe | Southern Europe |
| **SRR7866996** | 10/07/2018 | 1.1.2.28.195.457.10404 | Spain | Europe | Southern Europe |
| **SRR1965746** | 12/05/2014 | 1.1.2.31.31.1283.1729 | Spain | Europe | Southern Europe |
| **SRR3049564** | 05/06/2014 | 1.1.2.31.31.31.31 | Spain | Europe | Southern Europe |
| **SRR5193120** | 21/07/2015 | 1.1.2.63.1758.2065.3147 | Spain | Europe | Southern Europe |
| **SRR7468939** | 11/09/2015 | 1.1.2.63.1758.2065.3439 | Spain | Europe | Southern Europe |
| **SRR5220462** | 23/08/2016 | 1.1.2.63.1758.2065.5240 | Spain | Europe | Southern Europe |
| **SRR7828454** | 30/08/2018 | 1.1.2.63.1758.5385.11577 | Spain | Europe | Southern Europe |
| **SRR1967033** | 08/08/2014 | 1.1.2.63.63.1421.1926 | Spain | Europe | Southern Europe |
| **SRR7892158** | 11/09/2018 | 1.1.2.63.63.3346.11691 | Spain | Europe | Southern Europe |
| **SRR1967985** | 01/10/2014 | 1.1.2.63.63.603.2053 | Spain | Europe | Southern Europe |
| **SRR1965385** | 06/10/2014 | 1.1.2.63.63.63.1618 | Spain | Europe | Southern Europe |
| **SRR1966311** | 30/09/2014 | 1.1.2.63.63.63.2017 | Spain | Europe | Southern Europe |
| **SRR1966738** | 07/10/2014 | 1.1.2.63.63.755.940 | Spain | Europe | Southern Europe |
| **SRR1966199** | 23/05/2014 | 1.1.2.80.100.100.100 | Spain | Europe | Southern Europe |
| **SRR5215482** | 02/08/2016 | 1.1.2.80.100.100.5095 | Spain | Europe | Southern Europe |
| **SRR1968401** | 15/08/2014 | 1.1.2.80.100.100.565 | Spain | Europe | Southern Europe |
| **SRR6899509** | 08/08/2017 | 1.1.2.80.100.100.7880 | Spain | Europe | Southern Europe |
| **SRR6921916** | 15/09/2017 | 1.1.2.80.100.100.8421 | Spain | Europe | Southern Europe |
| **SRR5216277** | 05/05/2016 | 1.1.2.80.1121.1266.4641 | Spain | Europe | Southern Europe |
| **SRR5216238** | 06/05/2016 | 1.1.2.80.1121.1266.4643 | Spain | Europe | Southern Europe |
| **SRR8300686** | 31/08/2018 | 1.1.2.80.1373.1574.11564 | Spain | Europe | Southern Europe |
| **SRR8724642** | 31/07/2015 | 1.1.2.80.1373.1574.2152 | Spain | Europe | Southern Europe |
| **SRR5193426** | 08/07/2015 | 1.1.2.80.1373.1574.3125 | Spain | Europe | Southern Europe |
| **SRR7297935** | 17/07/2015 | 1.1.2.80.1373.1574.3138 | Spain | Europe | Southern Europe |
| **SRR8724952** | 16/07/2015 | 1.1.2.80.1373.1574.3139 | Spain | Europe | Southern Europe |
| **SRR6922565** | 21/06/2016 | 1.1.2.80.1373.1574.4036 | Spain | Europe | Southern Europe |

| | | | | | |
|---|---|---|---|---|---|
| **SRR8436106** | 09/07/2018 | 1.1.2.80.1373.4841.10377 | Spain | Europe | Southern Europe |
| **SRR7187243** | 22/09/2015 | 1.1.2.80.1389.1597.3645 | Spain | Europe | Southern Europe |
| **SRR1965204** | 24/02/2015 | 1.1.2.80.164.164.1160 | Spain | Europe | Southern Europe |
| **SRR1968590** | 17/07/2014 | 1.1.2.80.164.164.164 | Spain | Europe | Southern Europe |
| **SRR8380683** | 17/07/2018 | 1.1.2.80.1903.2273.10474 | Spain | Europe | Southern Europe |
| **SRR8368704** | 31/07/2018 | 1.1.2.80.1903.2273.10616 | Spain | Europe | Southern Europe |
| **SRR8106760** | 21/09/2018 | 1.1.2.80.1903.2273.11918 | Spain | Europe | Southern Europe |
| **SRR5220821** | 28/09/2015 | 1.1.2.80.1903.2273.3654 | Spain | Europe | Southern Europe |
| **SRR5193038** | 13/09/2016 | 1.1.2.80.1903.2273.5636 | Spain | Europe | Southern Europe |
| **SRR6898444** | 03/10/2017 | 1.1.2.80.1903.2273.7912 | Spain | Europe | Southern Europe |
| **SRR6899399** | 16/10/2017 | 1.1.2.80.1903.2273.8653 | Spain | Europe | Southern Europe |
| **SRR1957755** | 10/09/2014 | 1.1.2.80.207.207.1672 | Spain | Europe | Southern Europe |
| **SRR1965813** | 14/10/2014 | 1.1.2.80.207.207.207 | Spain | Europe | Southern Europe |
| **SRR1966754** | 11/09/2014 | 1.1.2.80.207.207.2261 | Spain | Europe | Southern Europe |
| **SRR7962240** | 13/09/2018 | 1.1.2.80.453.467.11709 | Spain | Europe | Southern Europe |
| **SRR1968857** | 10/11/2014 | 1.1.2.80.453.467.1182 | Spain | Europe | Southern Europe |
| **SRR1965671** | 04/11/2014 | 1.1.2.80.453.467.1923 | Spain | Europe | Southern Europe |
| **SRR5193652** | 24/05/2016 | 1.1.2.80.453.467.3463 | Spain | Europe | Southern Europe |
| **SRR6899256** | 03/08/2017 | 1.1.2.80.453.467.7814 | Spain | Europe | Southern Europe |
| **SRR7892210** | 01/08/2017 | 1.1.2.80.453.467.7898 | Spain | Europe | Southern Europe |
| **SRR6922666** | 19/07/2017 | 1.1.2.80.453.467.8021 | Spain | Europe | Southern Europe |
| **SRR6919161** | 14/09/2017 | 1.1.2.80.453.467.8382 | Spain | Europe | Southern Europe |
| **SRR7359062** | 01/10/2015 | 1.1.2.80.635.680.3656 | Spain | Europe | Southern Europe |
| **SRR1966215** | 16/09/2014 | 1.1.2.83.83.826.1187 | Spain | Europe | Southern Europe |
| **SRR1967838** | 13/08/2014 | 1.1.2.83.83.83.925 | Spain | Europe | Southern Europe |
| **SRR7410281** | 11/06/2018 | 1.1.228.1033.1681.1946.10114 | Spain | Europe | Southern Europe |
| **SRR3585376** | 11/05/2015 | 1.1.228.1033.1681.1946.2785 | Spain | Europe | Southern Europe |
| **SRR5220694** | 08/10/2015 | 1.1.228.1033.1681.1946.3797 | Spain | Europe | Southern Europe |
| **SRR6924068** | 02/08/2017 | 1.1.228.1033.1681.1946.8091 | Spain | Europe | Southern Europe |
| **SRR6900198** | 08/09/2017 | 1.1.228.1033.1681.1946.8144 | Spain | Europe | Southern Europe |
| **SRR6898843** | 07/09/2017 | 1.1.228.1033.1681.1946.8303 | Spain | Europe | Southern Europe |
| **SRR7495715** | 25/01/2018 | 1.1.228.1033.1681.1946.9305 | Spain | Europe | Southern Europe |
| **SRR7416262** | 06/07/2017 | 1.1.228.1033.1681.3935.7736 | Spain | Europe | Southern Europe |
| **SRR6900719** | 01/08/2017 | 1.1.228.1033.1681.3935.7827 | Spain | Europe | Southern Europe |
| **SRR7369167** | 10/11/2015 | 1.1.234.1045.1708.2437.4033 | Spain | Europe | Southern Europe |
| **SRR7842555** | 06/07/2018 | 1.1.63.97.1353.2016.10376 | Spain | Europe | Southern Europe |
| **SRR6897174** | 19/06/2017 | 1.1.63.97.1353.2016.7625 | Spain | Europe | Southern Europe |
| **SRR6922728** | 08/08/2017 | 1.1.63.97.1353.2016.7866 | Spain | Europe | Southern Europe |
| **SRR6922744** | 09/08/2017 | 1.1.63.97.1353.2016.7903 | Spain | Europe | Southern Europe |
| **SRR6900206** | 04/09/2017 | 1.1.63.97.1353.2016.8279 | Spain | Europe | Southern Europe |
| **SRR6918886** | 07/09/2017 | 1.1.63.97.1353.2016.8300 | Spain | Europe | Southern Europe |
| **SRR6901145** | 28/09/2017 | 1.1.63.97.1353.2016.8528 | Spain | Europe | Southern Europe |
| **SRR6900370** | 02/08/2017 | 1.1.63.97.1353.4020.8099 | Spain | Europe | Southern Europe |
| **SRR8131514** | 13/09/2018 | 1.12.724.2561.3885.5435.1171<br>3 | Spain | Europe | Southern Europe |
| **SRR7204302** | 19/08/2015 | 1.2.3.1089.1805.2138.3323 | Spain | Europe | Southern Europe |
| **SRR8118258** | 20/09/2018 | 1.2.3.151.151.151.11439 | Spain | Europe | Southern Europe |
| **SRR8936154** | 02/04/2019 | 1.2.3.151.151.151.11652 | Spain | Europe | Southern Europe |

| | | | | | |
|---|---|---|---|---|---|
| **SRR8738298** | 05/03/2019 | 1.2.3.151.151.151.12810 | Spain | Europe | Southern Europe |
| **SRR1965138** | 29/07/2014 | 1.2.3.151.151.151.1701 | Spain | Europe | Southern Europe |
| **SRR8735162** | 24/07/2015 | 1.2.3.151.151.151.3167 | Spain | Europe | Southern Europe |
| **SRR6900541** | 22/08/2016 | 1.2.3.151.151.151.5149 | Spain | Europe | Southern Europe |
| **SRR5193678** | 23/08/2016 | 1.2.3.151.151.151.5187 | Spain | Europe | Southern Europe |
| **SRR1967574** | 31/07/2014 | 1.2.3.151.151.151.798 | Spain | Europe | Southern Europe |
| **SRR6922497** | 07/09/2017 | 1.2.3.151.151.151.8155 | Spain | Europe | Southern Europe |
| **SRR6919162** | 18/07/2017 | 1.2.3.151.151.718.7750 | Spain | Europe | Southern Europe |
| **SRR7443860** | 19/06/2018 | 1.2.3.151.151.783.10149 | Spain | Europe | Southern Europe |
| **SRR7892257** | 11/09/2018 | 1.2.3.151.151.783.11684 | Spain | Europe | Southern Europe |
| **SRR7903045** | 12/09/2018 | 1.2.3.151.151.783.11694 | Spain | Europe | Southern Europe |
| **SRR8117004** | 15/10/2018 | 1.2.3.151.151.783.12059 | Spain | Europe | Southern Europe |
| **SRR8137420** | 10/10/2018 | 1.2.3.151.151.783.12124 | Spain | Europe | Southern Europe |
| **SRR8137363** | 17/10/2018 | 1.2.3.151.151.783.12212 | Spain | Europe | Southern Europe |
| **SRR8149424** | 19/10/2018 | 1.2.3.151.151.783.12269 | Spain | Europe | Southern Europe |
| **SRR7458571** | 14/09/2015 | 1.2.3.151.151.783.2809 | Spain | Europe | Southern Europe |
| **SRR5220942** | 16/10/2015 | 1.2.3.151.151.783.3026 | Spain | Europe | Southern Europe |
| **SRR5220325** | 29/09/2015 | 1.2.3.151.151.783.3681 | Spain | Europe | Southern Europe |
| **SRR5193493** | 06/10/2015 | 1.2.3.151.151.783.3739 | Spain | Europe | Southern Europe |
| **SRR5216121** | 07/07/2016 | 1.2.3.151.151.783.4927 | Spain | Europe | Southern Europe |
| **SRR8553930** | 01/08/2017 | 1.2.3.151.151.783.7812 | Spain | Europe | Southern Europe |
| **SRR6919750** | 18/07/2017 | 1.2.3.151.151.783.7828 | Spain | Europe | Southern Europe |
| **SRR6900688** | 17/07/2017 | 1.2.3.151.151.783.7913 | Spain | Europe | Southern Europe |
| **SRR7511922** | 13/08/2015 | 1.2.3.151.1800.2130.3307 | Spain | Europe | Southern Europe |
| **SRR1967215** | 29/07/2014 | 1.2.3.151.356.372.1078 | Spain | Europe | Southern Europe |
| **SRR1967921** | 03/07/2014 | 1.2.3.151.356.372.1202 | Spain | Europe | Southern Europe |
| **SRR1968355** | 28/10/2014 | 1.2.3.151.356.372.1400 | Spain | Europe | Southern Europe |
| **SRR1968620** | 01/07/2014 | 1.2.3.151.356.372.1849 | Spain | Europe | Southern Europe |
| **SRR5220506** | 29/09/2015 | 1.2.3.151.356.372.3670 | Spain | Europe | Southern Europe |
| **SRR1970316** | 16/09/2014 | 1.2.3.151.356.372.379 | Spain | Europe | Southern Europe |
| **SRR5216512** | 14/06/2016 | 1.2.3.151.356.372.4825 | Spain | Europe | Southern Europe |
| **SRR5193241** | 29/07/2016 | 1.2.3.151.356.372.5061 | Spain | Europe | Southern Europe |
| **SRR1968851** | 23/05/2014 | 1.2.3.151.356.372.641 | Spain | Europe | Southern Europe |
| **SRR1966120** | 31/07/2014 | 1.2.3.151.356.372.647 | Spain | Europe | Southern Europe |
| **SRR5585115** | 31/01/2017 | 1.2.3.151.356.372.6879 | Spain | Europe | Southern Europe |
| **SRR3049234** | 07/05/2014 | 1.2.3.151.356.372.889 | Spain | Europe | Southern Europe |
| **SRR3049690** | 08/10/2014 | 1.2.3.151.356.423.957 | Spain | Europe | Southern Europe |
| **SRR7475508** | 21/06/2018 | 1.2.3.151.356.4685.10165 | Spain | Europe | Southern Europe |
| **SRR8485047** | 12/10/2018 | 1.2.3.151.356.4685.12162 | Spain | Europe | Southern Europe |
| **SRR8116981** | 15/10/2018 | 1.2.3.151.356.4685.12202 | Spain | Europe | Southern Europe |
| **SRR3284735** | 08/07/2015 | 1.2.3.151.362.2038.3062 | Spain | Europe | Southern Europe |
| **SRR7884511** | 06/09/2018 | 1.2.3.151.362.363.11662 | Spain | Europe | Southern Europe |
| **SRR3312312** | 23/07/2015 | 1.2.3.151.362.363.3032 | Spain | Europe | Southern Europe |
| **SRR3319057** | 17/07/2015 | 1.2.3.151.362.363.3095 | Spain | Europe | Southern Europe |
| **SRR3319060** | 03/09/2015 | 1.2.3.151.362.363.3417 | Spain | Europe | Southern Europe |
| **SRR5216213** | 31/08/2016 | 1.2.3.151.362.363.5270 | Spain | Europe | Southern Europe |
| **SRR5193388** | 20/09/2016 | 1.2.3.151.362.363.5678 | Spain | Europe | Southern Europe |
| **SRR5220546** | 20/09/2016 | 1.2.3.151.362.363.5718 | Spain | Europe | Southern Europe |

| | | | | | |
|---|---|---|---|---|---|
| **SRR6900944** | 23/09/2016 | 1.2.3.151.362.363.5755 | Spain | Europe | Southern Europe |
| **SRR5194266** | 26/10/2016 | 1.2.3.151.362.363.6077 | Spain | Europe | Southern Europe |
| **SRR1968164** | 22/01/2015 | 1.2.3.151.378.381.388 | Spain | Europe | Southern Europe |
| **SRR1958347** | 09/10/2014 | 1.2.3.151.552.580.2303 | Spain | Europe | Southern Europe |
| **SRR1965796** | 19/05/2014 | 1.2.3.151.759.830.1054 | Spain | Europe | Southern Europe |
| **SRR1967754** | 13/05/2014 | 1.2.3.151.759.830.1077 | Spain | Europe | Southern Europe |
| **SRR3048603** | 20/01/2015 | 1.2.3.151.783.859.1096 | Spain | Europe | Southern Europe |
| **SRR1966899** | 05/06/2014 | 1.2.3.151.839.932.1196 | Spain | Europe | Southern Europe |
| **SRR8272559** | 10/09/2018 | 1.2.3.18.1686.1956.11657 | Spain | Europe | Southern Europe |
| **SRR3321876** | 29/05/2015 | 1.2.3.18.1686.1956.2829 | Spain | Europe | Southern Europe |
| **SRR7890409** | 22/05/2018 | 1.2.3.18.175.175.10000 | Spain | Europe | Southern Europe |
| **SRR8380676** | 17/07/2018 | 1.2.3.18.175.175.10445 | Spain | Europe | Southern Europe |
| **SRR6922603** | 29/09/2017 | 1.2.3.18.175.175.7619 | Spain | Europe | Southern Europe |
| **SRR6920114** | 04/07/2017 | 1.2.3.18.180.180.10797 | Spain | Europe | Southern Europe |
| **SRR6897887** | 04/05/2017 | 1.2.3.18.180.180.10820 | Spain | Europe | Southern Europe |
| **SRR6899257** | 26/06/2017 | 1.2.3.18.180.180.10888 | Spain | Europe | Southern Europe |
| **SRR6900919** | 05/09/2017 | 1.2.3.18.180.180.10899 | Spain | Europe | Southern Europe |
| **SRR8492378** | 19/06/2018 | 1.2.3.18.180.180.10909 | Spain | Europe | Southern Europe |
| **SRR6922473** | 06/09/2017 | 1.2.3.18.180.180.10916 | Spain | Europe | Southern Europe |
| **SRR6919256** | 30/08/2017 | 1.2.3.18.180.180.10929 | Spain | Europe | Southern Europe |
| **SRR8490656** | 28/06/2018 | 1.2.3.18.180.180.10942 | Spain | Europe | Southern Europe |
| **SRR8509387** | 23/02/2018 | 1.2.3.18.180.180.10944 | Spain | Europe | Southern Europe |
| **SRR6900867** | 07/09/2017 | 1.2.3.18.180.180.10961 | Spain | Europe | Southern Europe |
| **SRR7867006** | 16/08/2018 | 1.2.3.18.180.180.10985 | Spain | Europe | Southern Europe |
| **SRR7850565** | 23/07/2018 | 1.2.3.18.180.180.10987 | Spain | Europe | Southern Europe |
| **SRR6900660** | 06/09/2017 | 1.2.3.18.180.180.10998 | Spain | Europe | Southern Europe |
| **SRR6900760** | 11/10/2017 | 1.2.3.18.180.180.11021 | Spain | Europe | Southern Europe |
| **SRR7841531** | 17/07/2018 | 1.2.3.18.180.180.11037 | Spain | Europe | Southern Europe |
| **SRR6920443** | 07/09/2017 | 1.2.3.18.180.180.11047 | Spain | Europe | Southern Europe |
| **SRR7842532** | 09/07/2018 | 1.2.3.18.180.180.11053 | Spain | Europe | Southern Europe |
| **SRR6897896** | 30/08/2017 | 1.2.3.18.180.180.11056 | Spain | Europe | Southern Europe |
| **SRR6924093** | 29/08/2017 | 1.2.3.18.180.180.11082 | Spain | Europe | Southern Europe |
| **SRR6897884** | 29/08/2017 | 1.2.3.18.180.180.11156 | Spain | Europe | Southern Europe |
| **SRR6919804** | 30/08/2017 | 1.2.3.18.180.180.11185 | Spain | Europe | Southern Europe |
| **SRR7480136** | 23/06/2017 | 1.2.3.18.180.180.11205 | Spain | Europe | Southern Europe |
| **SRR7867207** | 08/08/2018 | 1.2.3.18.180.180.11379 | Spain | Europe | Southern Europe |
| **SRR7879524** | 21/08/2018 | 1.2.3.18.180.180.11401 | Spain | Europe | Southern Europe |
| **SRR8350312** | 13/08/2018 | 1.2.3.18.180.180.11411 | Spain | Europe | Southern Europe |
| **SRR8087146** | 01/10/2018 | 1.2.3.18.180.180.11599 | Spain | Europe | Southern Europe |
| **SRR7910408** | 13/09/2018 | 1.2.3.18.180.180.11600 | Spain | Europe | Southern Europe |
| **SRR8106751** | 27/09/2018 | 1.2.3.18.180.180.11748 | Spain | Europe | Southern Europe |
| **SRR8114896** | 20/09/2018 | 1.2.3.18.180.180.11801 | Spain | Europe | Southern Europe |
| **SRR7998244** | 24/09/2018 | 1.2.3.18.180.180.11839 | Spain | Europe | Southern Europe |
| **SRR7997046** | 21/09/2018 | 1.2.3.18.180.180.11959 | Spain | Europe | Southern Europe |
| **SRR8084338** | 02/10/2018 | 1.2.3.18.180.180.12056 | Spain | Europe | Southern Europe |
| **SRR8117048** | 03/10/2018 | 1.2.3.18.180.180.12116 | Spain | Europe | Southern Europe |
| **SRR8084258** | 09/10/2018 | 1.2.3.18.180.180.12121 | Spain | Europe | Southern Europe |
| **SRR8114914** | 11/10/2018 | 1.2.3.18.180.180.12178 | Spain | Europe | Southern Europe |

| | | | | | |
|---|---|---|---|---|---|
| **SRR8204806** | 07/11/2018 | 1.2.3.18.180.180.12375 | Spain | Europe | Southern Europe |
| **SRR8272657** | 08/11/2018 | 1.2.3.18.180.180.12401 | Spain | Europe | Southern Europe |
| **SRR8258092** | 09/11/2018 | 1.2.3.18.180.180.12416 | Spain | Europe | Southern Europe |
| **SRR8637893** | 31/01/2019 | 1.2.3.18.180.180.12704 | Spain | Europe | Southern Europe |
| **SRR8706107** | 25/02/2019 | 1.2.3.18.180.180.12791 | Spain | Europe | Southern Europe |
| **SRR8943116** | 02/04/2019 | 1.2.3.18.180.180.12909 | Spain | Europe | Southern Europe |
| **SRR6900610** | 21/09/2017 | 1.2.3.18.180.5092.10792 | Spain | Europe | Southern Europe |
| **SRR6920142** | 24/07/2017 | 1.2.3.18.180.5092.10808 | Spain | Europe | Southern Europe |
| **SRR6924077** | 13/09/2017 | 1.2.3.18.180.5092.10971 | Spain | Europe | Southern Europe |
| **SRR6901185** | 22/11/2017 | 1.2.3.18.3228.4197.8679 | Spain | Europe | Southern Europe |
| **SRR8509253** | 25/06/2018 | 1.2.3.18.353.353.10251 | Spain | Europe | Southern Europe |
| **SRR6924045** | 20/09/2017 | 1.2.3.18.359.360.8440 | Spain | Europe | Southern Europe |
| **SRR7850472** | 16/07/2018 | 1.2.3.18.3648.4900.10574 | Spain | Europe | Southern Europe |
| **SRR6918866** | 04/10/2017 | 1.2.3.18.382.2750.8592 | Spain | Europe | Southern Europe |
| **SRR5193953** | 07/10/2015 | 1.2.3.18.536.1947.2789 | Spain | Europe | Southern Europe |
| **SRR5220308** | 22/07/2015 | 1.2.3.18.536.1947.3184 | Spain | Europe | Southern Europe |
| **SRR6900460** | 03/10/2016 | 1.2.3.18.536.1947.5758 | Spain | Europe | Southern Europe |
| **SRR6918558** | 04/07/2017 | 1.2.3.18.536.1947.7701 | Spain | Europe | Southern Europe |
| **SRR8367065** | 11/12/2018 | 1.2.3.18.536.5744.12546 | Spain | Europe | Southern Europe |
| **SRR6898474** | 21/07/2017 | 1.2.3.2326.3120.4005.8054 | Spain | Europe | Southern Europe |
| **SRR5215542** | 23/08/2016 | 1.21.438.1592.2319.2889.5257 | Spain | Europe | Southern Europe |
| **SRR3049116** | 25/04/2014 | 1.5.159.280.280.280.280 | Spain | Europe | Southern Europe |
| **SRR5216495** | 06/08/2015 | 1.5.159.280.280.280.3258 | Spain | Europe | Southern Europe |
| **SRR6897936** | 30/05/2017 | 1.5.159.280.280.280.7377 | Spain | Europe | Southern Europe |
| **SRR5216332** | 01/09/2016 | 1.5.201.507.2358.2936.5345 | Spain | Europe | Southern Europe |
| **SRR6920138** | 21/09/2017 | 1.5.201.507.2358.4140.8479 | Spain | Europe | Southern Europe |
| **SRR6900131** | 03/05/2017 | 1.5.201.507.2954.3727.7220 | Spain | Europe | Southern Europe |
| **SRR7962274** | 17/09/2018 | 1.5.321.1298.3897.5453.11769 | Spain | Europe | Southern Europe |
| **SRR5220876** | 14/10/2015 | 1.5.72.115.115.2306.3833 | Spain | Europe | Southern Europe |
| **SRR7416369** | 12/12/2017 | 1.5.72.115.115.2656.6825 | Spain | Europe | Southern Europe |
| **SRR6922076** | 21/12/2017 | 1.5.72.115.115.2656.9053 | Spain | Europe | Southern Europe |
| **SRR6919989** | 16/08/2017 | 1.5.72.115.115.3133.8143 | Spain | Europe | Southern Europe |
| **SRR8117025** | 03/10/2018 | 1.75.332.2646.4024.5657.12184 | Spain | Europe | Southern Europe |
| **SRR5216123** | 24/05/2016 | 3.4.314.1278.2195.2693.4712 | Spain | Europe | Southern Europe |
| **SRR8172828** | 23/10/2018 | 3.4.49.2334.3142.4050.12295 | Spain | Europe | Southern Europe |
| **SRR6921960** | 22/08/2017 | 3.4.49.2334.3142.4050.8171 | Spain | Europe | Southern Europe |
| **SRR5193937** | 05/10/2015 | 3.4.8.14.14.2302.3727 | Spain | Europe | Southern Europe |
| **SRR6919024** | 01/09/2016 | 3.4.8.236.236.236.5330 | Spain | Europe | Southern Europe |
| **SRR1970029** | 28/08/2014 | 31.17.71.114.114.114.114 | Spain | Europe | Southern Europe |
| **SRR5757203** | 03/12/2015 | 31.48.261.1201.2041.2469.4114 | Spain | Europe | Southern Europe |
| **SRR5194003** | 04/10/2016 | 1.1.2.1140.1891.2257.5864 | Sri lanka | Asia | Southern Asia |
| **SRR7257133** | 14/09/2015 | 1.1.2.1326.544.2234.3544 | Sri lanka | Asia | Southern Asia |
| **SRR1969095** | 27/01/2015 | 1.1.2.178.178.1626.2223 | Sri lanka | Asia | Southern Asia |
| **SRR5193455** | 08/09/2015 | 1.1.2.178.1855.2205.4265 | Sri lanka | Asia | Southern Asia |
| **SRR8448461** | 04/01/2019 | 1.1.2.178.1855.4562.12609 | Sri lanka | Asia | Southern Asia |
| **SRR8490843** | 11/10/2018 | 1.1.2.178.3157.4073.12159 | Sri lanka | Asia | Southern Asia |
| **SRR8503807** | 08/06/2018 | 1.1.2.178.3588.4768.10100 | Sri lanka | Asia | Southern Asia |

| | | | | | |
|---|---|---|---|---|---|
| **SRR8648339** | 11/02/2019 | 1.1.2.178.4093.5822.12741 | Sri lanka | Asia | Southern Asia |
| **SRR3049702** | 30/04/2014 | 1.1.2.284.1119.1264.1697 | Sri lanka | Asia | Southern Asia |
| **SRR7187263** | 26/06/2015 | 1.1.2.284.2575.3234.6161 | Sri lanka | Asia | Southern Asia |
| **SRR1963349** | 07/01/2015 | 1.1.2.550.735.799.999 | Sri lanka | Asia | Southern Asia |
| **SRR3286613** | 08/04/2015 | 1.1.2.601.817.897.1146 | Sri lanka | Asia | Southern Asia |
| **SRR3284822** | 30/09/2015 | 1.1.2.67.1905.2275.3660 | Sri lanka | Asia | Southern Asia |
| **SRR6922720** | 31/12/2015 | 1.1.2.67.67.2517.4254 | Sri lanka | Asia | Southern Asia |
| **SRR5216144** | 01/12/2016 | 1.1.2.67.67.3304.6369 | Sri lanka | Asia | Southern Asia |
| **SRR7468879** | 20/11/2015 | 1.1.2.67.67.676.3916 | Sri lanka | Asia | Southern Asia |
| **SRR1969673** | 17/03/2015 | 1.1.2.67.67.676.827 | Sri lanka | Asia | Southern Asia |
| **SRR6918645** | 16/10/2017 | 1.30.188.2246.3226.4194.8665 | Sri lanka | Asia | Southern Asia |
| **SRR6924143** | 20/09/2016 | 1.1.2.28.2487.3109.5723 | China | Asia | Eastern Asia |
| **SRR6922527** | 12/09/2017 | 1.1.2.28.3171.4094.8354 | China | Asia | Eastern Asia |
| **SRR6901010** | 09/11/2017 | 1.1.2.28.3254.4250.8845 | China | Asia | Eastern Asia |
| **SRR5216253** | 17/11/2015 | 1.1.2.28.2025.2444.4050 | Tanzania | Africa | Sub-Saharan Africa |
| **SRR6899381** | 07/08/2017 | 1.1.2.28.2025.3956.7822 | Tanzania | Africa | Sub-Saharan Africa |
| **SRR1969721** | 02/09/2014 | 1.1.2.28.372.374.1179 | Tanzania | Africa | Sub-Saharan Africa |
| **SRR1969926** | 01/10/2014 | 1.1.2.28.372.374.381 | Tanzania | Africa | Sub-Saharan Africa |
| **SRR1966175** | 05/03/2015 | 1.1.2.28.372.374.409 | Tanzania | Africa | Sub-Saharan Africa |
| **SRR5216387** | 08/07/2016 | 1.1.2.28.372.374.4952 | Tanzania | Africa | Sub-Saharan Africa |
| **SRR1967629** | 17/09/2014 | 1.1.2.28.372.374.597 | Tanzania | Africa | Sub-Saharan Africa |
| **SRR7474377** | 09/05/2018 | 1.1.2.28.372.374.9926 | Tanzania | Africa | Sub-Saharan Africa |
| **SRR1969741** | 01/08/2014 | 1.1.2.28.372.583.692 | Tanzania | Africa | Sub-Saharan Africa |
| **SRR8182954** | 30/10/2018 | 1.1.2.28.4033.5683.12335 | Tanzania | Africa | Sub-Saharan Africa |
| **SRR6919812** | 27/09/2017 | 1.1.63.97.1353.2016.8525 | Tanzania | Africa | Sub-Saharan Africa |
| **SRR4063702** | 03/11/2015 | 1.2.3.18.365.2425.4012 | Tanzania | Africa | Sub-Saharan Africa |
| **SRR5216534** | 22/09/2016 | 1.1.2.1030.1856.2581.5810 | Thailand | Asia | South-eastern Asia |
| **SRR8185543** | 30/10/2018 | 1.1.2.1030.1856.2862.12341 | Thailand | Asia | South-eastern Asia |
| **SRR7410323** | 13/12/2016 | 1.1.2.1030.1856.2862.6454 | Thailand | Asia | South-eastern Asia |
| **SRR5631345** | 18/04/2017 | 1.1.2.1030.1856.2862.7176 | Thailand | Asia | South-eastern Asia |
| **SRR8514482** | 15/02/2018 | 1.1.2.1030.1856.2862.9421 | Thailand | Asia | South-eastern Asia |
| **SRR7480357** | 22/05/2018 | 1.1.2.1030.1856.4622.9995 | Thailand | Asia | South-eastern Asia |
| **SRR7867303** | 24/08/2018 | 1.1.2.1030.1856.5377.11505 | Thailand | Asia | South-eastern Asia |
| **SRR5216367** | 30/03/2016 | 1.1.2.1030.2152.2625.4543 | Thailand | Asia | South-eastern Asia |
| **SRR6897172** | 26/06/2017 | 1.1.2.1030.3080.3916.7677 | Thailand | Asia | South-eastern Asia |
| **SRR7402356** | 07/06/2018 | 1.1.2.1030.3581.4760.10081 | Thailand | Asia | South-eastern Asia |
| **SRR8717172** | 30/07/2015 | 1.1.2.1076.1769.2089.3213 | Thailand | Asia | South-eastern Asia |
| **SRR5633181** | 24/03/2017 | 1.1.2.1076.1769.3696.7114 | Thailand | Asia | South-eastern Asia |
| **SRR8437384** | 10/07/2018 | 1.1.2.1124.3595.4779.10319 | Thailand | Asia | South-eastern Asia |
| **SRR5220104** | 11/01/2017 | 1.1.2.116.2763.3579.6856 | Thailand | Asia | South-eastern Asia |
| **SRR6899000** | 23/12/2015 | 1.1.2.1164.2072.2507.4235 | Thailand | Asia | South-eastern Asia |
| **SRR5584725** | 10/05/2017 | 1.1.2.1164.2072.3743.7260 | Thailand | Asia | South-eastern Asia |
| **SRR7866795** | 01/08/2018 | 1.1.2.1164.2072.4965.10632 | Thailand | Asia | South-eastern Asia |
| **SRR8548900** | 23/01/2019 | 1.1.2.1164.2961.5794.12682 | Thailand | Asia | South-eastern Asia |
| **SRR6900612** | 13/12/2017 | 1.1.2.1890.3289.4322.9031 | Thailand | Asia | South-eastern Asia |
| **SRR3322968** | 27/10/2015 | 1.1.2.206.1997.2394.3921 | Thailand | Asia | South-eastern Asia |
| **SRR7866787** | 29/08/2018 | 1.1.2.215.215.215.11531 | Thailand | Asia | South-eastern Asia |
| **SRR1966361** | 04/11/2014 | 1.1.2.215.215.215.215 | Thailand | Asia | South-eastern Asia |

| | | | | | |
|---|---|---|---|---|---|
| **SRR6922134** | 23/05/2017 | 1.1.2.2259.2974.3767.7325 | Thailand | Asia | South-eastern Asia |
| **SRR7873996** | 15/08/2018 | 1.1.2.2336.3145.4054.11452 | Thailand | Asia | South-eastern Asia |
| **SRR1968780** | 12/11/2014 | 1.1.2.239.239.239.239 | Thailand | Asia | South-eastern Asia |
| **SRR1959386** | 09/12/2014 | 1.1.2.268.268.268.268 | Thailand | Asia | South-eastern Asia |
| **SRR1966976** | 09/03/2015 | 1.1.2.28.1463.1689.2328 | Thailand | Asia | South-eastern Asia |
| **SRR6900934** | 19/01/2016 | 1.1.2.28.1463.2552.4352 | Thailand | Asia | South-eastern Asia |
| **SRR5584696** | 06/03/2017 | 1.1.2.28.2255.3673.7038 | Thailand | Asia | South-eastern Asia |
| **SRR8116983** | 15/10/2018 | 1.1.2.28.4025.5658.12186 | Thailand | Asia | South-eastern Asia |
| **SRR5216208** | 23/12/2016 | 1.1.2.28.429.439.6601 | Thailand | Asia | South-eastern Asia |
| **SRR3286635** | 08/04/2015 | 1.1.2.28.562.593.706 | Thailand | Asia | South-eastern Asia |
| **SRR5193533** | 25/05/2016 | 1.1.2.34.2192.2686.4701 | Thailand | Asia | South-eastern Asia |
| **SRR6898826** | 28/07/2016 | 1.1.2.34.2264.2808.5053 | Thailand | Asia | South-eastern Asia |
| **SRR8492371** | 30/05/2018 | 1.1.2.34.2296.2850.10050 | Thailand | Asia | South-eastern Asia |
| **SRR8369254** | 27/07/2018 | 1.1.2.34.2296.2850.10548 | Thailand | Asia | South-eastern Asia |
| **SRR8508935** | 18/01/2019 | 1.1.2.34.2296.2850.12658 | Thailand | Asia | South-eastern Asia |
| **SRR8585165** | 29/01/2019 | 1.1.2.34.2296.2850.12696 | Thailand | Asia | South-eastern Asia |
| **SRR6900157** | 02/09/2016 | 1.1.2.34.2296.2850.5372 | Thailand | Asia | South-eastern Asia |
| **SRR6900183** | 30/03/2017 | 1.1.2.34.2296.2850.7124 | Thailand | Asia | South-eastern Asia |
| **SRR6919758** | 13/09/2017 | 1.1.2.34.2296.2850.8370 | Thailand | Asia | South-eastern Asia |
| **SRR7902697** | 07/02/2018 | 1.1.2.34.2296.2850.9375 | Thailand | Asia | South-eastern Asia |
| **SRR6897989** | 30/08/2017 | 1.1.2.34.2296.2850.9611 | Thailand | Asia | South-eastern Asia |
| **SRR7251549** | 20/03/2018 | 1.1.2.34.2296.2850.9724 | Thailand | Asia | South-eastern Asia |
| **SRR7416107** | 03/05/2018 | 1.1.2.34.2296.2850.9912 | Thailand | Asia | South-eastern Asia |
| **SRR7414869** | 09/02/2018 | 1.1.2.34.2296.4532.9482 | Thailand | Asia | South-eastern Asia |
| **SRR8509030** | 16/03/2018 | 1.1.2.34.2296.4629.9715 | Thailand | Asia | South-eastern Asia |
| **SRR8509152** | 24/04/2018 | 1.1.2.34.2296.4698.9861 | Thailand | Asia | South-eastern Asia |
| **SRR7522942** | 02/07/2018 | 1.1.2.34.2296.4829.10313 | Thailand | Asia | South-eastern Asia |
| **SRR8585003** | 30/01/2019 | 1.1.2.34.2296.5803.12697 | Thailand | Asia | South-eastern Asia |
| **SRR8704695** | 21/02/2019 | 1.1.2.34.2296.5839.12782 | Thailand | Asia | South-eastern Asia |
| **SRR8772191** | 11/03/2019 | 1.1.2.34.2296.5854.12834 | Thailand | Asia | South-eastern Asia |
| **SRR8820638** | 19/03/2019 | 1.1.2.34.2296.5862.12856 | Thailand | Asia | South-eastern Asia |
| **SRR3048902** | 03/06/2014 | 1.1.2.34.34.34.34 | Thailand | Asia | South-eastern Asia |
| **SRR8738375** | 04/03/2019 | 1.1.2.34.4102.5844.12805 | Thailand | Asia | South-eastern Asia |
| **SRR8292208** | 27/11/2018 | 1.1.2.34.687.745.12480 | Thailand | Asia | South-eastern Asia |
| **SRR1966964** | 17/02/2015 | 1.1.2.34.687.745.927 | Thailand | Asia | South-eastern Asia |
| **SRR6900189** | 06/12/2017 | 1.1.2.4.3283.4310.8995 | Thailand | Asia | South-eastern Asia |
| **SRR1968135** | 31/03/2015 | 1.1.2.403.443.455.497 | Thailand | Asia | South-eastern Asia |
| **SRR1968352** | 03/02/2015 | 1.1.2.635.882.984.1270 | Thailand | Asia | South-eastern Asia |
| **SRR3286941** | 25/06/2015 | 1.1.2.849.1723.2014.2974 | Thailand | Asia | South-eastern Asia |
| **SRR6918613** | 01/12/2017 | 1.1.2.849.3271.4295.8968 | Thailand | Asia | South-eastern Asia |
| **SRR7284412** | 27/04/2018 | 1.1.2.849.3545.4703.9874 | Thailand | Asia | South-eastern Asia |
| **SRR6924091** | 13/11/2017 | 1.1.200.491.2282.2832.8863 | Thailand | Asia | South-eastern Asia |
| **SRR6918601** | 13/01/2016 | 1.1.292.1231.2092.2540.4319 | Thailand | Asia | South-eastern Asia |
| **SRR5631948** | 17/01/2017 | 1.1.292.1231.2839.3587.6889 | Thailand | Asia | South-eastern Asia |
| **SRR6920103** | 21/09/2017 | 1.1.292.1231.3194.4134.8470 | Thailand | Asia | South-eastern Asia |
| **SRR5216186** | 27/06/2016 | 1.1.322.1300.2235.2764.4915 | Thailand | Asia | South-eastern Asia |
| **SRR5216293** | 23/08/2016 | 1.1.437.1591.2316.2886.5244 | Thailand | Asia | South-eastern Asia |
| **SRR7344646** | 18/04/2018 | 1.1.685.2420.3350.4419.9835 | Thailand | Asia | South-eastern Asia |

| | | | | | |
|---|---|---|---|---|---|
| **SRR5193422** | 06/09/2016 | 1.2.3.151.151.151.4817 | Thailand | Asia | South-eastern Asia |
| **SRR8839368** | 20/03/2019 | 4.142.773.2677.4114.5868.128<br>72 | Thailand | Asia | South-eastern Asia |
| **SRR6898863** | 23/09/2016 | 4.45.257.1106.2490.3112.5753 | Thailand | Asia | South-eastern Asia |
| **SRR8490651** | 04/07/2018 | 1.1.2.1124.3595.4779.10319 | Tunisia | Africa | Northern Africa |
| **SRR7867202** | 16/07/2018 | 1.1.2.1124.3595.4779.10412 | Tunisia | Africa | Northern Africa |
| **SRR7873865** | 12/07/2018 | 1.1.2.1124.3595.4779.10416 | Tunisia | Africa | Northern Africa |
| **SRR1966356** | 27/08/2014 | 1.1.2.119.119.119.119 | Tunisia | Africa | Northern Africa |
| **SRR1958671** | 24/09/2014 | 1.1.2.28.1054.1186.1584 | Tunisia | Africa | Northern Africa |
| **SRR1969660** | 08/08/2014 | 1.1.2.28.1284.1467.1989 | Tunisia | Africa | Northern Africa |
| **SRR8638526** | 04/02/2019 | 1.1.2.28.419.444.12708 | Tunisia | Africa | Northern Africa |
| **SRR1966919** | 12/05/2014 | 1.1.2.28.419.444.1884 | Tunisia | Africa | Northern Africa |
| **SRR1966481** | 01/08/2014 | 1.1.2.28.419.444.1979 | Tunisia | Africa | Northern Africa |
| **SRR7450794** | 03/09/2014 | 1.1.2.28.419.444.754 | Tunisia | Africa | Northern Africa |
| **SRR1957974** | 14/10/2014 | 1.1.2.28.419.630.756 | Tunisia | Africa | Northern Africa |
| **SRR1960326** | 13/11/2014 | 1.1.2.28.452.466.1703 | Tunisia | Africa | Northern Africa |
| **SRR1966470** | 17/09/2014 | 1.1.2.28.725.1265.1699 | Tunisia | Africa | Northern Africa |
| **SRR1968093** | 10/10/2014 | 1.1.2.28.725.788.983 | Tunisia | Africa | Northern Africa |
| **SRR3049331** | 02/10/2014 | 1.1.2.28.877.1790.2492 | Tunisia | Africa | Northern Africa |
| **SRR1967218** | 07/10/2014 | 1.1.2.28.877.976.1256 | Tunisia | Africa | Northern Africa |
| **SRR1958251** | 24/10/2014 | 1.1.2.512.656.707.864 | Tunisia | Africa | Northern Africa |
| **SRR1958118** | 08/10/2014 | 1.1.2.96.196.196.1231 | Tunisia | Africa | Northern Africa |
| **SRR1958113** | 23/09/2014 | 1.1.2.96.196.196.1435 | Tunisia | Africa | Northern Africa |
| **SRR1966765** | 08/08/2014 | 1.1.2.96.670.1860.2601 | Tunisia | Africa | Northern Africa |
| **SRR3048760** | 03/10/2014 | 1.1.2.96.670.723.895 | Tunisia | Africa | Northern Africa |
| **SRR5216513** | 08/06/2015 | 1.1.2.96.96.1983.2902 | Tunisia | Africa | Northern Africa |
| **SRR1967572** | 04/08/2014 | 1.1.2.96.96.96.96 | Tunisia | Africa | Northern Africa |
| **SRR6922682** | 15/11/2016 | 1.2.3.151.356.372.6293 | Tunisia | Africa | Northern Africa |
| **SRR1966532** | 29/09/2014 | 1.2.3.18.364.1101.1467 | Tunisia | Africa | Northern Africa |
| **SRR1960319** | 22/09/2014 | 1.2.3.18.364.1101.1983 | Tunisia | Africa | Northern Africa |
| **SRR1966718** | 21/10/2014 | 1.2.3.18.364.410.426 | Tunisia | Africa | Northern Africa |
| **SRR1965041** | 08/10/2014 | 1.2.3.18.369.371.1306 | Tunisia | Africa | Northern Africa |
| **SRR1960237** | 13/11/2014 | 1.2.3.18.369.371.2242 | Tunisia | Africa | Northern Africa |
| **SRR1967928** | 18/09/2014 | 1.2.3.18.369.371.430 | Tunisia | Africa | Northern Africa |
| **SRR1965117** | 19/08/2014 | 1.2.3.18.369.371.498 | Tunisia | Africa | Northern Africa |
| **SRR1965128** | 16/09/2014 | 1.2.3.18.369.513.578 | Tunisia | Africa | Northern Africa |
| **SRR1966022** | 30/09/2014 | 1.2.3.18.369.517.584 | Tunisia | Africa | Northern Africa |
| **SRR7873883** | 08/08/2018 | 1.2.3.18.369.5321.11363 | Tunisia | Africa | Northern Africa |
| **SRR7842451** | 16/08/2018 | 1.2.3.18.369.5361.11455 | Tunisia | Africa | Northern Africa |
| **SRR1969229** | 24/07/2014 | 1.2.3.18.38.38.393 | Tunisia | Africa | Northern Africa |
| **SRR3049544** | 16/07/2014 | 1.2.3.18.432.443.1736 | Tunisia | Africa | Northern Africa |
| **SRR1966071** | 04/07/2014 | 1.2.3.18.432.742.1199 | Tunisia | Africa | Northern Africa |
| **SRR1968440** | 02/07/2014 | 1.2.3.18.432.742.1244 | Tunisia | Africa | Northern Africa |
| **SRR1966534** | 15/08/2014 | 1.2.3.18.516.537.1648 | Tunisia | Africa | Northern Africa |
| **SRR1967243** | 29/10/2014 | 1.2.3.18.516.537.619 | Tunisia | Africa | Northern Africa |
| **SRR1969881** | 06/11/2014 | 1.2.3.18.990.1113.1485 | Tunisia | Africa | Northern Africa |
| **SRR8149186** | 24/10/2018 | 1.1.2.1161.1926.5676.12277 | Turkey | Asia | Western Asia |
| **SRR7450844** | 20/06/2018 | 1.1.2.12.12.590.10082 | Turkey | Asia | Western Asia |

| | | | | | |
|---|---|---|---|---|---|
| **SRR5216339** | 04/07/2016 | 1.1.2.1256.2146.2617.4902 | Turkey | Asia | Western Asia |
| **SRR5220537** | 18/08/2016 | 1.1.2.1256.2146.2617.5159 | Turkey | Asia | Western Asia |
| **SRR6900734** | 23/08/2016 | 1.1.2.1256.2146.2617.5199 | Turkey | Asia | Western Asia |
| **SRR6900464** | 25/08/2016 | 1.1.2.1256.2146.2617.5214 | Turkey | Asia | Western Asia |
| **SRR5194206** | 03/11/2016 | 1.1.2.1256.2146.2617.6245 | Turkey | Asia | Western Asia |
| **SRR6921985** | 17/10/2017 | 1.1.2.1256.2146.2617.8672 | Turkey | Asia | Western Asia |
| **SRR6924156** | 29/11/2017 | 1.1.2.1256.2146.2617.8938 | Turkey | Asia | Western Asia |
| **SRR1957978** | 25/09/2014 | 1.1.2.1326.1041.1173.1565 | Turkey | Asia | Western Asia |
| **SRR5220554** | 29/09/2015 | 1.1.2.1326.1904.2274.3659 | Turkey | Asia | Western Asia |
| **SRR7450853** | 30/05/2018 | 1.1.2.1326.2022.2792.10015 | Turkey | Asia | Western Asia |
| **SRR7475341** | 30/05/2018 | 1.1.2.1326.2022.2792.10037 | Turkey | Asia | Western Asia |
| **SRR7867109** | 14/08/2018 | 1.1.2.1326.2022.2792.11405 | Turkey | Asia | Western Asia |
| **SRR7841477** | 17/08/2018 | 1.1.2.1326.2022.2792.11451 | Turkey | Asia | Western Asia |
| **SRR7879669** | 20/08/2018 | 1.1.2.1326.2022.2792.11469 | Turkey | Asia | Western Asia |
| **SRR8325497** | 18/09/2018 | 1.1.2.1326.2022.2792.11527 | Turkey | Asia | Western Asia |
| **SRR8492436** | 29/08/2018 | 1.1.2.1326.2022.2792.11538 | Turkey | Asia | Western Asia |
| **SRR7842632** | 05/09/2018 | 1.1.2.1326.2022.2792.11637 | Turkey | Asia | Western Asia |
| **SRR7997191** | 10/09/2018 | 1.1.2.1326.2022.2792.11693 | Turkey | Asia | Western Asia |
| **SRR7998246** | 21/09/2018 | 1.1.2.1326.2022.2792.11924 | Turkey | Asia | Western Asia |
| **SRR8201805** | 06/11/2018 | 1.1.2.1326.2022.2792.12381 | Turkey | Asia | Western Asia |
| **SRR8204052** | 07/11/2018 | 1.1.2.1326.2022.2792.12389 | Turkey | Asia | Western Asia |
| **SRR5193910** | 19/07/2016 | 1.1.2.1326.2022.2792.5004 | Turkey | Asia | Western Asia |
| **SRR6898104** | 26/07/2017 | 1.1.2.1326.2022.3958.7830 | Turkey | Asia | Western Asia |
| **SRR5193853** | 13/07/2016 | 1.1.2.1326.2247.2781.4976 | Turkey | Asia | Western Asia |
| **SRR6898878** | 28/07/2016 | 1.1.2.1326.2247.2781.5037 | Turkey | Asia | Western Asia |
| **SRR6921971** | 31/08/2016 | 1.1.2.1326.2247.2781.5258 | Turkey | Asia | Western Asia |
| **SRR5216494** | 08/11/2016 | 1.1.2.1326.2247.2781.6260 | Turkey | Asia | Western Asia |
| **SRR6921949** | 03/05/2017 | 1.1.2.1326.2247.2781.7232 | Turkey | Asia | Western Asia |
| **SRR5215760** | 21/07/2016 | 1.1.2.1326.393.2719.5000 | Turkey | Asia | Western Asia |
| **SRR1958381** | 01/09/2014 | 1.1.2.1326.447.461.1171 | Turkey | Asia | Western Asia |
| **SRR1966579** | 30/07/2014 | 1.1.2.1326.447.461.1600 | Turkey | Asia | Western Asia |
| **SRR1958367** | 22/09/2014 | 1.1.2.1326.447.461.1691 | Turkey | Asia | Western Asia |
| **SRR1967004** | 27/08/2014 | 1.1.2.1326.447.461.2049 | Turkey | Asia | Western Asia |
| **SRR1969533** | 16/09/2014 | 1.1.2.1326.447.461.504 | Turkey | Asia | Western Asia |
| **SRR6919238** | 31/08/2016 | 1.1.2.1326.447.461.5262 | Turkey | Asia | Western Asia |
| **SRR6922739** | 06/10/2016 | 1.1.2.1326.447.461.5512 | Turkey | Asia | Western Asia |
| **SRR5194196** | 13/09/2016 | 1.1.2.1326.447.461.5650 | Turkey | Asia | Western Asia |
| **SRR5193223** | 26/09/2016 | 1.1.2.1326.447.461.5738 | Turkey | Asia | Western Asia |
| **SRR8661097** | 12/10/2016 | 1.1.2.1326.447.461.5942 | Turkey | Asia | Western Asia |
| **SRR6901173** | 13/06/2017 | 1.1.2.1326.447.461.7554 | Turkey | Asia | Western Asia |
| **SRR6924159** | 27/06/2017 | 1.1.2.1326.447.461.7654 | Turkey | Asia | Western Asia |
| **SRR6920120** | 03/10/2017 | 1.1.2.1326.447.461.8557 | Turkey | Asia | Western Asia |
| **SRR7157822** | 11/09/2015 | 1.1.2.1326.510.531.4291 | Turkey | Asia | Western Asia |
| **SRR1963286** | 22/09/2014 | 1.1.2.1326.510.531.607 | Turkey | Asia | Western Asia |
| **SRR7538877** | 11/06/2015 | 1.1.2.1326.838.1989.2916 | Turkey | Asia | Western Asia |
| **SRR7456864** | 29/06/2015 | 1.1.2.1326.838.1989.3009 | Turkey | Asia | Western Asia |
| **SRR7285751** | 09/07/2015 | 1.1.2.1326.838.1989.3059 | Turkey | Asia | Western Asia |
| **SRR8724939** | 08/07/2015 | 1.1.2.1326.838.1989.3083 | Turkey | Asia | Western Asia |

| | | | | | |
|---|---|---|---|---|---|
| **SRR1966646** | 14/07/2014 | 1.1.2.1326.838.930.1194 | Turkey | Asia | Western Asia |
| **SRR1968252** | 13/05/2014 | 1.1.2.1326.838.930.1766 | Turkey | Asia | Western Asia |
| **SRR7090694** | 15/06/2015 | 1.1.2.1326.838.930.2936 | Turkey | Asia | Western Asia |
| **SRR5220197** | 11/08/2015 | 1.1.2.1326.838.930.3265 | Turkey | Asia | Western Asia |
| **SRR8711760** | 28/08/2015 | 1.1.2.1326.838.930.3406 | Turkey | Asia | Western Asia |
| **SRR8711835** | 09/09/2015 | 1.1.2.1326.838.930.3435 | Turkey | Asia | Western Asia |
| **SRR5220218** | 01/09/2015 | 1.1.2.1326.838.930.3500 | Turkey | Asia | Western Asia |
| **SRR7310568** | 16/12/2015 | 1.1.2.1326.838.930.4215 | Turkey | Asia | Western Asia |
| **SRR5193256** | 16/08/2016 | 1.1.2.1326.838.930.5152 | Turkey | Asia | Western Asia |
| **SRR5193828** | 01/09/2016 | 1.1.2.1326.838.930.5462 | Turkey | Asia | Western Asia |
| **SRR7285796** | 16/06/2015 | 1.1.2.1326.928.1039.2944 | Turkey | Asia | Western Asia |
| **SRR7286850** | 18/06/2015 | 1.1.2.1326.928.1039.2980 | Turkey | Asia | Western Asia |
| **SRR3286933** | 29/06/2015 | 1.1.2.1326.928.1039.3018 | Turkey | Asia | Western Asia |
| **SRR1958134** | 14/10/2014 | 1.1.2.139.139.139.1888 | Turkey | Asia | Western Asia |
| **SRR1958254** | 23/09/2014 | 1.1.2.158.158.158.158 | Turkey | Asia | Western Asia |
| **SRR5216054** | 01/09/2016 | 1.1.2.1659.2406.2999.5439 | Turkey | Asia | Western Asia |
| **SRR5216561** | 14/06/2016 | 1.1.2.1711.1347.2737.4842 | Turkey | Asia | Western Asia |
| **SRR6900161** | 28/06/2016 | 1.1.2.1711.1347.2737.4864 | Turkey | Asia | Western Asia |
| **SRR6900957** | 22/07/2016 | 1.1.2.1711.1347.2737.5029 | Turkey | Asia | Western Asia |
| **SRR8492273** | 01/06/2018 | 1.1.2.1711.2003.2405.10020 | Turkey | Asia | Western Asia |
| **SRR7523071** | 03/07/2018 | 1.1.2.1711.2003.2405.10332 | Turkey | Asia | Western Asia |
| **SRR7842440** | 23/08/2018 | 1.1.2.1711.2003.2405.11428 | Turkey | Asia | Western Asia |
| **SRR7867210** | 22/08/2018 | 1.1.2.1711.2003.2405.11487 | Turkey | Asia | Western Asia |
| **SRR8239942** | 17/09/2018 | 1.1.2.1711.2003.2405.11759 | Turkey | Asia | Western Asia |
| **SRR8492456** | 27/09/2018 | 1.1.2.1711.2003.2405.12041 | Turkey | Asia | Western Asia |
| **SRR8087231** | 05/10/2018 | 1.1.2.1711.2003.2405.12113 | Turkey | Asia | Western Asia |
| **SRR5216167** | 27/06/2016 | 1.1.2.1711.2003.2405.4905 | Turkey | Asia | Western Asia |
| **SRR5220550** | 30/08/2016 | 1.1.2.1711.2003.2405.5349 | Turkey | Asia | Western Asia |
| **SRR5193919** | 21/09/2016 | 1.1.2.1711.2003.2405.5708 | Turkey | Asia | Western Asia |
| **SRR5193616** | 26/09/2016 | 1.1.2.1711.2003.2405.5770 | Turkey | Asia | Western Asia |
| **SRR5216192** | 19/10/2016 | 1.1.2.1711.2003.2405.5933 | Turkey | Asia | Western Asia |
| **SRR6918896** | 17/10/2016 | 1.1.2.1711.2003.2405.5960 | Turkey | Asia | Western Asia |
| **SRR5220464** | 03/11/2016 | 1.1.2.1711.2003.2405.6244 | Turkey | Asia | Western Asia |
| **SRR6900478** | 22/11/2016 | 1.1.2.1711.2003.2405.6333 | Turkey | Asia | Western Asia |
| **SRR6919167** | 07/08/2017 | 1.1.2.1711.2003.2405.7820 | Turkey | Asia | Western Asia |
| **SRR7533351** | 04/07/2018 | 1.1.2.1711.50.50.10326 | Turkey | Asia | Western Asia |
| **SRR1970279** | 03/07/2014 | 1.1.2.1711.50.50.50 | Turkey | Asia | Western Asia |
| **SRR8548724** | 15/08/2017 | 1.1.2.1711.50.50.7964 | Turkey | Asia | Western Asia |
| **SRR6920021** | 23/08/2017 | 1.1.2.1711.50.50.8153 | Turkey | Asia | Western Asia |
| **SRR6922786** | 14/09/2017 | 1.1.2.1711.50.50.8371 | Turkey | Asia | Western Asia |
| **SRR1967455** | 27/08/2014 | 1.1.2.1712.1457.1680.2312 | Turkey | Asia | Western Asia |
| **SRR3284720** | 18/09/2015 | 1.1.2.1712.1870.2222.3518 | Turkey | Asia | Western Asia |
| **SRR7426866** | 29/09/2015 | 1.1.2.1712.1927.2304.3730 | Turkey | Asia | Western Asia |
| **SRR6900951** | 20/09/2016 | 1.1.2.1712.2488.3110.5724 | Turkey | Asia | Western Asia |
| **SRR8300578** | 30/08/2018 | 1.1.2.1712.3863.5393.11559 | Turkey | Asia | Western Asia |
| **SRR5216156** | 21/09/2015 | 1.1.2.1712.475.2032.3577 | Turkey | Asia | Western Asia |
| **SRR8201789** | 02/11/2018 | 1.1.2.1712.475.3156.12369 | Turkey | Asia | Western Asia |
| **SRR8201826** | 06/11/2018 | 1.1.2.1712.475.3156.12373 | Turkey | Asia | Western Asia |

| | | | | | |
|---|---|---|---|---|---|
| **SRR8204039** | 07/11/2018 | 1.1.2.1712.475.3156.12385 | Turkey | Asia | Western Asia |
| **SRR6919852** | 07/10/2016 | 1.1.2.1712.475.3156.5913 | Turkey | Asia | Western Asia |
| **SRR5216372** | 16/11/2015 | 1.1.2.1712.475.492.4062 | Turkey | Asia | Western Asia |
| **SRR5220835** | 23/11/2015 | 1.1.2.1712.475.492.4084 | Turkey | Asia | Western Asia |
| **SRR1967903** | 06/08/2014 | 1.1.2.1712.475.492.759 | Turkey | Asia | Western Asia |
| **SRR1966344** | 18/08/2014 | 1.1.2.1712.475.492.875 | Turkey | Asia | Western Asia |
| **SRR5220872** | 28/08/2015 | 1.1.2.1712.58.2133.3352 | Turkey | Asia | Western Asia |
| **SRR5216280** | 31/05/2016 | 1.1.2.1712.58.2133.4722 | Turkey | Asia | Western Asia |
| **SRR5216521** | 22/06/2016 | 1.1.2.1712.58.2133.4810 | Turkey | Asia | Western Asia |
| **SRR5583161** | 03/05/2017 | 1.1.2.1712.58.2133.7231 | Turkey | Asia | Western Asia |
| **SRR5216346** | 26/08/2015 | 1.1.2.1712.58.2162.3369 | Turkey | Asia | Western Asia |
| **SRR5220534** | 23/09/2015 | 1.1.2.1712.58.2226.3525 | Turkey | Asia | Western Asia |
| **SRR5193534** | 08/11/2016 | 1.1.2.1712.58.3273.6265 | Turkey | Asia | Western Asia |
| **SRR5220489** | 12/05/2015 | 1.1.2.1712.58.370.2811 | Turkey | Asia | Western Asia |
| **SRR7369164** | 29/07/2015 | 1.1.2.1712.58.370.3192 | Turkey | Asia | Western Asia |
| **SRR5216554** | 20/10/2015 | 1.1.2.1712.58.370.3873 | Turkey | Asia | Western Asia |
| **SRR6899267** | 02/08/2016 | 1.1.2.1712.58.370.5078 | Turkey | Asia | Western Asia |
| **SRR5215767** | 07/09/2016 | 1.1.2.1712.58.370.5591 | Turkey | Asia | Western Asia |
| **SRR6920009** | 02/10/2017 | 1.1.2.1712.58.370.8549 | Turkey | Asia | Western Asia |
| **SRR3048784** | 22/08/2014 | 1.1.2.1712.58.566.665 | Turkey | Asia | Western Asia |
| **SRR1970143** | 30/07/2014 | 1.1.2.1712.58.570.1866 | Turkey | Asia | Western Asia |
| **SRR5215754** | 28/09/2015 | 1.1.2.1712.58.570.3683 | Turkey | Asia | Western Asia |
| **SRR3049891** | 04/12/2014 | 1.1.2.1712.58.570.680 | Turkey | Asia | Western Asia |
| **SRR1966074** | 07/10/2014 | 1.1.2.1712.912.1021.1338 | Turkey | Asia | Western Asia |
| **SRR1967522** | 03/10/2014 | 1.1.2.1712.926.1037.1367 | Turkey | Asia | Western Asia |
| **SRR1965488** | 12/08/2014 | 1.1.2.1712.926.1263.1696 | Turkey | Asia | Western Asia |
| **SRR1966568** | 27/06/2014 | 1.1.2.1714.48.48.48 | Turkey | Asia | Western Asia |
| **SRR1969482** | 05/11/2014 | 1.1.2.1714.513.534.615 | Turkey | Asia | Western Asia |
| **SRR1966690** | 19/09/2014 | 1.1.2.176.176.176.176 | Turkey | Asia | Western Asia |
| **SRR8131536** | 14/09/2018 | 1.1.2.1804.3890.5444.11740 | Turkey | Asia | Western Asia |
| **SRR1967009** | 08/08/2014 | 1.1.2.19.19.448.2640 | Turkey | Asia | Western Asia |
| **SRR1966940** | 20/06/2014 | 1.1.2.19.19.448.902 | Turkey | Asia | Western Asia |
| **SRR1967313** | 22/07/2014 | 1.1.2.19.19.515.580 | Turkey | Asia | Western Asia |
| **SRR1958419** | 16/10/2014 | 1.1.2.209.209.209.1237 | Turkey | Asia | Western Asia |
| **SRR1968935** | 21/10/2014 | 1.1.2.209.209.209.209 | Turkey | Asia | Western Asia |
| **SRR6901006** | 05/09/2017 | 1.1.2.2241.3159.4075.8277 | Turkey | Asia | Western Asia |
| **SRR6918309** | 05/09/2017 | 1.1.2.2341.3160.4076.8281 | Turkey | Asia | Western Asia |
| **SRR7458744** | 30/05/2018 | 1.1.2.28.149.149.10058 | Turkey | Asia | Western Asia |
| **SRR7458177** | 19/06/2018 | 1.1.2.28.149.149.10152 | Turkey | Asia | Western Asia |
| **SRR7480299** | 21/06/2018 | 1.1.2.28.149.149.10171 | Turkey | Asia | Western Asia |
| **SRR7480387** | 27/06/2018 | 1.1.2.28.149.149.10180 | Turkey | Asia | Western Asia |
| **SRR7495584** | 25/06/2018 | 1.1.2.28.149.149.10191 | Turkey | Asia | Western Asia |
| **SRR7850622** | 11/07/2018 | 1.1.2.28.149.149.10389 | Turkey | Asia | Western Asia |
| **SRR7828462** | 06/08/2018 | 1.1.2.28.149.149.10419 | Turkey | Asia | Western Asia |
| **SRR7902693** | 13/09/2018 | 1.1.2.28.149.149.11417 | Turkey | Asia | Western Asia |
| **SRR7866896** | 03/09/2018 | 1.1.2.28.149.149.11420 | Turkey | Asia | Western Asia |
| **SRR8399307** | 18/12/2018 | 1.1.2.28.149.149.11650 | Turkey | Asia | Western Asia |
| **SRR7997072** | 19/09/2018 | 1.1.2.28.149.149.11821 | Turkey | Asia | Western Asia |

| | | | | | |
|---|---|---|---|---|---|
| **SRR8087138** | 27/09/2018 | 1.1.2.28.149.149.12019 | Turkey | Asia | Western Asia |
| **SRR8272584** | 27/09/2018 | 1.1.2.28.149.149.12029 | Turkey | Asia | Western Asia |
| **SRR8087242** | 01/10/2018 | 1.1.2.28.149.149.12044 | Turkey | Asia | Western Asia |
| **SRR8084331** | 02/10/2018 | 1.1.2.28.149.149.12070 | Turkey | Asia | Western Asia |
| **SRR8084247** | 02/10/2018 | 1.1.2.28.149.149.12080 | Turkey | Asia | Western Asia |
| **SRR8204756** | 03/10/2018 | 1.1.2.28.149.149.12107 | Turkey | Asia | Western Asia |
| **SRR8084308** | 09/10/2018 | 1.1.2.28.149.149.12147 | Turkey | Asia | Western Asia |
| **SRR8297316** | 28/08/2018 | 1.1.2.28.149.149.12281 | Turkey | Asia | Western Asia |
| **SRR8137418** | 17/10/2018 | 1.1.2.28.149.149.12283 | Turkey | Asia | Western Asia |
| **SRR8172829** | 26/10/2018 | 1.1.2.28.149.149.12305 | Turkey | Asia | Western Asia |
| **SRR8204752** | 29/10/2018 | 1.1.2.28.149.149.12320 | Turkey | Asia | Western Asia |
| **SRR8775490** | 13/03/2019 | 1.1.2.28.149.149.12849 | Turkey | Asia | Western Asia |
| **SRR7350851** | 18/06/2015 | 1.1.2.28.149.149.149 | Turkey | Asia | Western Asia |
| **SRR3285404** | 21/04/2015 | 1.1.2.28.149.149.2745 | Turkey | Asia | Western Asia |
| **SRR3284837** | 28/04/2015 | 1.1.2.28.149.149.2749 | Turkey | Asia | Western Asia |
| **SRR5193402** | 08/06/2015 | 1.1.2.28.149.149.2898 | Turkey | Asia | Western Asia |
| **SRR5193417** | 08/06/2015 | 1.1.2.28.149.149.2904 | Turkey | Asia | Western Asia |
| **SRR5220707** | 11/06/2015 | 1.1.2.28.149.149.2908 | Turkey | Asia | Western Asia |
| **SRR5215823** | 11/06/2015 | 1.1.2.28.149.149.2929 | Turkey | Asia | Western Asia |
| **SRR5220886** | 01/07/2015 | 1.1.2.28.149.149.3002 | Turkey | Asia | Western Asia |
| **SRR7444046** | 30/06/2015 | 1.1.2.28.149.149.3038 | Turkey | Asia | Western Asia |
| **SRR5220196** | 16/07/2015 | 1.1.2.28.149.149.3145 | Turkey | Asia | Western Asia |
| **SRR5194201** | 21/07/2015 | 1.1.2.28.149.149.3161 | Turkey | Asia | Western Asia |
| **SRR7284472** | 21/07/2015 | 1.1.2.28.149.149.3162 | Turkey | Asia | Western Asia |
| **SRR7223181** | 04/08/2016 | 1.1.2.28.149.149.3255 | Turkey | Asia | Western Asia |
| **SRR7962271** | 25/08/2015 | 1.1.2.28.149.149.3347 | Turkey | Asia | Western Asia |
| **SRR5193838** | 20/10/2015 | 1.1.2.28.149.149.3870 | Turkey | Asia | Western Asia |
| **SRR5194049** | 03/11/2015 | 1.1.2.28.149.149.4018 | Turkey | Asia | Western Asia |
| **SRR6920102** | 24/06/2016 | 1.1.2.28.149.149.4877 | Turkey | Asia | Western Asia |
| **SRR6898812** | 04/07/2017 | 1.1.2.28.149.149.7688 | Turkey | Asia | Western Asia |
| **SRR6898409** | 12/07/2017 | 1.1.2.28.149.149.7781 | Turkey | Asia | Western Asia |
| **SRR6897828** | 27/07/2017 | 1.1.2.28.149.149.7811 | Turkey | Asia | Western Asia |
| **SRR6897666** | 19/07/2017 | 1.1.2.28.149.149.7826 | Turkey | Asia | Western Asia |
| **SRR7480133** | 15/08/2017 | 1.1.2.28.149.149.7874 | Turkey | Asia | Western Asia |
| **SRR6919805** | 04/08/2017 | 1.1.2.28.149.149.7916 | Turkey | Asia | Western Asia |
| **SRR6900278** | 31/07/2017 | 1.1.2.28.149.149.8065 | Turkey | Asia | Western Asia |
| **SRR6898827** | 23/08/2017 | 1.1.2.28.149.149.8158 | Turkey | Asia | Western Asia |
| **SRR6921908** | 22/08/2017 | 1.1.2.28.149.149.8187 | Turkey | Asia | Western Asia |
| **SRR6919215** | 16/08/2017 | 1.1.2.28.149.149.8193 | Turkey | Asia | Western Asia |
| **SRR6898473** | 18/09/2017 | 1.1.2.28.149.149.8372 | Turkey | Asia | Western Asia |
| **SRR6919976** | 15/09/2017 | 1.1.2.28.149.149.8381 | Turkey | Asia | Western Asia |
| **SRR6918858** | 04/10/2017 | 1.1.2.28.149.149.8594 | Turkey | Asia | Western Asia |
| **SRR6899271** | 18/10/2017 | 1.1.2.28.149.149.8687 | Turkey | Asia | Western Asia |
| **SRR6921975** | 23/10/2017 | 1.1.2.28.149.149.8726 | Turkey | Asia | Western Asia |
| **SRR6924136** | 29/11/2017 | 1.1.2.28.149.149.8940 | Turkey | Asia | Western Asia |
| **SRR6898833** | 30/08/2017 | 1.1.2.28.149.149.9597 | Turkey | Asia | Western Asia |
| **SRR5220540** | 09/06/2015 | 1.1.2.28.149.1986.2911 | Turkey | Asia | Western Asia |
| **SRR8724640** | 17/06/2015 | 1.1.2.28.149.2008.2957 | Turkey | Asia | Western Asia |

| | | | | | |
|---|---|---|---|---|---|
| **SRR1968048** | 29/07/2014 | 1.1.2.28.149.926.1189 | Turkey | Asia | Western Asia |
| **SRR5216348** | 27/07/2015 | 1.1.2.28.1761.2070.3164 | Turkey | Asia | Western Asia |
| **SRR7867161** | 31/07/2018 | 1.1.2.28.3641.4889.10550 | Turkey | Asia | Western Asia |
| **SRR3049060** | 17/06/2014 | 1.1.2.28.387.390.401 | Turkey | Asia | Western Asia |
| **SRR6920101** | 06/12/2016 | 1.1.2.29.2627.3319.6419 | Turkey | Asia | Western Asia |
| **SRR8717171** | 19/08/2015 | 1.1.2.29.29.2132.3309 | Turkey | Asia | Western Asia |
| **SRR1965336** | 20/05/2014 | 1.1.2.29.29.29.29 | Turkey | Asia | Western Asia |
| **SRR3049665** | 10/06/2014 | 1.1.2.29.29.29.491 | Turkey | Asia | Western Asia |
| **SRR7828426** | 09/08/2018 | 1.1.2.29.29.5338.11397 | Turkey | Asia | Western Asia |
| **SRR7850538** | 13/08/2018 | 1.1.2.29.29.5338.11413 | Turkey | Asia | Western Asia |
| **SRR7903094** | 22/08/2018 | 1.1.2.29.29.5338.11478 | Turkey | Asia | Western Asia |
| **SRR5194023** | 04/11/2016 | 1.1.2.29.937.1049.6228 | Turkey | Asia | Western Asia |
| **SRR3048858** | 04/06/2014 | 1.1.2.48.598.638.771 | Turkey | Asia | Western Asia |
| **SRR7191862** | 03/11/2015 | 1.1.2.63.63.63.3992 | Turkey | Asia | Western Asia |
| **SRR3049473** | 21/07/2014 | 1.1.2.65.65.65.65 | Turkey | Asia | Western Asia |
| **SRR5193905** | 30/08/2016 | 1.1.2.80.1946.2988.5425 | Turkey | Asia | Western Asia |
| **SRR6900846** | 31/07/2017 | 1.1.2.80.453.467.7980 | Turkey | Asia | Western Asia |
| **SRR7439485** | 22/09/2015 | 1.1.540.2067.2883.3657.3692 | Turkey | Asia | Western Asia |
| **SRR7523769** | 13/10/2015 | 1.1.540.2067.2883.3657.3704 | Turkey | Asia | Western Asia |
| **SRR6899436** | 06/09/2016 | 1.1.540.2067.2883.3657.5528 | Turkey | Asia | Western Asia |
| **SRR6924120** | 26/10/2016 | 1.1.540.2067.2883.3657.6093 | Turkey | Asia | Western Asia |
| **SRR6919043** | 05/06/2017 | 1.1.540.2067.2883.3657.7426 | Turkey | Asia | Western Asia |
| **SRR6900148** | 13/06/2017 | 1.1.540.2067.2883.3657.7544 | Turkey | Asia | Western Asia |
| **SRR8304794** | 21/08/2018 | 1.1.540.2067.2883.5353.11434 | Turkey | Asia | Western Asia |
| **SRR8272633** | 07/09/2018 | 1.1.540.2067.2883.5353.11656 | Turkey | Asia | Western Asia |
| **SRR1968089** | 09/09/2014 | 1.11.82.135.135.135.135 | Turkey | Asia | Western Asia |
| **SRR7516257** | 28/06/2018 | 1.2.270.2317.3556.4720.10284 | Turkey | Asia | Western Asia |
| **SRR8249754** | 15/11/2018 | 1.2.270.2317.3556.4720.12446 | Turkey | Asia | Western Asia |
| **SRR7841372** | 10/08/2018 | 1.2.270.2317.3556.5334.11389 | Turkey | Asia | Western Asia |
| **SRR7850530** | 14/08/2018 | 1.2.270.2317.3556.5334.11403 | Turkey | Asia | Western Asia |
| **SRR8333797** | 15/08/2018 | 1.2.270.2317.3556.5334.11474 | Turkey | Asia | Western Asia |
| **SRR7903058** | 11/09/2018 | 1.2.270.2317.3556.5334.11525 | Turkey | Asia | Western Asia |
| **SRR8293775** | 04/09/2018 | 1.2.270.2317.3556.5334.11592 | Turkey | Asia | Western Asia |
| **SRR7962279** | 19/09/2018 | 1.2.270.2317.3556.5334.11719 | Turkey | Asia | Western Asia |
| **SRR8204780** | 16/10/2018 | 1.2.270.2317.3556.5334.12191 | Turkey | Asia | Western Asia |
| **SRR7523798** | 17/09/2015 | 1.2.3.151.151.783.2809 | Turkey | Asia | Western Asia |
| **SRR7523124** | 02/07/2018 | 1.2.3.151.3522.4653.10135 | Turkey | Asia | Western Asia |
| **SRR5216536** | 17/11/2016 | 1.2.3.151.356.372.6293 | Turkey | Asia | Western Asia |
| **SRR1962576** | 18/11/2014 | 1.2.3.18.1338.1534.2089 | Turkey | Asia | Western Asia |
| **SRR4063719** | 14/07/2015 | 1.2.3.18.1755.2061.3135 | Turkey | Asia | Western Asia |
| **SRR6900710** | 22/08/2017 | 1.2.3.18.1755.4051.8179 | Turkey | Asia | Western Asia |
| **SRR7527826** | 08/09/2015 | 1.2.3.18.180.180.10869 | Turkey | Asia | Western Asia |
| **SRR6899339** | 17/08/2017 | 1.2.3.18.180.180.10885 | Turkey | Asia | Western Asia |
| **SRR6899308** | 29/06/2016 | 1.2.3.18.180.180.10897 | Turkey | Asia | Western Asia |
| **SRR7284432** | 18/09/2015 | 1.2.3.18.180.180.11054 | Turkey | Asia | Western Asia |
| **SRR5220854** | 07/10/2015 | 1.2.3.18.180.180.11139 | Turkey | Asia | Western Asia |
| **SRR8705985** | 15/10/2015 | 1.2.3.18.180.180.11142 | Turkey | Asia | Western Asia |
| **SRR6920736** | 12/09/2017 | 1.2.3.18.180.180.11159 | Turkey | Asia | Western Asia |

| | | | | | |
|---|---|---|---|---|---|
| **SRR5193539** | 12/07/2016 | 1.2.3.18.180.180.11226 | Turkey | Asia | Western Asia |
| **SRR6919165** | 06/12/2017 | 1.2.3.18.180.180.11280 | Turkey | Asia | Western Asia |
| **SRR6922071** | 26/08/2016 | 1.2.3.18.180.180.11286 | Turkey | Asia | Western Asia |
| **SRR6919791** | 23/08/2017 | 1.2.3.18.180.180.11287 | Turkey | Asia | Western Asia |
| **SRR8490782** | 12/09/2018 | 1.2.3.18.180.180.11706 | Turkey | Asia | Western Asia |
| **SRR8182953** | 24/10/2018 | 1.2.3.18.180.180.12273 | Turkey | Asia | Western Asia |
| **SRR1969107** | 13/11/2014 | 1.2.3.18.180.5110.10833 | Turkey | Asia | Western Asia |
| **SRR7842629** | 30/08/2018 | 1.2.3.18.180.5398.11564 | Turkey | Asia | Western Asia |
| **SRR7962299** | 17/09/2018 | 1.2.3.18.180.5398.11754 | Turkey | Asia | Western Asia |
| **SRR5220754** | 02/10/2015 | 1.2.3.18.1936.2319.3762 | Turkey | Asia | Western Asia |
| **SRR6898073** | 11/07/2017 | 1.2.3.18.1979.2369.7782 | Turkey | Asia | Western Asia |
| **SRR5215758** | 04/05/2016 | 1.2.3.18.2180.2669.4650 | Turkey | Asia | Western Asia |
| **SRR7873972** | 20/07/2018 | 1.2.3.18.2180.4874.10505 | Turkey | Asia | Western Asia |
| **SRR6900879** | 22/06/2016 | 1.2.3.18.2223.2744.4861 | Turkey | Asia | Western Asia |
| **SRR5220672** | 31/08/2016 | 1.2.3.18.2440.3042.5515 | Turkey | Asia | Western Asia |
| **SRR5194219** | 06/09/2016 | 1.2.3.18.2445.3047.5539 | Turkey | Asia | Western Asia |
| **SRR6919263** | 08/09/2016 | 1.2.3.18.2445.3047.5562 | Turkey | Asia | Western Asia |
| **SRR5194312** | 12/09/2016 | 1.2.3.18.2445.3047.5633 | Turkey | Asia | Western Asia |
| **SRR6900927** | 06/10/2016 | 1.2.3.18.2445.3047.5897 | Turkey | Asia | Western Asia |
| **SRR5215630** | 11/10/2016 | 1.2.3.18.2445.3047.5925 | Turkey | Asia | Western Asia |
| **SRR5193723** | 11/10/2016 | 1.2.3.18.2445.3047.5928 | Turkey | Asia | Western Asia |
| **SRR5194333** | 07/09/2016 | 1.2.3.18.2445.3051.5547 | Turkey | Asia | Western Asia |
| **SRR7475284** | 30/05/2018 | 1.2.3.18.3094.3949.10063 | Turkey | Asia | Western Asia |
| **SRR6897021** | 03/08/2017 | 1.2.3.18.3094.3949.7793 | Turkey | Asia | Western Asia |
| **SRR6922626** | 19/07/2017 | 1.2.3.18.3094.3949.8022 | Turkey | Asia | Western Asia |
| **SRR7349209** | 29/05/2018 | 1.2.3.18.3097.3953.10041 | Turkey | Asia | Western Asia |
| **SRR8492395** | 05/06/2018 | 1.2.3.18.3097.3953.10087 | Turkey | Asia | Western Asia |
| **SRR8445028** | 05/07/2018 | 1.2.3.18.3097.3953.10335 | Turkey | Asia | Western Asia |
| **SRR7879561** | 11/07/2018 | 1.2.3.18.3097.3953.10387 | Turkey | Asia | Western Asia |
| **SRR7866994** | 17/07/2018 | 1.2.3.18.3097.3953.10440 | Turkey | Asia | Western Asia |
| **SRR8131581** | 18/09/2018 | 1.2.3.18.3097.3953.10470 | Turkey | Asia | Western Asia |
| **SRR7903068** | 12/09/2018 | 1.2.3.18.3097.3953.11697 | Turkey | Asia | Western Asia |
| **SRR8087245** | 02/10/2018 | 1.2.3.18.3097.3953.12061 | Turkey | Asia | Western Asia |
| **SRR6899421** | 18/08/2017 | 1.2.3.18.3097.3953.8151 | Turkey | Asia | Western Asia |
| **SRR6900921** | 17/10/2017 | 1.2.3.18.3097.3953.8674 | Turkey | Asia | Western Asia |
| **SRR6897947** | 08/08/2017 | 1.2.3.18.3099.3963.7845 | Turkey | Asia | Western Asia |
| **SRR6920159** | 31/08/2017 | 1.2.3.18.3155.4068.8254 | Turkey | Asia | Western Asia |
| **SRR6897664** | 12/10/2017 | 1.2.3.18.3223.4188.8644 | Turkey | Asia | Western Asia |
| **SRR6921931** | 06/11/2017 | 1.2.3.18.3247.4236.8791 | Turkey | Asia | Western Asia |
| **SRR1957763** | 10/09/2014 | 1.2.3.18.353.1017.1320 | Turkey | Asia | Western Asia |
| **SRR1969011** | 29/05/2014 | 1.2.3.18.353.1112.1483 | Turkey | Asia | Western Asia |
| **SRR1957762** | 10/09/2014 | 1.2.3.18.353.1123.1496 | Turkey | Asia | Western Asia |
| **SRR3048861** | 04/06/2014 | 1.2.3.18.353.1172.1564 | Turkey | Asia | Western Asia |
| **SRR3048836** | 02/10/2014 | 1.2.3.18.353.1246.1675 | Turkey | Asia | Western Asia |
| **SRR1965514** | 01/07/2014 | 1.2.3.18.353.1340.1808 | Turkey | Asia | Western Asia |
| **SRR8724711** | 29/07/2015 | 1.2.3.18.353.1340.3199 | Turkey | Asia | Western Asia |
| **SRR1966309** | 11/09/2014 | 1.2.3.18.353.1468.2068 | Turkey | Asia | Western Asia |
| **SRR7468867** | 04/08/2015 | 1.2.3.18.353.1468.3163 | Turkey | Asia | Western Asia |

| | | | | | |
|---|---|---|---|---|---|
| **SRR5215620** | 07/07/2015 | 1.2.3.18.353.1744.2976 | Turkey | Asia | Western Asia |
| **SRR7292844** | 21/07/2015 | 1.2.3.18.353.1744.3160 | Turkey | Asia | Western Asia |
| **SRR5194197** | 27/07/2015 | 1.2.3.18.353.1744.3177 | Turkey | Asia | Western Asia |
| **SRR7358937** | 11/08/2015 | 1.2.3.18.353.1744.3291 | Turkey | Asia | Western Asia |
| **SRR5220437** | 14/08/2015 | 1.2.3.18.353.1744.3321 | Turkey | Asia | Western Asia |
| **SRR7480281** | 03/11/2015 | 1.2.3.18.353.1744.3994 | Turkey | Asia | Western Asia |
| **SRR7439501** | 08/09/2015 | 1.2.3.18.353.1744.4281 | Turkey | Asia | Western Asia |
| **SRR6900990** | 21/06/2016 | 1.2.3.18.353.1744.4858 | Turkey | Asia | Western Asia |
| **SRR1959484** | 22/09/2014 | 1.2.3.18.353.1843.2575 | Turkey | Asia | Western Asia |
| **SRR5220941** | 18/06/2015 | 1.2.3.18.353.2009.2958 | Turkey | Asia | Western Asia |
| **SRR8724785** | 26/06/2015 | 1.2.3.18.353.2030.3021 | Turkey | Asia | Western Asia |
| **SRR5220603** | 30/09/2015 | 1.2.3.18.353.2030.3691 | Turkey | Asia | Western Asia |
| **SRR8705989** | 06/10/2015 | 1.2.3.18.353.2030.3742 | Turkey | Asia | Western Asia |
| **SRR6921948** | 19/10/2016 | 1.2.3.18.353.2030.6049 | Turkey | Asia | Western Asia |
| **SRR5220662** | 28/07/2015 | 1.2.3.18.353.2081.3195 | Turkey | Asia | Western Asia |
| **SRR7527780** | 20/08/2015 | 1.2.3.18.353.2081.3310 | Turkey | Asia | Western Asia |
| **SRR5220759** | 20/08/2015 | 1.2.3.18.353.2081.3311 | Turkey | Asia | Western Asia |
| **SRR8711380** | 08/09/2015 | 1.2.3.18.353.2081.3472 | Turkey | Asia | Western Asia |
| **SRR5193522** | 09/09/2015 | 1.2.3.18.353.2081.3508 | Turkey | Asia | Western Asia |
| **SRR8711623** | 04/09/2015 | 1.2.3.18.353.2081.3546 | Turkey | Asia | Western Asia |
| **SRR7443924** | 15/09/2015 | 1.2.3.18.353.2081.3635 | Turkey | Asia | Western Asia |
| **SRR7867057** | 09/08/2018 | 1.2.3.18.353.2111.11378 | Turkey | Asia | Western Asia |
| **SRR7828371** | 29/08/2018 | 1.2.3.18.353.2111.11492 | Turkey | Asia | Western Asia |
| **SRR7850459** | 31/08/2018 | 1.2.3.18.353.2111.11534 | Turkey | Asia | Western Asia |
| **SRR8114886** | 28/09/2018 | 1.2.3.18.353.2111.12046 | Turkey | Asia | Western Asia |
| **SRR8166236** | 16/10/2018 | 1.2.3.18.353.2111.12213 | Turkey | Asia | Western Asia |
| **SRR8142758** | 18/10/2018 | 1.2.3.18.353.2111.12214 | Turkey | Asia | Western Asia |
| **SRR8137419** | 17/10/2018 | 1.2.3.18.353.2111.12225 | Turkey | Asia | Western Asia |
| **SRR8149430** | 24/10/2018 | 1.2.3.18.353.2111.12271 | Turkey | Asia | Western Asia |
| **SRR5220424** | 26/10/2016 | 1.2.3.18.353.2111.6103 | Turkey | Asia | Western Asia |
| **SRR7157809** | 12/08/2015 | 1.2.3.18.353.2115.3282 | Turkey | Asia | Western Asia |
| **SRR5193871** | 13/09/2016 | 1.2.3.18.353.2128.5668 | Turkey | Asia | Western Asia |
| **SRR5216286** | 20/09/2016 | 1.2.3.18.353.2128.5980 | Turkey | Asia | Western Asia |
| **SRR4063718** | 14/09/2015 | 1.2.3.18.353.2236.3552 | Turkey | Asia | Western Asia |
| **SRR4063754** | 06/10/2015 | 1.2.3.18.353.2293.3706 | Turkey | Asia | Western Asia |
| **SRR5220851** | 09/11/2015 | 1.2.3.18.353.2413.3977 | Turkey | Asia | Western Asia |
| **SRR4063734** | 09/11/2015 | 1.2.3.18.353.2415.3982 | Turkey | Asia | Western Asia |
| **SRR7828377** | 31/08/2018 | 1.2.3.18.353.2461.10481 | Turkey | Asia | Western Asia |
| **SRR8149126** | 22/10/2018 | 1.2.3.18.353.2461.12264 | Turkey | Asia | Western Asia |
| **SRR8172823** | 26/10/2018 | 1.2.3.18.353.2461.12304 | Turkey | Asia | Western Asia |
| **SRR5220433** | 25/08/2015 | 1.2.3.18.353.2861.3356 | Turkey | Asia | Western Asia |
| **SRR5220503** | 03/09/2015 | 1.2.3.18.353.2861.3411 | Turkey | Asia | Western Asia |
| **SRR5216231** | 25/08/2016 | 1.2.3.18.353.2864.5204 | Turkey | Asia | Western Asia |
| **SRR7850659** | 15/08/2018 | 1.2.3.18.353.353.10019 | Turkey | Asia | Western Asia |
| **SRR8499126** | 04/06/2018 | 1.2.3.18.353.353.10051 | Turkey | Asia | Western Asia |
| **SRR8499354** | 04/06/2018 | 1.2.3.18.353.353.10065 | Turkey | Asia | Western Asia |
| **SRR1967231** | 16/09/2014 | 1.2.3.18.353.353.1008 | Turkey | Asia | Western Asia |
| **SRR8492414** | 11/06/2018 | 1.2.3.18.353.353.10112 | Turkey | Asia | Western Asia |

| | | | | | |
|---|---|---|---|---|---|
| **SRR7439421** | 30/05/2018 | 1.2.3.18.353.353.10122 | Turkey | Asia | Western Asia |
| **SRR7533243** | 05/07/2018 | 1.2.3.18.353.353.10339 | Turkey | Asia | Western Asia |
| **SRR7828464** | 16/08/2018 | 1.2.3.18.353.353.11424 | Turkey | Asia | Western Asia |
| **SRR8307305** | 17/08/2018 | 1.2.3.18.353.353.11437 | Turkey | Asia | Western Asia |
| **SRR7842507** | 20/08/2018 | 1.2.3.18.353.353.11467 | Turkey | Asia | Western Asia |
| **SRR7873868** | 22/08/2018 | 1.2.3.18.353.353.11483 | Turkey | Asia | Western Asia |
| **SRR7885124** | 22/08/2018 | 1.2.3.18.353.353.11493 | Turkey | Asia | Western Asia |
| **SRR8293822** | 05/09/2018 | 1.2.3.18.353.353.11635 | Turkey | Asia | Western Asia |
| **SRR7911467** | 13/09/2018 | 1.2.3.18.353.353.11723 | Turkey | Asia | Western Asia |
| **SRR7962323** | 18/09/2018 | 1.2.3.18.353.353.11791 | Turkey | Asia | Western Asia |
| **SRR8204032** | 07/11/2018 | 1.2.3.18.353.353.11799 | Turkey | Asia | Western Asia |
| **SRR8204748** | 19/09/2018 | 1.2.3.18.353.353.11800 | Turkey | Asia | Western Asia |
| **SRR7962000** | 20/09/2018 | 1.2.3.18.353.353.11803 | Turkey | Asia | Western Asia |
| **SRR7962025** | 19/09/2018 | 1.2.3.18.353.353.11816 | Turkey | Asia | Western Asia |
| **SRR8490764** | 01/10/2018 | 1.2.3.18.353.353.12035 | Turkey | Asia | Western Asia |
| **SRR8106777** | 02/10/2018 | 1.2.3.18.353.353.12068 | Turkey | Asia | Western Asia |
| **SRR8084291** | 08/10/2018 | 1.2.3.18.353.353.12092 | Turkey | Asia | Western Asia |
| **SRR8485116** | 08/10/2018 | 1.2.3.18.353.353.12097 | Turkey | Asia | Western Asia |
| **SRR8114910** | 05/10/2018 | 1.2.3.18.353.353.12106 | Turkey | Asia | Western Asia |
| **SRR8098430** | 10/10/2018 | 1.2.3.18.353.353.12122 | Turkey | Asia | Western Asia |
| **SRR8087198** | 10/10/2018 | 1.2.3.18.353.353.12129 | Turkey | Asia | Western Asia |
| **SRR8098327** | 09/10/2018 | 1.2.3.18.353.353.12141 | Turkey | Asia | Western Asia |
| **SRR8204801** | 01/11/2018 | 1.2.3.18.353.353.12160 | Turkey | Asia | Western Asia |
| **SRR8117006** | 16/10/2018 | 1.2.3.18.353.353.12190 | Turkey | Asia | Western Asia |
| **SRR8204788** | 24/10/2018 | 1.2.3.18.353.353.12274 | Turkey | Asia | Western Asia |
| **SRR8293817** | 07/11/2018 | 1.2.3.18.353.353.12390 | Turkey | Asia | Western Asia |
| **SRR8249902** | 13/11/2018 | 1.2.3.18.353.353.12438 | Turkey | Asia | Western Asia |
| **SRR8438038** | 31/12/2018 | 1.2.3.18.353.353.12601 | Turkey | Asia | Western Asia |
| **SRR8548619** | 23/01/2019 | 1.2.3.18.353.353.12681 | Turkey | Asia | Western Asia |
| **SRR9287708** | 17/04/2019 | 1.2.3.18.353.353.12981 | Turkey | Asia | Western Asia |
| **SRR1958479** | 13/10/2014 | 1.2.3.18.353.353.1374 | Turkey | Asia | Western Asia |
| **SRR1967839** | 05/11/2014 | 1.2.3.18.353.353.1463 | Turkey | Asia | Western Asia |
| **SRR3049019** | 13/06/2014 | 1.2.3.18.353.353.1669 | Turkey | Asia | Western Asia |
| **SRR1957921** | 15/09/2014 | 1.2.3.18.353.353.1965 | Turkey | Asia | Western Asia |
| **SRR1969715** | 04/08/2014 | 1.2.3.18.353.353.2022 | Turkey | Asia | Western Asia |
| **SRR1967698** | 18/09/2014 | 1.2.3.18.353.353.2057 | Turkey | Asia | Western Asia |
| **SRR1966741** | 29/07/2014 | 1.2.3.18.353.353.2322 | Turkey | Asia | Western Asia |
| **SRR1965594** | 23/07/2014 | 1.2.3.18.353.353.2330 | Turkey | Asia | Western Asia |
| **SRR1969337** | 17/09/2014 | 1.2.3.18.353.353.2545 | Turkey | Asia | Western Asia |
| **SRR3048806** | 11/06/2014 | 1.2.3.18.353.353.2662 | Turkey | Asia | Western Asia |
| **SRR1958580** | 15/09/2014 | 1.2.3.18.353.353.2666 | Turkey | Asia | Western Asia |
| **SRR3286717** | 18/05/2015 | 1.2.3.18.353.353.2791 | Turkey | Asia | Western Asia |
| **SRR3286947** | 01/07/2015 | 1.2.3.18.353.353.3016 | Turkey | Asia | Western Asia |
| **SRR5220895** | 10/07/2015 | 1.2.3.18.353.353.3080 | Turkey | Asia | Western Asia |
| **SRR5215707** | 10/07/2015 | 1.2.3.18.353.353.3085 | Turkey | Asia | Western Asia |
| **SRR3285457** | 17/07/2015 | 1.2.3.18.353.353.3105 | Turkey | Asia | Western Asia |
| **SRR5193793** | 24/09/2015 | 1.2.3.18.353.353.3266 | Turkey | Asia | Western Asia |
| **SRR5220837** | 10/08/2015 | 1.2.3.18.353.353.3285 | Turkey | Asia | Western Asia |

| | | | | | |
|---|---|---|---|---|---|
| **SRR7480452** | 14/08/2015 | 1.2.3.18.353.353.3304 | Turkey | Asia | Western Asia |
| **SRR5193423** | 14/09/2015 | 1.2.3.18.353.353.3407 | Turkey | Asia | Western Asia |
| **SRR5220210** | 03/09/2015 | 1.2.3.18.353.353.3408 | Turkey | Asia | Western Asia |
| **SRR7890211** | 21/09/2015 | 1.2.3.18.353.353.3465 | Turkey | Asia | Western Asia |
| **SRR5216058** | 17/09/2015 | 1.2.3.18.353.353.3522 | Turkey | Asia | Western Asia |
| **SRR1970036** | 29/07/2014 | 1.2.3.18.353.353.353 | Turkey | Asia | Western Asia |
| **SRR7292684** | 02/10/2015 | 1.2.3.18.353.353.3535 | Turkey | Asia | Western Asia |
| **SRR8711357** | 04/09/2015 | 1.2.3.18.353.353.3561 | Turkey | Asia | Western Asia |
| **SRR5220526** | 07/09/2015 | 1.2.3.18.353.353.3562 | Turkey | Asia | Western Asia |
| **SRR5193349** | 09/09/2015 | 1.2.3.18.353.353.3583 | Turkey | Asia | Western Asia |
| **SRR7416205** | 07/09/2015 | 1.2.3.18.353.353.3613 | Turkey | Asia | Western Asia |
| **SRR7443876** | 24/09/2015 | 1.2.3.18.353.353.3633 | Turkey | Asia | Western Asia |
| **SRR7456820** | 29/09/2015 | 1.2.3.18.353.353.3673 | Turkey | Asia | Western Asia |
| **SRR5193693** | 10/11/2015 | 1.2.3.18.353.353.3709 | Turkey | Asia | Western Asia |
| **SRR8706139** | 07/10/2015 | 1.2.3.18.353.353.3774 | Turkey | Asia | Western Asia |
| **SRR7458016** | 27/10/2015 | 1.2.3.18.353.353.3932 | Turkey | Asia | Western Asia |
| **SRR3284755** | 03/11/2015 | 1.2.3.18.353.353.4008 | Turkey | Asia | Western Asia |
| **SRR5220150** | 16/11/2015 | 1.2.3.18.353.353.4063 | Turkey | Asia | Western Asia |
| **SRR8172833** | 23/10/2018 | 1.2.3.18.353.353.4079 | Turkey | Asia | Western Asia |
| **SRR7890455** | 20/11/2015 | 1.2.3.18.353.353.4143 | Turkey | Asia | Western Asia |
| **SRR5215747** | 10/06/2016 | 1.2.3.18.353.353.4257 | Turkey | Asia | Western Asia |
| **SRR1958072** | 23/09/2014 | 1.2.3.18.353.353.465 | Turkey | Asia | Western Asia |
| **SRR5220657** | 18/05/2016 | 1.2.3.18.353.353.4688 | Turkey | Asia | Western Asia |
| **SRR5220606** | 01/06/2016 | 1.2.3.18.353.353.4753 | Turkey | Asia | Western Asia |
| **SRR7351552** | 07/06/2016 | 1.2.3.18.353.353.4790 | Turkey | Asia | Western Asia |
| **SRR6900209** | 12/08/2016 | 1.2.3.18.353.353.4794 | Turkey | Asia | Western Asia |
| **SRR1968327** | 26/08/2014 | 1.2.3.18.353.353.481 | Turkey | Asia | Western Asia |
| **SRR6918584** | 21/06/2016 | 1.2.3.18.353.353.4869 | Turkey | Asia | Western Asia |
| **SRR6900642** | 04/07/2016 | 1.2.3.18.353.353.4888 | Turkey | Asia | Western Asia |
| **SRR1967592** | 17/09/2014 | 1.2.3.18.353.353.489 | Turkey | Asia | Western Asia |
| **SRR5215675** | 05/07/2016 | 1.2.3.18.353.353.4936 | Turkey | Asia | Western Asia |
| **SRR6919268** | 19/07/2016 | 1.2.3.18.353.353.5008 | Turkey | Asia | Western Asia |
| **SRR7516667** | 05/08/2016 | 1.2.3.18.353.353.5034 | Turkey | Asia | Western Asia |
| **SRR6919759** | 27/07/2016 | 1.2.3.18.353.353.5051 | Turkey | Asia | Western Asia |
| **SRR6898394** | 04/08/2016 | 1.2.3.18.353.353.5079 | Turkey | Asia | Western Asia |
| **SRR5220923** | 19/08/2016 | 1.2.3.18.353.353.5190 | Turkey | Asia | Western Asia |
| **SRR5220435** | 24/08/2016 | 1.2.3.18.353.353.5220 | Turkey | Asia | Western Asia |
| **SRR6918297** | 25/08/2016 | 1.2.3.18.353.353.5221 | Turkey | Asia | Western Asia |
| **SRR5215753** | 23/08/2016 | 1.2.3.18.353.353.5233 | Turkey | Asia | Western Asia |
| **SRR5220658** | 23/08/2016 | 1.2.3.18.353.353.5253 | Turkey | Asia | Western Asia |
| **SRR6924118** | 02/09/2016 | 1.2.3.18.353.353.5338 | Turkey | Asia | Western Asia |
| **SRR5193476** | 01/09/2016 | 1.2.3.18.353.353.5413 | Turkey | Asia | Western Asia |
| **SRR1965726** | 22/10/2014 | 1.2.3.18.353.353.546 | Turkey | Asia | Western Asia |
| **SRR6918608** | 01/09/2016 | 1.2.3.18.353.353.5486 | Turkey | Asia | Western Asia |
| **SRR5193093** | 01/09/2016 | 1.2.3.18.353.353.5493 | Turkey | Asia | Western Asia |
| **SRR5194059** | 08/11/2016 | 1.2.3.18.353.353.5548 | Turkey | Asia | Western Asia |
| **SRR5193033** | 13/09/2016 | 1.2.3.18.353.353.5615 | Turkey | Asia | Western Asia |
| **SRR5215479** | 13/09/2016 | 1.2.3.18.353.353.5621 | Turkey | Asia | Western Asia |

| | | | | | |
|---|---|---|---|---|---|
| **SRR5194230** | 13/09/2016 | 1.2.3.18.353.353.5624 | Turkey | Asia | Western Asia |
| **SRR5193260** | 19/09/2016 | 1.2.3.18.353.353.5689 | Turkey | Asia | Western Asia |
| **SRR5216157** | 20/09/2016 | 1.2.3.18.353.353.5716 | Turkey | Asia | Western Asia |
| **SRR5194098** | 23/09/2016 | 1.2.3.18.353.353.5748 | Turkey | Asia | Western Asia |
| **SRR5193258** | 28/09/2016 | 1.2.3.18.353.353.5816 | Turkey | Asia | Western Asia |
| **SRR5220758** | 03/10/2016 | 1.2.3.18.353.353.5820 | Turkey | Asia | Western Asia |
| **SRR6919029** | 05/10/2016 | 1.2.3.18.353.353.5874 | Turkey | Asia | Western Asia |
| **SRR5193284** | 11/10/2016 | 1.2.3.18.353.353.5937 | Turkey | Asia | Western Asia |
| **SRR8658160** | 12/10/2016 | 1.2.3.18.353.353.5950 | Turkey | Asia | Western Asia |
| **SRR5216351** | 25/10/2016 | 1.2.3.18.353.353.6105 | Turkey | Asia | Western Asia |
| **SRR5194327** | 24/10/2016 | 1.2.3.18.353.353.6119 | Turkey | Asia | Western Asia |
| **SRR5584712** | 05/05/2017 | 1.2.3.18.353.353.7223 | Turkey | Asia | Western Asia |
| **SRR6899306** | 12/06/2017 | 1.2.3.18.353.353.7457 | Turkey | Asia | Western Asia |
| **SRR6922562** | 15/06/2017 | 1.2.3.18.353.353.7541 | Turkey | Asia | Western Asia |
| **SRR6898841** | 11/07/2017 | 1.2.3.18.353.353.7751 | Turkey | Asia | Western Asia |
| **SRR7842545** | 30/08/2018 | 1.2.3.18.353.353.7752 | Turkey | Asia | Western Asia |
| **SRR6919845** | 11/07/2017 | 1.2.3.18.353.353.7763 | Turkey | Asia | Western Asia |
| **SRR6898890** | 10/08/2017 | 1.2.3.18.353.353.7799 | Turkey | Asia | Western Asia |
| **SRR6900741** | 09/08/2017 | 1.2.3.18.353.353.7837 | Turkey | Asia | Western Asia |
| **SRR7511873** | 25/07/2017 | 1.2.3.18.353.353.7943 | Turkey | Asia | Western Asia |
| **SRR8553940** | 01/08/2017 | 1.2.3.18.353.353.7975 | Turkey | Asia | Western Asia |
| **SRR1967119** | 09/09/2014 | 1.2.3.18.353.353.800 | Turkey | Asia | Western Asia |
| **SRR6921986** | 14/07/2017 | 1.2.3.18.353.353.8029 | Turkey | Asia | Western Asia |
| **SRR6920604** | 02/08/2017 | 1.2.3.18.353.353.8109 | Turkey | Asia | Western Asia |
| **SRR6898382** | 17/08/2017 | 1.2.3.18.353.353.8136 | Turkey | Asia | Western Asia |
| **SRR6897659** | 04/09/2017 | 1.2.3.18.353.353.8273 | Turkey | Asia | Western Asia |
| **SRR6899430** | 06/09/2017 | 1.2.3.18.353.353.8299 | Turkey | Asia | Western Asia |
| **SRR6921980** | 08/09/2017 | 1.2.3.18.353.353.8331 | Turkey | Asia | Western Asia |
| **SRR6919818** | 08/09/2017 | 1.2.3.18.353.353.8336 | Turkey | Asia | Western Asia |
| **SRR6898381** | 12/09/2017 | 1.2.3.18.353.353.8368 | Turkey | Asia | Western Asia |
| **SRR8543185** | 14/09/2017 | 1.2.3.18.353.353.8378 | Turkey | Asia | Western Asia |
| **SRR6919210** | 21/09/2017 | 1.2.3.18.353.353.8460 | Turkey | Asia | Western Asia |
| **SRR6919141** | 21/09/2017 | 1.2.3.18.353.353.8462 | Turkey | Asia | Western Asia |
| **SRR6919243** | 22/09/2017 | 1.2.3.18.353.353.8463 | Turkey | Asia | Western Asia |
| **SRR6919326** | 25/09/2017 | 1.2.3.18.353.353.8482 | Turkey | Asia | Western Asia |
| **SRR6899265** | 27/09/2017 | 1.2.3.18.353.353.8526 | Turkey | Asia | Western Asia |
| **SRR6898187** | 04/10/2017 | 1.2.3.18.353.353.8601 | Turkey | Asia | Western Asia |
| **SRR6922727** | 10/10/2017 | 1.2.3.18.353.353.8626 | Turkey | Asia | Western Asia |
| **SRR6922787** | 11/10/2017 | 1.2.3.18.353.353.8639 | Turkey | Asia | Western Asia |
| **SRR6918577** | 06/10/2017 | 1.2.3.18.353.353.8652 | Turkey | Asia | Western Asia |
| **SRR6918317** | 17/10/2017 | 1.2.3.18.353.353.8670 | Turkey | Asia | Western Asia |
| **SRR6900165** | 17/10/2017 | 1.2.3.18.353.353.8671 | Turkey | Asia | Western Asia |
| **SRR6900140** | 12/10/2017 | 1.2.3.18.353.353.8685 | Turkey | Asia | Western Asia |
| **SRR6919309** | 27/10/2017 | 1.2.3.18.353.353.8730 | Turkey | Asia | Western Asia |
| **SRR6919924** | 25/10/2017 | 1.2.3.18.353.353.8745 | Turkey | Asia | Western Asia |
| **SRR6898940** | 30/10/2017 | 1.2.3.18.353.353.8752 | Turkey | Asia | Western Asia |
| **SRR1967648** | 08/07/2014 | 1.2.3.18.353.353.877 | Turkey | Asia | Western Asia |
| **SRR6919928** | 07/11/2017 | 1.2.3.18.353.353.8811 | Turkey | Asia | Western Asia |

| | | | | | |
|---|---|---|---|---|---|
| **SRR6919800** | 08/11/2017 | 1.2.3.18.353.353.8831 | Turkey | Asia | Western Asia |
| **SRR6918602** | 13/11/2017 | 1.2.3.18.353.353.8865 | Turkey | Asia | Western Asia |
| **SRR6922656** | 06/09/2017 | 1.2.3.18.353.353.9580 | Turkey | Asia | Western Asia |
| **SRR6900334** | 30/08/2017 | 1.2.3.18.353.353.9596 | Turkey | Asia | Western Asia |
| **SRR6900101** | 24/08/2017 | 1.2.3.18.353.353.9604 | Turkey | Asia | Western Asia |
| **SRR6922019** | 30/08/2017 | 1.2.3.18.353.353.9658 | Turkey | Asia | Western Asia |
| **SRR1968239** | 01/07/2014 | 1.2.3.18.353.361.363 | Turkey | Asia | Western Asia |
| **SRR7140655** | 21/08/2017 | 1.2.3.18.353.361.8007 | Turkey | Asia | Western Asia |
| **SRR6900732** | 28/06/2017 | 1.2.3.18.353.3917.7680 | Turkey | Asia | Western Asia |
| **SRR5220761** | 08/07/2015 | 1.2.3.18.353.400.3088 | Turkey | Asia | Western Asia |
| **SRR6921927** | 22/08/2017 | 1.2.3.18.353.4040.8148 | Turkey | Asia | Western Asia |
| **SRR1957979** | 15/10/2014 | 1.2.3.18.353.407.2638 | Turkey | Asia | Western Asia |
| **SRR1965564** | 18/07/2014 | 1.2.3.18.353.407.793 | Turkey | Asia | Western Asia |
| **SRR1958264** | 29/08/2014 | 1.2.3.18.353.407.919 | Turkey | Asia | Western Asia |
| **SRR6922525** | 26/09/2017 | 1.2.3.18.353.4145.8499 | Turkey | Asia | Western Asia |
| **SRR6900886** | 25/10/2017 | 1.2.3.18.353.4218.8731 | Turkey | Asia | Western Asia |
| **SRR1967945** | 04/09/2014 | 1.2.3.18.353.473.1025 | Turkey | Asia | Western Asia |
| **SRR8445034** | 30/05/2018 | 1.2.3.18.353.473.10338 | Turkey | Asia | Western Asia |
| **SRR7828532** | 30/07/2018 | 1.2.3.18.353.473.10542 | Turkey | Asia | Western Asia |
| **SRR7866912** | 29/08/2018 | 1.2.3.18.353.473.11544 | Turkey | Asia | Western Asia |
| **SRR7997123** | 26/09/2018 | 1.2.3.18.353.473.11845 | Turkey | Asia | Western Asia |
| **SRR3048515** | 22/08/2014 | 1.2.3.18.353.473.2092 | Turkey | Asia | Western Asia |
| **SRR7523819** | 29/07/2015 | 1.2.3.18.353.473.3207 | Turkey | Asia | Western Asia |
| **SRR7172565** | 11/08/2015 | 1.2.3.18.353.473.3237 | Turkey | Asia | Western Asia |
| **SRR5216323** | 29/07/2016 | 1.2.3.18.353.473.5056 | Turkey | Asia | Western Asia |
| **SRR5215844** | 06/09/2016 | 1.2.3.18.353.473.5124 | Turkey | Asia | Western Asia |
| **SRR6924058** | 16/08/2016 | 1.2.3.18.353.473.5145 | Turkey | Asia | Western Asia |
| **SRR1967607** | 16/09/2014 | 1.2.3.18.353.473.518 | Turkey | Asia | Western Asia |
| **SRR6900855** | 14/09/2016 | 1.2.3.18.353.473.5597 | Turkey | Asia | Western Asia |
| **SRR5193431** | 16/09/2016 | 1.2.3.18.353.473.5667 | Turkey | Asia | Western Asia |
| **SRR5216533** | 22/09/2016 | 1.2.3.18.353.473.5801 | Turkey | Asia | Western Asia |
| **SRR6918576** | 26/06/2017 | 1.2.3.18.353.473.7672 | Turkey | Asia | Western Asia |
| **SRR6897653** | 25/10/2017 | 1.2.3.18.353.473.8740 | Turkey | Asia | Western Asia |
| **SRR7444147** | 21/03/2018 | 1.2.3.18.353.473.9738 | Turkey | Asia | Western Asia |
| **SRR7402422** | 22/05/2018 | 1.2.3.18.353.473.9993 | Turkey | Asia | Western Asia |
| **SRR8369287** | 30/07/2018 | 1.2.3.18.353.4808.10536 | Turkey | Asia | Western Asia |
| **SRR8370528** | 24/07/2018 | 1.2.3.18.353.4880.10517 | Turkey | Asia | Western Asia |
| **SRR8292165** | 09/10/2018 | 1.2.3.18.353.4880.12133 | Turkey | Asia | Western Asia |
| **SRR1969194** | 19/05/2014 | 1.2.3.18.353.490.545 | Turkey | Asia | Western Asia |
| **SRR1967652** | 04/07/2014 | 1.2.3.18.353.494.552 | Turkey | Asia | Western Asia |
| **SRR1967506** | 01/07/2014 | 1.2.3.18.353.494.739 | Turkey | Asia | Western Asia |
| **SRR7879469** | 05/09/2018 | 1.2.3.18.353.5399.11564 | Turkey | Asia | Western Asia |
| **SRR7879493** | 04/09/2018 | 1.2.3.18.353.5422.11647 | Turkey | Asia | Western Asia |
| **SRR3049262** | 30/09/2014 | 1.2.3.18.353.555.1899 | Turkey | Asia | Western Asia |
| **SRR3048656** | 08/10/2014 | 1.2.3.18.353.555.710 | Turkey | Asia | Western Asia |
| **SRR8201853** | 26/10/2018 | 1.2.3.18.353.5619.12193 | Turkey | Asia | Western Asia |
| **SRR8485187** | 15/10/2018 | 1.2.3.18.353.5619.12196 | Turkey | Asia | Western Asia |
| **SRR8249766** | 09/11/2018 | 1.2.3.18.353.5701.12414 | Turkey | Asia | Western Asia |

| | | | | | |
|---|---|---|---|---|---|
| **SRR1966420** | 03/11/2014 | 1.2.3.18.353.613.2493 | Turkey | Asia | Western Asia |
| **SRR6896960** | 04/07/2017 | 1.2.3.18.353.613.7731 | Turkey | Asia | Western Asia |
| **SRR1968974** | 14/08/2014 | 1.2.3.18.353.634.1371 | Turkey | Asia | Western Asia |
| **SRR1966846** | 24/06/2014 | 1.2.3.18.353.659.801 | Turkey | Asia | Western Asia |
| **SRR5215866** | 06/08/2015 | 1.2.3.18.353.668.3259 | Turkey | Asia | Western Asia |
| **SRR1968346** | 16/09/2014 | 1.2.3.18.353.722.2054 | Turkey | Asia | Western Asia |
| **SRR1958270** | 09/07/2014 | 1.2.3.18.353.874.1115 | Turkey | Asia | Western Asia |
| **SRR1969315** | 08/07/2014 | 1.2.3.18.353.874.1209 | Turkey | Asia | Western Asia |
| **SRR5193976** | 17/08/2016 | 1.2.3.18.353.874.5183 | Turkey | Asia | Western Asia |
| **SRR8492366** | 23/05/2018 | 1.2.3.18.3576.4753.10046 | Turkey | Asia | Western Asia |
| **SRR6919925** | 26/09/2017 | 1.2.3.18.359.360.8271 | Turkey | Asia | Western Asia |
| **SRR7884520** | 31/07/2018 | 1.2.3.18.3644.4894.10555 | Turkey | Asia | Western Asia |
| **SRR8181939** | 29/10/2018 | 1.2.3.18.3644.4894.12322 | Turkey | Asia | Western Asia |
| **SRR1969353** | 03/07/2014 | 1.2.3.18.369.513.2127 | Turkey | Asia | Western Asia |
| **SRR7873992** | 20/08/2018 | 1.2.3.18.3852.5351.11430 | Turkey | Asia | Western Asia |
| **SRR8307289** | 16/08/2018 | 1.2.3.18.3856.5367.11473 | Turkey | Asia | Western Asia |
| **SRR3049585** | 03/06/2014 | 1.2.3.18.391.396.1704 | Turkey | Asia | Western Asia |
| **SRR1965392** | 02/07/2014 | 1.2.3.18.391.396.419 | Turkey | Asia | Western Asia |
| **SRR6918307** | 30/10/2017 | 1.30.673.2368.3241.4226.8764 | Turkey | Asia | Western Asia |
| **SRR8293804** | 06/09/2018 | 1.5.69.531.3873.5412.11623 | Turkey | Asia | Western Asia |
| **SRR8142783** | 17/09/2018 | 27.69.726.2566.3895.5450.117 55 | Turkey | Asia | Western Asia |
| **SRR5220893** | 14/06/2016 | 4.71.317.1292.2215.2729.4829 | Turkey | Asia | Western Asia |
| **SRR3286886** | 02/06/2015 | 1.1.2.1044.1704.1976.2889 | United Arab Emirates | Asia | Western Asia |
| **SRR7533386** | 30/05/2017 | 1.1.2.1044.1704.3782.7376 | United Arab Emirates | Asia | Western Asia |
| **SRR7351391** | 30/10/2015 | 1.1.2.1180.2004.2407.3967 | United Arab Emirates | Asia | Western Asia |
| **SRR8717124** | 07/10/2015 | 1.1.2.123.123.123.3771 | United Arab Emirates | Asia | Western Asia |
| **SRR5194273** | 12/01/2016 | 1.1.2.1326.2093.2543.4329 | United Arab Emirates | Asia | Western Asia |
| **SRR8717218** | 02/10/2015 | 1.1.2.1326.544.2309.3738 | United Arab Emirates | Asia | Western Asia |
| **SRR8724687** | 27/02/2019 | 1.1.2.1326.544.2643.12784 | United Arab Emirates | Asia | Western Asia |
| **SRR8758308** | 06/03/2019 | 1.1.2.1326.544.2643.12819 | United Arab Emirates | Asia | Western Asia |
| **SRR5194181** | 10/11/2016 | 1.1.2.1326.544.2643.6036 | United Arab Emirates | Asia | Western Asia |
| **SRR5194092** | 14/11/2016 | 1.1.2.1326.544.2643.6295 | United Arab Emirates | Asia | Western Asia |
| **SRR5193103** | 22/12/2016 | 1.1.2.1326.544.2643.6519 | United Arab Emirates | Asia | Western Asia |
| **SRR7892153** | 06/02/2018 | 1.1.2.1326.544.2643.9449 | United Arab Emirates | Asia | Western Asia |
| **SRR8509114** | 20/03/2018 | 1.1.2.1326.544.2643.9731 | United Arab Emirates | Asia | Western Asia |
| **SRR8293784** | 09/11/2018 | 1.1.2.1326.544.4631.12402 | United Arab Emirates | Asia | Western Asia |
| **SRR8503769** | 20/03/2018 | 1.1.2.1326.544.4631.9717 | United Arab Emirates | Asia | Western Asia |
| **SRR7221354** | 10/05/2018 | 1.1.2.1326.544.4717.9938 | United Arab Emirates | Asia | Western Asia |
| **SRR8292214** | 23/11/2018 | 1.1.2.1326.544.571.12485 | United Arab Emirates | Asia | Western Asia |
| **SRR3049031** | 10/06/2014 | 1.1.2.1326.544.571.2044 | United Arab Emirates | Asia | Western Asia |
| **SRR3286630** | 08/04/2015 | 1.1.2.1326.544.571.2190 | United Arab Emirates | Asia | Western Asia |

| | | | | | |
|---|---|---|---|---|---|
| **SRR1965216** | 31/03/2015 | 1.1.2.1326.544.571.2482 | United Arab Emirates | Asia | Western Asia |
| **SRR7257459** | 29/04/2015 | 1.1.2.1326.544.571.2748 | United Arab Emirates | Asia | Western Asia |
| **SRR5193653** | 01/05/2015 | 1.1.2.1326.544.571.2764 | United Arab Emirates | Asia | Western Asia |
| **SRR7538786** | 11/05/2015 | 1.1.2.1326.544.571.2790 | United Arab Emirates | Asia | Western Asia |
| **SRR7401974** | 09/06/2015 | 1.1.2.1326.544.571.2910 | United Arab Emirates | Asia | Western Asia |
| **SRR5193567** | 26/08/2015 | 1.1.2.1326.544.571.3362 | United Arab Emirates | Asia | Western Asia |
| **SRR8704702** | 10/11/2015 | 1.1.2.1326.544.571.4041 | United Arab Emirates | Asia | Western Asia |
| **SRR6900350** | 08/03/2016 | 1.1.2.1326.544.571.4480 | United Arab Emirates | Asia | Western Asia |
| **SRR5194008** | 06/07/2016 | 1.1.2.1326.544.571.4955 | United Arab Emirates | Asia | Western Asia |
| **SRR5193502** | 08/09/2016 | 1.1.2.1326.544.571.5369 | United Arab Emirates | Asia | Western Asia |
| **SRR5220548** | 24/10/2016 | 1.1.2.1326.544.571.6113 | United Arab Emirates | Asia | Western Asia |
| **SRR5215771** | 08/11/2016 | 1.1.2.1326.544.571.6232 | United Arab Emirates | Asia | Western Asia |
| **SRR1968228** | 14/01/2015 | 1.1.2.1326.544.571.675 | United Arab Emirates | Asia | Western Asia |
| **SRR6922675** | 27/06/2017 | 1.1.2.1326.544.571.7657 | United Arab Emirates | Asia | Western Asia |
| **SRR1963409** | 27/10/2014 | 1.1.2.1326.544.571.782 | United Arab Emirates | Asia | Western Asia |
| **SRR6898451** | 22/08/2017 | 1.1.2.1326.544.571.8184 | United Arab Emirates | Asia | Western Asia |
| **SRR7523698** | 24/04/2018 | 1.1.2.1326.544.571.9838 | United Arab Emirates | Asia | Western Asia |
| **SRR5215519** | 04/02/2016 | 1.1.2.1712.475.2575.4402 | United Arab Emirates | Asia | Western Asia |
| **SRR1969834** | 29/01/2015 | 1.1.2.178.178.1626.2223 | United Arab Emirates | Asia | Western Asia |
| **SRR6899329** | 02/03/2016 | 1.1.2.178.1855.2205.4468 | United Arab Emirates | Asia | Western Asia |
| **SRR5633244** | 06/03/2017 | 1.1.2.2244.2924.3675.7042 | United Arab Emirates | Asia | Western Asia |
| **SRR1960210** | 13/11/2014 | 1.1.2.243.243.243.1548 | United Arab Emirates | Asia | Western Asia |
| **SRR5215671** | 01/05/2015 | 1.1.2.274.1674.1936.2767 | United Arab Emirates | Asia | Western Asia |
| **SRR8724946** | 30/07/2015 | 1.1.2.274.274.2080.3194 | United Arab Emirates | Asia | Western Asia |
| **SRR8509111** | 16/04/2018 | 1.1.2.274.3539.4687.9823 | United Arab Emirates | Asia | Western Asia |
| **SRR1970115** | 15/10/2014 | 1.1.2.274.737.801.1003 | United Arab Emirates | Asia | Western Asia |
| **SRR8297307** | 28/08/2018 | 1.1.2.28.195.195.11517 | United Arab Emirates | Asia | Western Asia |
| **SRR3284679** | 25/08/2015 | 1.1.2.359.1682.1948.3377 | United Arab Emirates | Asia | Western Asia |
| **SRR1958102** | 07/11/2014 | 1.1.2.359.584.619.2295 | United Arab Emirates | Asia | Western Asia |
| **SRR6898391** | 04/01/2016 | 1.1.2.359.584.619.4070 | United Arab Emirates | Asia | Western Asia |
| **SRR1968779** | 19/08/2014 | 1.1.2.359.983.1104.1473 | United Arab Emirates | Asia | Western Asia |
| **SRR7257405** | 24/09/2015 | 1.1.2.359.983.1104.3601 | United Arab Emirates | Asia | Western Asia |
| **SRR5583115** | 30/03/2017 | 1.1.2.504.2340.2914.7130 | United Arab Emirates | Asia | Western Asia |
| **SRR6900347** | 17/08/2016 | 1.1.327.1316.2276.2825.5178 | United Arab Emirates | Asia | Western Asia |
| **SRR1967987** | 10/11/2014 | 1.2.3.18.1052.1184.1582 | United Arab Emirates | Asia | Western Asia |
| **SRR1960986** | 19/11/2014 | 1.2.3.18.1052.1184.1661 | United Arab Emirates | Asia | Western Asia |

| | | | | | |
|---|---|---|---|---|---|
| **SRR6919181** | 21/11/2017 | 1.2.3.18.175.175.8773 | United Arab Emirates | Asia | Western Asia |
| **SRR7285811** | 17/04/2018 | 1.2.3.18.180.180.10883 | United Arab Emirates | Asia | Western Asia |
| **SRR8503975** | 17/04/2018 | 1.2.3.18.180.180.10910 | United Arab Emirates | Asia | Western Asia |
| **SRR7892259** | 17/07/2018 | 1.2.3.18.353.353.10454 | United Arab Emirates | Asia | Western Asia |
| **SRR6919491** | 17/11/2017 | 1.2.3.18.353.353.8761 | United Arab Emirates | Asia | Western Asia |
| **SRR8367057** | 11/12/2018 | 1.2.3.18.353.5619.12196 | United Arab Emirates | Asia | Western Asia |
| **SRR8419416** | 28/12/2018 | 1.2.3.18.353.5619.12506 | United Arab Emirates | Asia | Western Asia |
| **SRR8292196** | 27/11/2018 | 1.2.3.18.4050.5721.12484 | United Arab Emirates | Asia | Western Asia |
| **SRR8464950** | 08/01/2019 | 1.3.249.2665.4078.5775.12635 | United Arab Emirates | Asia | Western Asia |
| **SRR8116967** | 16/10/2018 | 17.38.223.2647.4026.5659.12195 | United Arab Emirates | Asia | Western Asia |
| **SRR7841583** | 03/07/2018 | 1.1.2.12.12.12.10421 | United states | Americas | Northern America |
| **SRR7444201** | 19/06/2018 | 1.1.2.15.361.1950.10141 | United states | Americas | Northern America |
| **SRR6918568** | 19/07/2017 | 1.1.2.15.361.3993.8011 | United states | Americas | Northern America |
| **SRR1969057** | 15/07/2014 | 1.1.2.28.1310.1496.2033 | United states | Americas | Northern America |
| **SRR8201772** | 27/09/2018 | 1.1.2.28.149.149.11775 | United states | Americas | Northern America |
| **SRR8106765** | 24/09/2018 | 1.1.2.28.149.149.11821 | United states | Americas | Northern America |
| **SRR7292956** | 08/07/2016 | 1.1.2.28.195.195.4963 | United states | Americas | Northern America |
| **SRR5194192** | 08/06/2016 | 1.1.2.61.61.561.4796 | United states | Americas | Northern America |
| **SRR5215744** | 19/01/2016 | 1.1.2.61.61.61.4356 | United states | Americas | Northern America |
| **SRR1957872** | 29/10/2014 | 1.1.2.61.61.701.857 | United states | Americas | Northern America |
| **SRR5220972** | 30/08/2016 | 1.1.333.103.103.548.5359 | United states | Americas | Northern America |
| **SRR6897709** | 23/05/2017 | 1.2.3.18.175.175.7339 | United states | Americas | Northern America |
| **SRR8484210** | 06/11/2018 | 1.2.3.18.180.180.10985 | United states | Americas | Northern America |
| **SRR1969471** | 07/07/2014 | 1.2.3.18.3658.4934.10650 | United states | Americas | Northern America |
| **SRR1957719** | 24/06/2014 | 1.5.61.118.145.145.145 | United states | Americas | Northern America |
| **SRR7879494** | 10/07/2018 | 1.5.638.2256.3620.4845.10392 | United states | Americas | Northern America |
| **SRR7350642** | 21/09/2015 | 1.5.69.531.1861.2213.3499 | United states | Americas | Northern America |
| **SRR3048690** | 25/04/2014 | 1.1.2.224.224.224.224 | Vietnam | Asia | South-eastern Asia |
| **SRR6919153** | 11/04/2017 | 1.1.2.242.242.3710.7160 | Vietnam | Asia | South-eastern Asia |
| **SRR3286610** | 08/04/2015 | 1.1.2.53.780.855.1090 | Vietnam | Asia | South-eastern Asia |
| **SRR5220289** | 24/08/2016 | 1.1.2.53.780.855.5217 | Vietnam | Asia | South-eastern Asia |
| **SRR6901081** | 02/09/2016 | 1.1.2.53.808.2935.5342 | Vietnam | Asia | South-eastern Asia |
| **SRR1961943** | 08/12/2014 | 1.1.2.7.1178.1338.1803 | Vietnam | Asia | South-eastern Asia |
| **SRR6922101** | 18/12/2017 | 1.1.2.7.1178.1338.9041 | Vietnam | Asia | South-eastern Asia |
| **SRR8820634** | 18/03/2019 | 1.1.2.7.1178.4664.12861 | Vietnam | Asia | South-eastern Asia |
| **SRR7850527** | 12/07/2018 | 1.1.2.7.1178.4713.10430 | Vietnam | Asia | South-eastern Asia |
| **SRR7892221** | 15/05/2018 | 1.1.2.7.1178.4713.9961 | Vietnam | Asia | South-eastern Asia |
| **SRR1960134** | 30/12/2014 | 1.1.2.849.1297.1481.2012 | Vietnam | Asia | South-eastern Asia |
| **SRR5220879** | 30/04/2015 | 1.1.2.849.1672.1934.2761 | Vietnam | Asia | South-eastern Asia |

| | | | | | |
|---|---|---|---|---|---|
| **SRR7275070** | 14/03/2018 | 1.1.685.2420.3350.4419.9706 | Vietnam | Asia | South-eastern Asia |
| **SRR6898922** | 04/10/2016 | 1.2.3.18.323.2409.5713 | Vietnam | Asia | South-eastern Asia |
| **SRR6919108** | 16/05/2017 | 4.108.639.2257.2970.3763.7308 | Vietnam | Asia | South-eastern Asia |

### Table S4. Validation sample list and associated metadata.

| Accession | Collection Country | Model Classification | Collection Date | Collection Body | Reference |
|---|---|---|---|---|---|
| **ERR2278734** | Poland | Hungary | 2016 | National Veterinary Research Institute | NA |
| **ERR2278723** | Poland | Poland | 2016 | National Veterinary Research Institute | NA |
| **ERR2278728** | Poland | Poland | 2016 | National Veterinary Research Institute | NA |
| **ERR2278729** | Poland | Poland | 2016 | National Veterinary Research Institute | NA |
| **ERR2278739** | Poland | Poland | 2016 | National Veterinary Research Institute | NA |
| **ERR2278742** | Poland | Poland | 2016 | National Veterinary Research Institute | NA |
| **ERR2278743** | Poland | Poland | 2016 | National Veterinary Research Institute | NA |
| **ERR2278749** | Poland | Poland | 2016 | National Veterinary Research Institute | NA |
| **ERR2278713** | Poland | Poland | 2016 | National Veterinary Research Institute | NA |
| **ERR2278718** | Poland | Poland | 2016 | National Veterinary Research Institute | NA |
| **ERR2278724** | Poland | Poland | 2016 | National Veterinary Research Institute | NA |
| **ERR2278738** | Poland | Poland | 2016 | National Veterinary Research Institute | NA |
| **ERR2278745** | Poland | Poland | 2016 | National Veterinary Research Institute | NA |
| **ERR2278715** | Poland | Poland | 2016 | National Veterinary Research Institute | NA |
| **ERR2278722** | Poland | Poland | 2016 | National Veterinary Research Institute | NA |
| **ERR2278725** | Poland | Poland | 2016 | National Veterinary Research Institute | NA |
| **ERR2278740** | Poland | Poland | 2016 | National Veterinary Research Institute | NA |
| **ERR2278744** | Poland | Poland | 2016 | National Veterinary Research Institute | NA |
| **ERR2278748** | Poland | Poland | 2016 | National Veterinary Research Institute | NA |
| **ERR2278721** | Poland | Spain | 2016 | National Veterinary Research Institute | NA |
| **ERR2278733** | Poland | Spain | 2016 | National Veterinary Research Institute | NA |
| **ERR2278717** | Poland | Spain | 2016 | National Veterinary Research Institute | NA |
| **ERR2278720** | Poland | Spain | 2016 | National Veterinary Research Institute | NA |
| **ERR2278726** | Poland | Spain | 2015 | National Veterinary Research Institute | NA |
| **ERR2278727** | Poland | Spain | 2016 | National Veterinary Research Institute | NA |
| **ERR2278735** | Poland | Spain | 2016 | National Veterinary Research Institute | NA |
| **ERR2278716** | Poland | Spain | 2016 | National Veterinary Research Institute | NA |
| **ERR2278736** | Poland | Spain | 2016 | National Veterinary Research Institute | NA |
| **ERR2278731** | Poland | Spain | 2016 | National Veterinary Research Institute | NA |
| **ERR2278741** | Poland | Spain | 2016 | National Veterinary Research Institute | NA |
| **ERR2278714** | Poland | Spain | 2016 | National Veterinary Research Institute | NA |
| **ERR2278719** | Poland | Spain | 2016 | National Veterinary Research Institute | NA |
| **ERR2278737** | Poland | Spain | 2016 | National Veterinary Research Institute | NA |
| **ERR2278746** | Poland | Spain | 2017 | National Veterinary Research Institute | NA |
| **ERR2278747** | Poland | Turkey | 2017 | National Veterinary Research Institute | NA |
| **SRR7777155** | Singapore | Indonesia | 2015 | Ministry Of Health | NA |
| **SRR7777164** | Singapore | Indonesia | 2014 | Ministry Of Health | NA |
| **SRR7777165** | Singapore | Malaysia | 2016 | Ministry Of Health | NA |
| **SRR7777161** | Singapore | Singapore | 2014 | Ministry Of Health | NA |
| **SRR7777162** | Singapore | Singapore | 2014 | Ministry Of Health | NA |

| | | | | | |
|---|---|---|---|---|---|
| **SRR7777163** | Singapore | Singapore | 2014 | Ministry Of Health | NA |
| **SRR7777172** | Singapore | Singapore | 2016 | Ministry Of Health | NA |
| **SRR7777167** | Singapore | Singapore | 2016 | Ministry Of Health | NA |
| **SRR7777170** | Singapore | Singapore | 2016 | Ministry Of Health | NA |
| **SRR7777178** | Singapore | Singapore | 2014 | Ministry Of Health | NA |
| **SRR7777184** | Singapore | Singapore | 2016 | Ministry Of Health | NA |
| **SRR7777200** | Singapore | Singapore | 2017 | Ministry Of Health | NA |
| **SRR7777175** | Singapore | Singapore | 2015 | Ministry Of Health | NA |
| **SRR7777183** | Singapore | Singapore | 2016 | Ministry Of Health | NA |
| **SRR7777202** | Singapore | Singapore | 2017 | Ministry Of Health | NA |
| **SRR7777212** | Singapore | Singapore | 2016 | Ministry Of Health | NA |
| **SRR7777215** | Singapore | Singapore | 2016 | Ministry Of Health | NA |
| **SRR7777218** | Singapore | Singapore | 2016 | Ministry Of Health | NA |
| **SRR7777223** | Singapore | Singapore | 2016 | Ministry Of Health | NA |
| **SRR7777206** | Singapore | Singapore | 2016 | Ministry Of Health | NA |
| **SRR7777207** | Singapore | Singapore | 2016 | Ministry Of Health | NA |
| **SRR7777222** | Singapore | Singapore | 2016 | Ministry Of Health | NA |
| **SRR7777150** | Singapore | Singapore | 2014 | Ministry Of Health | NA |
| **SRR7777166** | Singapore | Singapore | 2016 | Ministry Of Health | NA |
| **SRR7777171** | Singapore | Singapore | 2016 | Ministry Of Health | NA |
| **SRR7777185** | Singapore | Singapore | 2016 | Ministry Of Health | NA |
| **SRR7777193** | Singapore | Singapore | 2014 | Ministry Of Health | NA |
| **SRR7777194** | Singapore | Singapore | 2014 | Ministry Of Health | NA |
| **SRR7777208** | Singapore | Singapore | 2016 | Ministry Of Health | NA |
| **SRR7777209** | Singapore | Singapore | 2016 | Ministry Of Health | NA |
| **SRR7777211** | Singapore | Singapore | 2015 | Ministry Of Health | NA |
| **SRR7777214** | Singapore | Singapore | 2017 | Ministry Of Health | NA |
| **SRR7777217** | Singapore | Singapore | 2016 | Ministry Of Health | NA |
| **SRR7777219** | Singapore | Singapore | 2016 | Ministry Of Health | NA |
| **SRR7777220** | Singapore | Singapore | 2016 | Ministry Of Health | NA |
| **SRR7777224** | Singapore | Singapore | 2016 | Ministry Of Health | NA |
| **SRR7777169** | Singapore | Singapore | 2016 | Ministry Of Health | NA |
| **SRR7777173** | Singapore | Singapore | 2016 | Ministry Of Health | NA |
| **SRR7777176** | Singapore | Singapore | 2016 | Ministry Of Health | NA |
| **SRR7777180** | Singapore | Singapore | 2016 | Ministry Of Health | NA |
| **SRR7777182** | Singapore | Singapore | 2016 | Ministry Of Health | NA |
| **SRR7777190** | Singapore | Singapore | 2016 | Ministry Of Health | NA |
| **SRR7777204** | Singapore | Singapore | 2017 | Ministry Of Health | NA |
| **SRR7777205** | Singapore | Singapore | 2015 | Ministry Of Health | NA |
| **SRR7777213** | Singapore | Singapore | 2016 | Ministry Of Health | NA |
| **SRR7777216** | Singapore | Singapore | 2016 | Ministry Of Health | NA |
| **SRR7777221** | Singapore | Singapore | 2016 | Ministry Of Health | NA |
| **SRR7777160** | Singapore | Thailand | 2014 | Ministry Of Health | NA |
| **SRR12223838** | South Africa | South africa | 2018 | National Institute for Communicable Diseases | NA |
| **SRR12223839** | South Africa | South africa | 2018 | National Institute for Communicable Diseases | NA |
| **SRR12223842** | South Africa | South africa | 2018 | National Institute for Communicable Diseases | NA |
| **SRR12223843** | South Africa | South africa | 2018 | National Institute for Communicable Diseases | NA |
| **SRR12223846** | South Africa | South africa | 2018 | National Institute for Communicable Diseases | NA |
| **SRR12223852** | South Africa | South africa | 2018 | National Institute for Communicable Diseases | NA |
| **SRR12223853** | South Africa | South africa | 2018 | National Institute for Communicable Diseases | NA |
| **SRR12223831** | South Africa | South africa | 2018 | National Institute for Communicable Diseases | NA |
| **SRR12223834** | South Africa | South africa | 2018 | National Institute for Communicable Diseases | NA |
| **SRR12223836** | South Africa | South africa | 2018 | National Institute for Communicable Diseases | NA |
| **SRR12223837** | South Africa | South africa | 2018 | National Institute for Communicable Diseases | NA |
| **SRR12223840** | South Africa | South africa | 2018 | National Institute for | NA |

| | | | | | |
|---|---|---|---|---|---|
| | | | | Communicable Diseases | |
| **SRR12223832** | South Africa | South africa | 2018 | National Institute for Communicable Diseases | NA |
| **SRR12223841** | South Africa | South africa | 2018 | National Institute for Communicable Diseases | NA |
| **SRR12223849** | South Africa | South africa | 2018 | National Institute for Communicable Diseases | NA |
| **SRR12223851** | South Africa | South africa | 2018 | National Institute for Communicable Diseases | NA |
| **SRR12223854** | South Africa | South africa | 2018 | National Institute for Communicable Diseases | NA |
| **SRR12223844** | South Africa | South africa | 2018 | National Institute for Communicable Diseases | NA |
| **SRR12223845** | South Africa | South africa | 2018 | National Institute for Communicable Diseases | NA |
| **SRR12223847** | South Africa | South africa | 2018 | National Institute for Communicable Diseases | NA |
| **SRR12223850** | South Africa | South africa | 2018 | National Institute for Communicable Diseases | NA |
| **SRR12223855** | South Africa | South africa | 2018 | National Institute for Communicable Diseases | NA |
| **SRR12223833** | South Africa | South africa | 2018 | National Institute for Communicable Diseases | NA |
| **SRR12223835** | South Africa | South africa | 2018 | National Institute for Communicable Diseases | NA |
| **SRR12223848** | South Africa | South africa | 2018 | National Institute for Communicable Diseases | NA |
| **SRR7523733** | Poland | Spain | 25/05/18 | UKHSA/PHE | Pijnacker et al., 2019 |
| **SRR7350599** | Poland | Spain | 30/05/18 | UKHSA/PHE | Pijnacker et al., 2019 |
| **SRR8492323** | Poland | Spain | 21/06/18 | UKHSA/PHE | Pijnacker et al., 2019 |
| **SRR8204740** | Poland | Spain | 10/10/18 | UKHSA/PHE | Pijnacker et al., 2019 |
| **SRR7841505** | Poland | Spain | 31/07/18 | UKHSA/PHE | Pijnacker et al., 2019 |
| **SRR8367067** | Poland | Spain | 03/08/18 | UKHSA/PHE | Pijnacker et al., 2019 |
| **SRR7850650** | Poland | Spain | 30/08/18 | UKHSA/PHE | Pijnacker et al., 2019 |
| **SRR7850533** | Poland | Spain | 04/09/18 | UKHSA/PHE | Pijnacker et al., 2019 |
| **SRR7892168** | Poland | Spain | 07/09/18 | UKHSA/PHE | Pijnacker et al., 2019 |
| **SRR7890472** | Poland | Spain | 10/09/18 | UKHSA/PHE | Pijnacker et al., 2019 |
| **SRR7885266** | Poland | Spain | 10/09/18 | UKHSA/PHE | Pijnacker et al., 2019 |
| **SRR8137417** | Poland | Spain | 12/09/18 | UKHSA/PHE | Pijnacker et al., 2019 |
| **SRR8131528** | Poland | Spain | 14/09/18 | UKHSA/PHE | Pijnacker et al., 2019 |
| **SRR8114905** | Poland | Spain | 25/09/18 | UKHSA/PHE | Pijnacker et al., 2019 |
| **SRR7910360** | Poland | Spain | 10/09/18 | UKHSA/PHE | Pijnacker et al., 2019 |
| **SRR8131506** | Poland | Spain | 13/09/18 | UKHSA/PHE | Pijnacker et al., 2019 |
| **SRR7962058** | Poland | Spain | 18/09/18 | UKHSA/PHE | Pijnacker et al., 2019 |
| **SRR7998203** | Poland | Spain | 24/09/18 | UKHSA/PHE | Pijnacker et al., 2019 |
| **SRR8097951** | Poland | Spain | 27/09/18 | UKHSA/PHE | Pijnacker et al., 2019 |
| **SRR8097943** | Poland | Spain | 27/09/18 | UKHSA/PHE | Pijnacker et al., 2019 |
| **SRR8097924** | Poland | Spain | 27/09/18 | UKHSA/PHE | Pijnacker et al., 2019 |
| **SRR8149423** | Poland | Spain | 28/09/18 | UKHSA/PHE | Pijnacker et al., 2019 |
| **SRR8149431** | Poland | Spain | 24/10/18 | UKHSA/PHE | Pijnacker et al., 2019 |
| **SRR4063746** | Poland | Spain | 22/07/15 | UKHSA/PHE | Pijnacker et al., 2019 |
| **SRR4063750** | Poland | Portugal | 03/12/15 | UKHSA/PHE | Pijnacker et al., 2019 |
| **SRR4063723** | Poland | Portugal | 11/12/15 | UKHSA/PHE | Pijnacker et al., 2019 |
| **SRR4063755** | Poland | Spain | 18/12/15 | UKHSA/PHE | Pijnacker et al., 2019 |
| **SRR5215891** | Poland | Portugal | 24/07/15 | UKHSA/PHE | Pijnacker et al., 2019 |
| **SRR5220233** | Poland | Portugal | 03/08/15 | UKHSA/PHE | Pijnacker et al., 2019 |
| **SRR7538879** | Poland | Spain | 28/07/15 | UKHSA/PHE | Pijnacker et al., 2019 |
| **SRR4063738** | Poland | Portugal | 04/08/15 | UKHSA/PHE | Pijnacker et al., 2019 |
| **SRR4063747** | Poland | Spain | 07/08/15 | UKHSA/PHE | Pijnacker et al., 2019 |
| **SRR4063715** | Poland | Spain | 19/08/15 | UKHSA/PHE | Pijnacker et al., 2019 |
| **SRR3284815** | Poland | Spain | 27/11/15 | UKHSA/PHE | Pijnacker et al., 2019 |
| **SRR6901054** | Poland | Spain | 30/12/15 | UKHSA/PHE | Pijnacker et al., 2019 |
| **SRR4063752** | Poland | Spain | 12/01/16 | UKHSA/PHE | Pijnacker et al., 2019 |
| **SRR4063699** | Poland | Spain | 22/01/16 | UKHSA/PHE | Pijnacker et al., 2019 |
| **SRR4063700** | Poland | Spain | 09/02/16 | UKHSA/PHE | Pijnacker et al., 2019 |
| **SRR6900097** | Poland | Spain | 15/09/17 | UKHSA/PHE | Pijnacker et al., 2019 |
| **SRR6899258** | Poland | Spain | 19/09/17 | UKHSA/PHE | Pijnacker et al., 2019 |

| | | | | | |
|---|---|---|---|---|---|
| **SRR6921964** | Poland | Spain | 27/09/17 | UKHSA/PHE | Pijnacker et al., 2019 |
| **SRR7523138** | Poland | Spain | 26/06/18 | UKHSA/PHE | Pijnacker et al., 2019 |
| **SRR7997085** | Poland | Spain | 21/09/18 | UKHSA/PHE | Pijnacker et al., 2019 |
| **SRR5193132** | Poland | Spain | 05/12/16 | UKHSA/PHE | Pijnacker et al., 2019 |
| **SRR6922690** | Poland | Spain | 21/12/16 | UKHSA/PHE | Pijnacker et al., 2019 |
| **SRR5584922** | Poland | Portugal | 10/04/17 | UKHSA/PHE | Pijnacker et al., 2019 |
| **SRR6900878** | Poland | Spain | 19/07/17 | UKHSA/PHE | Pijnacker et al., 2019 |
| **SRR6920541** | Poland | Portugal | 31/05/17 | UKHSA/PHE | Pijnacker et al., 2019 |
| **SRR6922652** | Poland | Spain | 15/05/17 | UKHSA/PHE | Pijnacker et al., 2019 |
| **SRR6918861** | Poland | Spain | 22/05/17 | UKHSA/PHE | Pijnacker et al., 2019 |
| **SRR6900984** | Poland | Portugal | 24/05/17 | UKHSA/PHE | Pijnacker et al., 2019 |
| **SRR6898839** | Poland | Spain | 19/05/17 | UKHSA/PHE | Pijnacker et al., 2019 |
| **SRR6898941** | Poland | Spain | 30/05/17 | UKHSA/PHE | Pijnacker et al., 2019 |
| **SRR6922699** | Poland | Portugal | 02/06/17 | UKHSA/PHE | Pijnacker et al., 2019 |
| **SRR6898404** | Poland | Spain | 12/07/17 | UKHSA/PHE | Pijnacker et al., 2019 |
| **SRR6920168** | Poland | Portugal | 02/08/17 | UKHSA/PHE | Pijnacker et al., 2019 |
| **SRR6922068** | Poland | Portugal | 18/07/17 | UKHSA/PHE | Pijnacker et al., 2019 |
| **SRR6919917** | Poland | Portugal | 18/07/17 | UKHSA/PHE | Pijnacker et al., 2019 |
| **SRR6922020** | Poland | Spain | 19/07/17 | UKHSA/PHE | Pijnacker et al., 2019 |
| **SRR6897980** | Poland | Spain | 20/07/17 | UKHSA/PHE | Pijnacker et al., 2019 |
| **SRR6922660** | Poland | Spain | 24/07/17 | UKHSA/PHE | Pijnacker et al., 2019 |
| **SRR6918604** | Poland | Spain | 28/07/17 | UKHSA/PHE | Pijnacker et al., 2019 |
| **SRR6898007** | Poland | Spain | 28/07/17 | UKHSA/PHE | Pijnacker et al., 2019 |
| **SRR6897870** | Poland | Portugal | 04/08/17 | UKHSA/PHE | Pijnacker et al., 2019 |
| **SRR6898121** | Poland | Spain | 08/08/17 | UKHSA/PHE | Pijnacker et al., 2019 |
| **SRR6918299** | Poland | Spain | 02/08/17 | UKHSA/PHE | Pijnacker et al., 2019 |
| **SRR6898018** | Poland | Spain | 01/09/17 | UKHSA/PHE | Pijnacker et al., 2019 |
| **SRR6919991** | Poland | Portugal | 04/09/17 | UKHSA/PHE | Pijnacker et al., 2019 |
| **SRR6899374** | Poland | Spain | 04/09/17 | UKHSA/PHE | Pijnacker et al., 2019 |
| **SRR6919914** | Poland | Spain | 06/09/17 | UKHSA/PHE | Pijnacker et al., 2019 |
| **SRR6901178** | Poland | Spain | 06/09/17 | UKHSA/PHE | Pijnacker et al., 2019 |
| **SRR6897886** | Poland | Spain | 06/09/17 | UKHSA/PHE | Pijnacker et al., 2019 |
| **SRR6901062** | Poland | Spain | 07/09/17 | UKHSA/PHE | Pijnacker et al., 2019 |
| **SRR6898821** | Poland | Spain | 11/09/17 | UKHSA/PHE | Pijnacker et al., 2019 |
| **SRR6919951** | Poland | Spain | 12/09/17 | UKHSA/PHE | Pijnacker et al., 2019 |
| **SRR6924113** | Poland | Spain | 13/09/17 | UKHSA/PHE | Pijnacker et al., 2019 |
| **SRR6919112** | Poland | Spain | 13/09/17 | UKHSA/PHE | Pijnacker et al., 2019 |
| **SRR6918852** | Poland | Spain | 13/09/17 | UKHSA/PHE | Pijnacker et al., 2019 |
| **SRR6898438** | Poland | Spain | 13/09/17 | UKHSA/PHE | Pijnacker et al., 2019 |
| **SRR6898849** | Poland | Spain | 15/09/17 | UKHSA/PHE | Pijnacker et al., 2019 |
| **SRR6897054** | Poland | Spain | 15/09/17 | UKHSA/PHE | Pijnacker et al., 2019 |
| **SRR6924084** | Poland | Spain | 20/09/17 | UKHSA/PHE | Pijnacker et al., 2019 |
| **SRR6922669** | Poland | Spain | 27/09/17 | UKHSA/PHE | Pijnacker et al., 2019 |
| **SRR6900193** | Poland | Greece | 08/09/17 | UKHSA/PHE | Pijnacker et al., 2019 |
| **SRR6922607** | Poland | Spain | 12/09/17 | UKHSA/PHE | Pijnacker et al., 2019 |
| **SRR6900871** | Poland | Spain | 15/09/17 | UKHSA/PHE | Pijnacker et al., 2019 |
| **SRR6918644** | Poland | Spain | 20/09/17 | UKHSA/PHE | Pijnacker et al., 2019 |
| **SRR6919808** | Poland | Spain | 15/09/17 | UKHSA/PHE | Pijnacker et al., 2019 |
| **SRR6897051** | Poland | Spain | 15/09/17 | UKHSA/PHE | Pijnacker et al., 2019 |
| **SRR6919100** | Poland | Spain | 15/09/17 | UKHSA/PHE | Pijnacker et al., 2019 |
| **SRR6919796** | Poland | Spain | 12/09/17 | UKHSA/PHE | Pijnacker et al., 2019 |
| **SRR6920097** | Poland | Spain | 12/09/17 | UKHSA/PHE | Pijnacker et al., 2019 |
| **SRR6900909** | Poland | Spain | 19/09/17 | UKHSA/PHE | Pijnacker et al., 2019 |
| **SRR6924076** | Poland | Spain | 20/09/17 | UKHSA/PHE | Pijnacker et al., 2019 |
| **SRR6899438** | Poland | Spain | 09/10/17 | UKHSA/PHE | Pijnacker et al., 2019 |
| **SRR6924133** | Poland | Portugal | 17/10/17 | UKHSA/PHE | Pijnacker et al., 2019 |
| **SRR6900887** | Poland | Spain | 06/10/17 | UKHSA/PHE | Pijnacker et al., 2019 |
| **SRR6919293** | Poland | Spain | 12/10/17 | UKHSA/PHE | Pijnacker et al., 2019 |
| **SRR6918294** | Poland | Spain | 04/12/17 | UKHSA/PHE | Pijnacker et al., 2019 |
| **SRR6898422** | Poland | Spain | 30/08/17 | UKHSA/PHE | Pijnacker et al., 2019 |

| | | | | | |
|---|---|---|---|---|---|
| **SRR6898083** | Poland | Spain | 06/09/17 | UKHSA/PHE | Pijnacker et al., 2019 |
| **SRR7351458** | Poland | Portugal | 24/04/18 | UKHSA/PHE | Pijnacker et al., 2019 |
| **SRR8499000** | Poland | Spain | 08/05/18 | UKHSA/PHE | Pijnacker et al., 2019 |
| **SRR8515670** | Poland | Portugal | 09/05/18 | UKHSA/PHE | Pijnacker et al., 2019 |
| **SRR7516466** | Poland | Portugal | 09/05/18 | UKHSA/PHE | Pijnacker et al., 2019 |
| **SRR7416093** | Poland | Portugal | 09/05/18 | UKHSA/PHE | Pijnacker et al., 2019 |
| **SRR7278047** | Poland | Portugal | 09/05/18 | UKHSA/PHE | Pijnacker et al., 2019 |
| **SRR8509115** | Poland | Spain | 10/05/18 | UKHSA/PHE | Pijnacker et al., 2019 |
| **SRR8503803** | Poland | Spain | 10/05/18 | UKHSA/PHE | Pijnacker et al., 2019 |
| **SRR8499035** | Poland | Spain | 10/05/18 | UKHSA/PHE | Pijnacker et al., 2019 |
| **SRR7873952** | Poland | Spain | 10/05/18 | UKHSA/PHE | Pijnacker et al., 2019 |
| **SRR7456768** | Poland | Spain | 10/05/18 | UKHSA/PHE | Pijnacker et al., 2019 |
| **SRR7439595** | Poland | Portugal | 10/05/18 | UKHSA/PHE | Pijnacker et al., 2019 |
| **SRR7249838** | Poland | Spain | 10/05/18 | UKHSA/PHE | Pijnacker et al., 2019 |
| **SRR7359066** | Poland | Spain | 15/05/18 | UKHSA/PHE | Pijnacker et al., 2019 |
| **SRR7350588** | Poland | Portugal | 15/05/18 | UKHSA/PHE | Pijnacker et al., 2019 |
| **SRR8499138** | Poland | Spain | 16/05/18 | UKHSA/PHE | Pijnacker et al., 2019 |
| **SRR7456711** | Poland | Portugal | 16/05/18 | UKHSA/PHE | Pijnacker et al., 2019 |
| **SRR7344650** | Poland | Portugal | 16/05/18 | UKHSA/PHE | Pijnacker et al., 2019 |
| **SRR7297922** | Poland | Spain | 16/05/18 | UKHSA/PHE | Pijnacker et al., 2019 |
| **SRR7275015** | Poland | Portugal | 16/05/18 | UKHSA/PHE | Pijnacker et al., 2019 |
| **SRR7237430** | Poland | Spain | 16/05/18 | UKHSA/PHE | Pijnacker et al., 2019 |
| **SRR7223242** | Poland | Portugal | 16/05/18 | UKHSA/PHE | Pijnacker et al., 2019 |
| **SRR8508664** | Poland | Spain | 22/05/18 | UKHSA/PHE | Pijnacker et al., 2019 |
| **SRR7469150** | Poland | Spain | 22/05/18 | UKHSA/PHE | Pijnacker et al., 2019 |
| **SRR7415099** | Poland | Spain | 10/05/18 | UKHSA/PHE | Pijnacker et al., 2019 |
| **SRR8498997** | Poland | Spain | 08/05/18 | UKHSA/PHE | Pijnacker et al., 2019 |
| **SRR8499137** | Poland | Spain | 11/05/18 | UKHSA/PHE | Pijnacker et al., 2019 |
| **SRR8499334** | Poland | Spain | 15/05/18 | UKHSA/PHE | Pijnacker et al., 2019 |
| **SRR7416295** | Poland | Portugal | 17/05/18 | UKHSA/PHE | Pijnacker et al., 2019 |
| **SRR7292910** | Poland | Spain | 22/05/18 | UKHSA/PHE | Pijnacker et al., 2019 |
| **SRR1957942** | Spain | Spain | 04/10/14 | UKHSA/PHE | Inns et al., 2016 |
| **SRR1958017** | Spain | Spain | 15/09/14 | UKHSA/PHE | Inns et al., 2016 |
| **SRR1958156** | Spain | Spain | 03/07/14 | UKHSA/PHE | Inns et al., 2016 |
| **SRR1960253** | Spain | Spain | 30/12/14 | UKHSA/PHE | Inns et al., 2016 |
| **SRR1966021** | Spain | Spain | 03/03/15 | UKHSA/PHE | Inns et al., 2016 |
| **SRR1966200** | Spain | Spain | 22/01/15 | UKHSA/PHE | Inns et al., 2016 |
| **SRR1966421** | Spain | Spain | 09/03/15 | UKHSA/PHE | Inns et al., 2016 |
| **SRR1966697** | Spain | Spain | 19/03/15 | UKHSA/PHE | Inns et al., 2016 |
| **SRR1967334** | Spain | Spain | 15/04/14 | UKHSA/PHE | Inns et al., 2016 |
| **SRR1967401** | Spain | Spain | 31/03/14 | UKHSA/PHE | Inns et al., 2016 |
| **SRR1967425** | Spain | Spain | 17/06/14 | UKHSA/PHE | Inns et al., 2016 |
| **SRR1967498** | Spain | Spain | 15/03/15 | UKHSA/PHE | Inns et al., 2016 |
| **SRR1967559** | Spain | Spain | 18/03/15 | UKHSA/PHE | Inns et al., 2016 |
| **SRR1967568** | Spain | Spain | 30/09/14 | UKHSA/PHE | Inns et al., 2016 |
| **SRR1968388** | Spain | Spain | 19/03/15 | UKHSA/PHE | Inns et al., 2016 |
| **SRR1968584** | Spain | Spain | 03/07/14 | UKHSA/PHE | Inns et al., 2016 |
| **SRR1969326** | Spain | Spain | 27/02/15 | UKHSA/PHE | Inns et al., 2016 |
| **SRR1969428** | Spain | Spain | 19/03/15 | UKHSA/PHE | Inns et al., 2016 |
| **SRR1969547** | Spain | Spain | 26/07/14 | UKHSA/PHE | Inns et al., 2016 |
| **SRR1969612** | Spain | Spain | 24/03/15 | UKHSA/PHE | Inns et al., 2016 |
| **SRR1969925** | Spain | Spain | 09/04/14 | UKHSA/PHE | Inns et al., 2016 |
| **SRR1970075** | Spain | Spain | 03/03/15 | UKHSA/PHE | Inns et al., 2016 |
| **SRR3048713** | Spain | Spain | 31/03/14 | UKHSA/PHE | Inns et al., 2016 |
| **SRR3049279** | Spain | Spain | 27/03/14 | UKHSA/PHE | Inns et al., 2016 |
| **SRR3049325** | Spain | Spain | 10/03/15 | UKHSA/PHE | Inns et al., 2016 |
| **SRR3049369** | Spain | Spain | 19/03/15 | UKHSA/PHE | Inns et al., 2016 |
| **SRR3049372** | Spain | Spain | 03/04/14 | UKHSA/PHE | Inns et al., 2016 |
| **SRR3049653** | Spain | Spain | 26/03/14 | UKHSA/PHE | Inns et al., 2016 |
| **SRR3120667** | Spain | Spain | 22/07/14 | UKHSA/PHE | Inns et al., 2016 |

| | | | | | |
|---|---|---|---|---|---|
| **SRR3284680** | Spain | Spain | 14/04/15 | UKHSA/PHE | Inns et al., 2016 |
| **SRR3284683** | Spain | Spain | 04/08/15 | UKHSA/PHE | Inns et al., 2016 |
| **SRR3284688** | Spain | Spain | 04/08/15 | UKHSA/PHE | Inns et al., 2016 |
| **SRR3284710** | Spain | Spain | 07/09/15 | UKHSA/PHE | Inns et al., 2016 |
| **SRR3284729** | Spain | Spain | 30/04/15 | UKHSA/PHE | Inns et al., 2016 |
| **SRR3284746** | Spain | Spain | 29/04/15 | UKHSA/PHE | Inns et al., 2016 |
| **SRR3284869** | Spain | Spain | 31/08/15 | UKHSA/PHE | Inns et al., 2016 |
| **SRR3285245** | Spain | Spain | 17/04/15 | UKHSA/PHE | Inns et al., 2016 |
| **SRR3285424** | Spain | Spain | 09/04/15 | UKHSA/PHE | Inns et al., 2016 |
| **SRR3285449** | Spain | Spain | 04/08/15 | UKHSA/PHE | Inns et al., 2016 |
| **SRR3285480** | Spain | Spain | 01/10/15 | UKHSA/PHE | Inns et al., 2016 |
| **SRR3285485** | Spain | Spain | 30/04/15 | UKHSA/PHE | Inns et al., 2016 |
| **SRR3286569** | Spain | Spain | 26/03/15 | UKHSA/PHE | Inns et al., 2016 |
| **SRR3286571** | Spain | Spain | 12/04/15 | UKHSA/PHE | Inns et al., 2016 |
| **SRR3286583** | Spain | Spain | 08/04/15 | UKHSA/PHE | Inns et al., 2016 |
| **SRR3286586** | Spain | Spain | 30/03/15 | UKHSA/PHE | Inns et al., 2016 |
| **SRR3286593** | Spain | Spain | 21/04/15 | UKHSA/PHE | Inns et al., 2016 |
| **SRR3286606** | Spain | Spain | 02/04/15 | UKHSA/PHE | Inns et al., 2016 |
| **SRR3286609** | Spain | Spain | 15/04/15 | UKHSA/PHE | Inns et al., 2016 |
| **SRR3286632** | Spain | Spain | 20/04/15 | UKHSA/PHE | Inns et al., 2016 |
| **SRR3286636** | Spain | Spain | 22/04/15 | UKHSA/PHE | Inns et al., 2016 |
| **SRR3286643** | Spain | Spain | 10/04/15 | UKHSA/PHE | Inns et al., 2016 |
| **SRR3286645** | Spain | Spain | 09/04/15 | UKHSA/PHE | Inns et al., 2016 |
| **SRR3286655** | Spain | Spain | 11/04/15 | UKHSA/PHE | Inns et al., 2016 |
| **SRR3286657** | Spain | Spain | 04/04/15 | UKHSA/PHE | Inns et al., 2016 |
| **SRR3286661** | Spain | Spain | 01/04/15 | UKHSA/PHE | Inns et al., 2016 |
| **SRR3286675** | Spain | Spain | 31/03/15 | UKHSA/PHE | Inns et al., 2016 |
| **SRR3286690** | Spain | Spain | 13/04/15 | UKHSA/PHE | Inns et al., 2016 |
| **SRR3286698** | Spain | Spain | 14/05/15 | UKHSA/PHE | Inns et al., 2016 |
| **SRR3286700** | Spain | Spain | 07/05/15 | UKHSA/PHE | Inns et al., 2016 |
| **SRR3286701** | Spain | Spain | 15/05/15 | UKHSA/PHE | Inns et al., 2016 |
| **SRR3286708** | Spain | Spain | 14/05/15 | UKHSA/PHE | Inns et al., 2016 |
| **SRR3286710** | Spain | Spain | 20/05/15 | UKHSA/PHE | Inns et al., 2016 |
| **SRR3286778** | Spain | Spain | 11/05/15 | UKHSA/PHE | Inns et al., 2016 |
| **SRR3286809** | Spain | Spain | 11/05/15 | UKHSA/PHE | Inns et al., 2016 |
| **SRR3286853** | Spain | Spain | 26/05/15 | UKHSA/PHE | Inns et al., 2016 |
| **SRR3286877** | Spain | Spain | 02/06/15 | UKHSA/PHE | Inns et al., 2016 |
| **SRR3286908** | Spain | Spain | 15/06/15 | UKHSA/PHE | Inns et al., 2016 |
| **SRR3286928** | Spain | Spain | 27/06/15 | UKHSA/PHE | Inns et al., 2016 |
| **SRR3312072** | Spain | Spain | 28/06/15 | UKHSA/PHE | Inns et al., 2016 |
| **SRR3312073** | Spain | Spain | 22/05/15 | UKHSA/PHE | Inns et al., 2016 |
| **SRR3312161** | Spain | Spain | 17/08/15 | UKHSA/PHE | Inns et al., 2016 |
| **SRR3312173** | Spain | Spain | 20/04/15 | UKHSA/PHE | Inns et al., 2016 |
| **SRR3312174** | Spain | Spain | 02/09/15 | UKHSA/PHE | Inns et al., 2016 |
| **SRR3312187** | Spain | Spain | 18/08/15 | UKHSA/PHE | Inns et al., 2016 |
| **SRR3312276** | Spain | Spain | 26/06/15 | UKHSA/PHE | Inns et al., 2016 |
| **SRR3312277** | Spain | Spain | 21/08/15 | UKHSA/PHE | Inns et al., 2016 |
| **SRR3312300** | Spain | Spain | 31/07/15 | UKHSA/PHE | Inns et al., 2016 |
| **SRR3312324** | Spain | Spain | 07/09/15 | UKHSA/PHE | Inns et al., 2016 |
| **SRR3312335** | Spain | Spain | 01/09/15 | UKHSA/PHE | Inns et al., 2016 |
| **SRR3312337** | Spain | Spain | 05/09/15 | UKHSA/PHE | Inns et al., 2016 |
| **SRR3312339** | Spain | Spain | 20/08/15 | UKHSA/PHE | Inns et al., 2016 |
| **SRR3312340** | Spain | Spain | 17/07/15 | UKHSA/PHE | Inns et al., 2016 |
| **SRR3312341** | Spain | Spain | 25/08/15 | UKHSA/PHE | Inns et al., 2016 |
| **SRR3312342** | Spain | Spain | 23/07/15 | UKHSA/PHE | Inns et al., 2016 |
| **SRR3312343** | Spain | Spain | 22/06/15 | UKHSA/PHE | Inns et al., 2016 |
| **SRR3312344** | Spain | Spain | 02/09/15 | UKHSA/PHE | Inns et al., 2016 |
| **SRR3312345** | Spain | Spain | 18/08/15 | UKHSA/PHE | Inns et al., 2016 |
| **SRR3312348** | Spain | Spain | 24/04/15 | UKHSA/PHE | Inns et al., 2016 |
| **SRR3312356** | Spain | Spain | 24/08/15 | UKHSA/PHE | Inns et al., 2016 |

| | | | | | |
|---|---|---|---|---|---|
| **SRR3312492** | Spain | Spain | 07/08/15 | UKHSA/PHE | Inns et al., 2016 |
| **SRR3312512** | Spain | Spain | 20/09/15 | UKHSA/PHE | Inns et al., 2016 |
| **SRR3312513** | Spain | Spain | 22/04/15 | UKHSA/PHE | Inns et al., 2016 |
| **SRR3312514** | Spain | Spain | 13/05/15 | UKHSA/PHE | Inns et al., 2016 |
| **SRR3312520** | Spain | Spain | 28/08/15 | UKHSA/PHE | Inns et al., 2016 |
| **SRR3312525** | Spain | Spain | 28/08/15 | UKHSA/PHE | Inns et al., 2016 |
| **SRR3312528** | Spain | Spain | 05/06/15 | UKHSA/PHE | Inns et al., 2016 |
| **SRR3312529** | Spain | Spain | 11/09/15 | UKHSA/PHE | Inns et al., 2016 |
| **SRR3312530** | Spain | Spain | 04/08/15 | UKHSA/PHE | Inns et al., 2016 |
| **SRR3312531** | Spain | Spain | 01/09/15 | UKHSA/PHE | Inns et al., 2016 |
| **SRR3312533** | Spain | Spain | 04/09/15 | UKHSA/PHE | Inns et al., 2016 |
| **SRR3312545** | Spain | Spain | 27/08/15 | UKHSA/PHE | Inns et al., 2016 |
| **SRR3312546** | Spain | Spain | 05/08/15 | UKHSA/PHE | Inns et al., 2016 |
| **SRR3312547** | Spain | Spain | 27/08/15 | UKHSA/PHE | Inns et al., 2016 |
| **SRR3312548** | Spain | Spain | 10/09/15 | UKHSA/PHE | Inns et al., 2016 |
| **SRR3312549** | Spain | Spain | 03/05/15 | UKHSA/PHE | Inns et al., 2016 |
| **SRR3312550** | Spain | Spain | 02/09/15 | UKHSA/PHE | Inns et al., 2016 |
| **SRR3312551** | Spain | Spain | 16/04/15 | UKHSA/PHE | Inns et al., 2016 |
| **SRR3312559** | Spain | Spain | 15/04/15 | UKHSA/PHE | Inns et al., 2016 |
| **SRR3312576** | Spain | Spain | 21/05/15 | UKHSA/PHE | Inns et al., 2016 |
| **SRR3312577** | Spain | Spain | 24/09/15 | UKHSA/PHE | Inns et al., 2016 |
| **SRR3312578** | Spain | Spain | 29/04/15 | UKHSA/PHE | Inns et al., 2016 |
| **SRR3312579** | Spain | Spain | 08/09/15 | UKHSA/PHE | Inns et al., 2016 |
| **SRR3312589** | Spain | Spain | 04/09/15 | UKHSA/PHE | Inns et al., 2016 |
| **SRR3312591** | Spain | Spain | 01/09/15 | UKHSA/PHE | Inns et al., 2016 |
| **SRR3312597** | Spain | Spain | 21/08/15 | UKHSA/PHE | Inns et al., 2016 |
| **SRR3312604** | Spain | Spain | 01/09/15 | UKHSA/PHE | Inns et al., 2016 |
| **SRR3319055** | Spain | Spain | 26/08/15 | UKHSA/PHE | Inns et al., 2016 |
| **SRR3319056** | Spain | Spain | 24/08/15 | UKHSA/PHE | Inns et al., 2016 |
| **SRR3319058** | Spain | Spain | 09/09/15 | UKHSA/PHE | Inns et al., 2016 |
| **SRR3319059** | Spain | Spain | 30/08/15 | UKHSA/PHE | Inns et al., 2016 |
| **SRR3319061** | Spain | Spain | 26/08/15 | UKHSA/PHE | Inns et al., 2016 |
| **SRR3319062** | Spain | Spain | 27/08/15 | UKHSA/PHE | Inns et al., 2016 |
| **SRR3319065** | Spain | Spain | 30/09/15 | UKHSA/PHE | Inns et al., 2016 |
| **SRR3319066** | Spain | Spain | 29/04/15 | UKHSA/PHE | Inns et al., 2016 |
| **SRR3319067** | Spain | Spain | 23/07/15 | UKHSA/PHE | Inns et al., 2016 |
| **SRR3319068** | Spain | Spain | 14/08/15 | UKHSA/PHE | Inns et al., 2016 |
| **SRR3319069** | Spain | Spain | 10/05/15 | UKHSA/PHE | Inns et al., 2016 |
| **SRR3319071** | Spain | Spain | 24/08/15 | UKHSA/PHE | Inns et al., 2016 |
